# Genetic and environmental regulation of caudate nucleus transcriptome: insight into schizophrenia risk and the dopamine system

**DOI:** 10.1101/2020.11.18.20230540

**Authors:** Kynon JM Benjamin, Arthur S Feltrin, André Rocha Barbosa, Andrew E Jaffe, Leonardo Collado-Torres, Emily E Burke, Joo Heon Shin, William S Ulrich, Amy Deep-Soboslay, Ran Tao, the BrainSeq Consortium, Thomas M Hyde, Joel E Kleinman, Jennifer A Erwin, Daniel R Weinberger, Apuã CM Paquola

## Abstract

Increased dopamine (DA) signaling in the striatum has been a cornerstone hypothesis about psychosis for over 50 years. Increased dopamine release results in psychotic symptoms, while D2 dopamine receptor (DRD2) antagonists are antipsychotic. Recent schizophrenia GWAS identified risk-associated common variants near the DRD2 gene, but the risk mechanism has been unclear. To gain novel insight into risk mechanisms underlying schizophrenia, we performed a comprehensive analysis of the genetic and transcriptional landscape of schizophrenia in postmortem caudate nucleus from a cohort of 444 individuals. Integrating expression quantitative trait loci (eQTL) analysis, transcriptome wide association study (TWAS), and differential expression analysis, we found many new genes associated with schizophrenia through genetic modulation of gene expression. Using a new approach based on deep neural networks, we construct caudate nucleus gene expression networks that highlight interactions involving schizophrenia risk. Interestingly, we found that genetic risk for schizophrenia is associated with decreased expression of the short isoform of *DRD2*, which encodes the presynaptic autoreceptor, and not with the long isoform, which encodes the postsynaptic receptor. This association suggests that decreased control of presynaptic DA release is a potential genetic mechanism of schizophrenia risk. Altogether, these analyses provide a new resource for the study of schizophrenia that can bring insight into risk mechanisms and potential novel therapeutic targets.

## Main Text

Schizophrenia is a devastating neuropsychiatric disorder that affects ∼1% of the world population. It is characterized by positive symptoms (such as hallucinations and delusions), negative symptoms (such as avolition and withdrawal) and cognitive dysfunction (*1*). Clinical evidence suggests that excessive dopaminergic modulation of striatal function may mediate psychosis, which has been a central hypothesis for sixty years. Dopamine (DA) was the first neurotransmitter implicated in schizophrenia, and the efficacy of most antipsychotic drugs are highly correlated with their ability to block dopamine D2 receptors in striatum (*2*). Yet, until the results of recent genetic association studies, it was unclear whether dopamine was of primary pathogenic importance or merely a secondary association with illness.

Schizophrenia is highly heritable, and genetic studies are playing a pivotal role in identifying potential biomarkers and causal disease mechanisms with the hope of informing new treatments. Recent genome-wide association studies (GWAS) (*3, 4*) identified nearly one hundred and fifty loci associated with schizophrenia risk, one of which includes the gene *DRD2*, which encodes the dopamine D2 receptor. While this association implicates DRD2 as a risk factor, translating this statistical genetic information into a pathogenic mechanism remains a significant challenge. Gene expression studies in human postmortem brain tissue such as the BrainSeq, PsychENCODE and CommonMind consortia have generated neurogenomic datasets that link functional genomic elements and molecules to the biology of brain disorders. These efforts have created maps and integrated models of widespread genetic, developmental, brain region and schizophrenia-associated changes in gene expression, isoform regulation, and cell type proportions (*5, 6*). So far, however, they have failed to identify a molecular association and potential mechanism related to the DRD2 locus. This failure to date may be explained at least in part by the fact that these studies have focused almost exclusively on cortical areas (*7, 8*) where dopamine D2 receptors are very lowly expressed, even though subcortical regions are also prominently implicated in schizophrenia pathogenesis (*9–12*). In this study, we performed a comprehensive analysis of the genetic and transcriptional landscape of schizophrenia in postmortem caudate nucleus where DRD2 expression is abundant (Fig. 1).

**Fig. 1:**
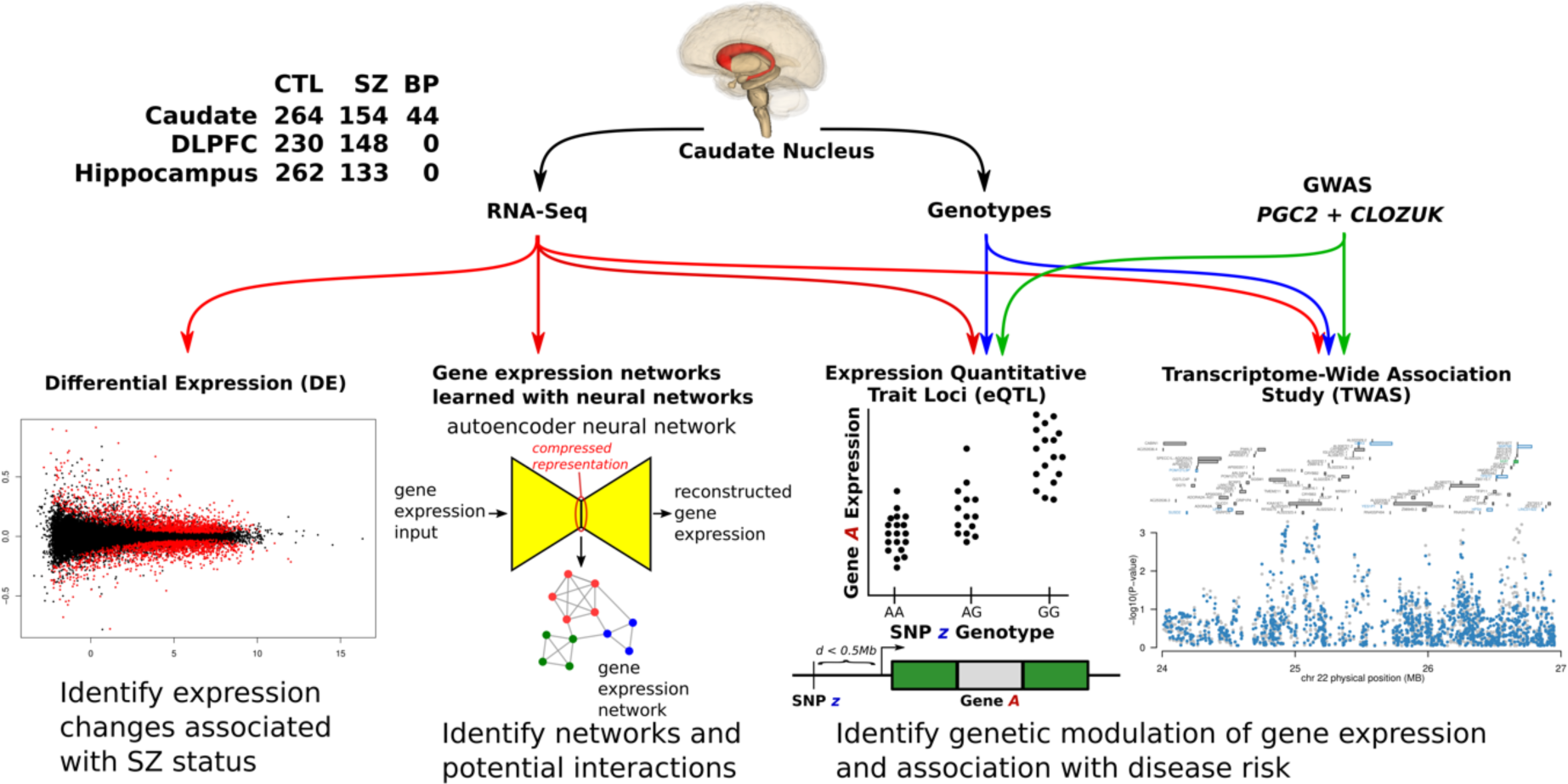
Overview of computational analysis. Using genotypes and RNA-sequencing data from postmortem caudate nucleus from 444 individuals, we interrogate genes, transcripts, exons, and exon-exon junctions for associations with schizophrenia. We perform eQTL and TWAS analyses to identify genetic modulation of gene expression, integrating with genetic risk information from GWAS. We perform differential expression analysis to identify expression changes associated with disease status. We integrate our analysis with previously published DLPFC and hippocampus data. Using a new approach based on deep neural networks, we construct gene expression networks to gain insight into interactions involving schizophrenia risk genes and uncover potential novel therapeutic targets.

## Results

### Genetic regulation of gene expression in the caudate nucleus

To gain insight into how genetic risk for schizophrenia manifests in changes in RNA expression, we set out to identify expression quantitative trait loci (eQTLs) in the caudate nucleus and compare them with eQTLs found in hippocampus and DLPFC in the BrainSeq Phase II dataset (*6*) and the CommonMind Consortium (CMC) DLPFC dataset (*13*), as well as in the smaller caudate GTEx dataset (*14*). Using genotypes and RNA-Seq data from the Lieber Institute for Brain Development (LIBD) brain repository, we identified cis-eQTLs for genes, transcripts, exons and exon-exon junctions for 444 individuals (age > 13, 246 controls, 154 schizophrenia patients, 44 bipolar patients) using a false discovery rate (FDR) cutoff of 5% (Data S1, Table S1) and identified eQTLs for SNPs associated with schizophrenia risk (GWAS p-value < 5e-8) based on the current PGC2+CLOZUK GWAS (*3*) (Fig. 2A and Table S1). The top 10 caudate cis-eQTL for genes (*XRRA1*, *RPS26*, *ERAP2*, *NSA2*, *AP000350.6*, *ITGB3BP*, *SPATA7*, *AC092821.1*, *GABPB1-AS1*, and *CUTALP*) are shown in Fig. S1, whereas the top 10 gene eQTLs with SNPs associated with schizophrenia risk (*SDAD1P1*, *ALMS1P1*, *PPP1R13B*, *PCCB*, *TYW5*, *GNL3*, *THOC7*, *TOM1L2*, *SNX19*, and *SRR*) are shown in Fig. S2.

**Fig. 2:**
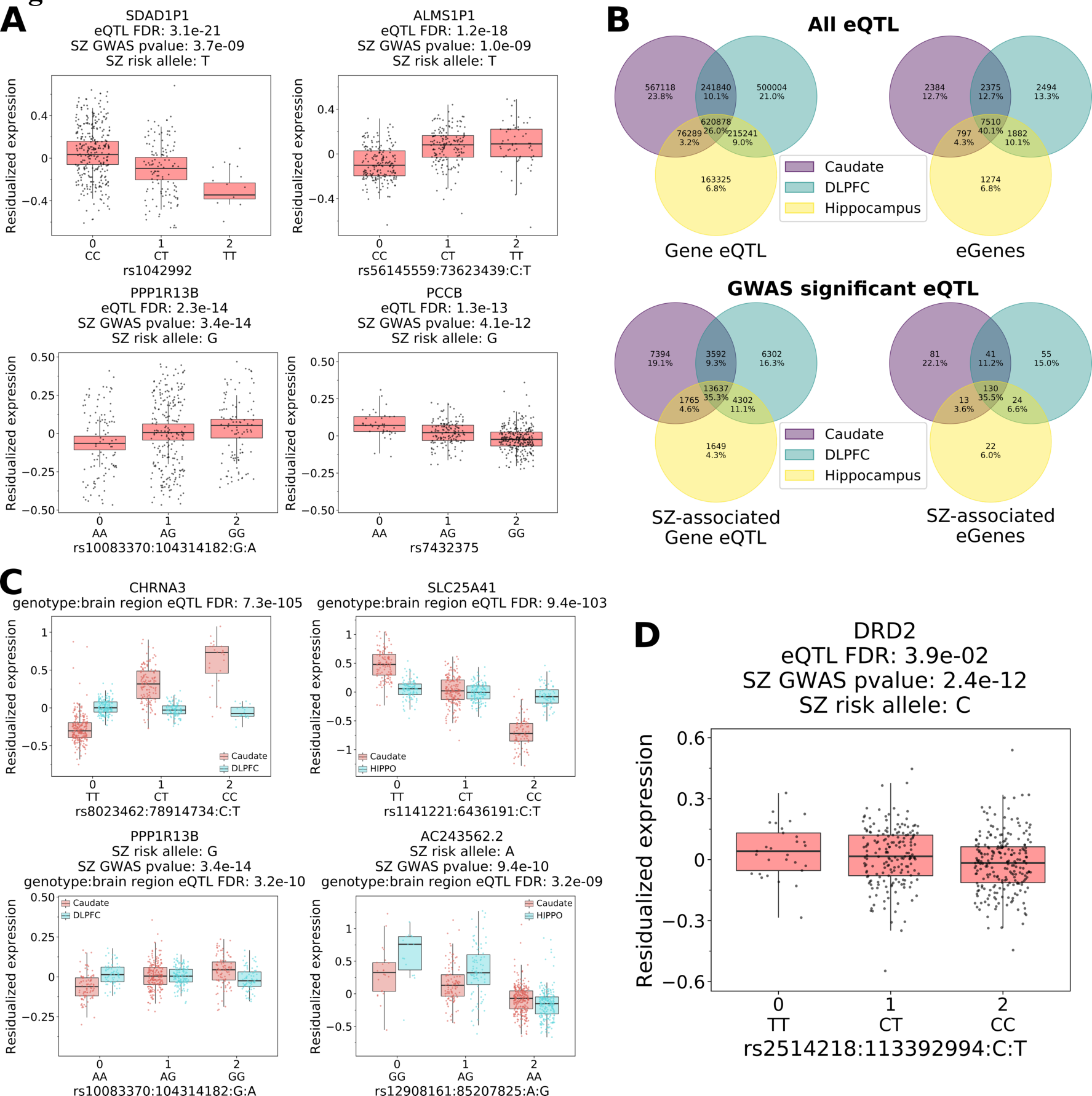
Genetic regulation of expression in the caudate nucleus. **(A)** The four most significant gene-level cis-eQTLs associated with a schizophrenia risk index SNP according to PGC2 + CLOZUK GWAS (*3*). **(B)** Comparative analysis of eQTLs and eGenes across brain regions (top) and those associated with schizophrenia risk (bottom), FDR < 0.01. **(C)** Representative boxplot of gene-level brain region-dependent cis-eQTL (top) comparing caudate with DLPFC (left) and hippocampus (right), and cis-eQTL for GWAS-significant index SNPs (bottom). **(D)** Dopamine receptor D2 gene cis-eQTL is associated with schizophrenia risk (eQTL, FDR < 0.05) in the caudate nucleus.

To understand the regional specificity of caudate eQTLs, we compared significant gene-level cis-eQTLs in caudate (at FDR < 0.01 and 0.05) to those reported as statistically significant in BrainSeq Phase II DLPFC and Hippocampus (FDR < 0.01) (*6*), GTEx caudate (FDR < 0.05) and CMC DLPFC (FDR < 0.05) (*13*) with respect to direction of effect. The replication rate for significant eQTLs in these other datasets ranged from 9.4% to 57.3% (Tables S2-S3). The vast majority (>98.0%) of matching eQTLs have concordant directionality across different brain regions and across studies that use different eQTL identification methodology, suggesting that most cis-eQTLs have an intrinsic genotype to gene expression directionality relationship that is independent of brain region or cell type.

We next asked about the proportion of eQTLs detected in one or across multiple brain regions. We compared the caudate nucleus eQTLs with the hippocampus and DLPFC eQTLs reported in BrainSeq Phase II (*6*). 26.7% of gene level eQTLs (40.1% of genes with eQTLs, or eGenes) are detected (at FDR < 0.01) in all three brain regions, 22.3% of eQTLs (27.1% of eGenes) are detected in two brain regions and 51% of eQTLs (32.8% of eGenes) are detected in one brain region alone (Fig. 2B). Similarly, eQTLs with SNPs significantly associated with schizophrenia risk (GWAS p-value < 5e-08) (*3*) were also generally shared across multiple brain regions (Fig. 2B). This pattern was observed in transcripts, exons, and junction analyses (Fig. S3-S5).

To identify brain region-dependent eQTLs, we combined data from pairs of brain regions and performed eQTL mapping testing for statistical interaction between genotype and brain region (FDR < 0.05). For caudate and DLPFC, we found 354,474 gene-level brain region-dependent eQTLs, involving 6,510 genes, 79 of which are associated with schizophrenia risk loci. For caudate and hippocampus, we found 204,016 gene-level brain region-dependent eQTLs, involving 4,710 genes, 45 of which are associated with schizophrenia risk loci. For DLPFC and hippocampus, we found 86,980 gene-level brain region-dependent eQTLs, involving 2,247 genes, 16 of which are associated with schizophrenia risk. Table S4 shows the numbers of gene, transcript, exon, and junction eQTLs found for each pair of brain regions. Examples of the top region-dependent eQTLs and eQTLs with SNPs significantly associated with schizophrenia risk (GWAS p-value<5e-08) for caudate compared to DLPFC and hippocampus are shown in Fig. 2C. For instance, *PPP1R13B* expression increases with schizophrenia risk allele G (SNP: rs10083370) in caudate, while it decreases slightly in DLPFC. Interestingly, there were twice as many region-dependent eQTLs in pairs involving caudate (Table S4), suggesting that caudate has a more dissimilar gene regulation structure than DLPFC and hippocampus.

All eQTL analyses are available for visualization on the LIBD eQTL browser (http://erwinpaquolalab.libd.org/caudate_eqtl/) and for download on supplemental Data S1.

### Reduced expression of the short isoform of *DRD2* in caudate is associated with genetic risk for schizophrenia

Excessive activity of the striatal dopamine system has been long implicated as playing a pivotal role in schizophrenia and its treatment (*15–19*). Thus, we examined the eQTL results for dopamine receptor D2 associated SNPs with schizophrenia risk in the caudate nucleus (Fig. 2D). The *DRD2* gene level eQTL analysis indicates that the schizophrenia risk associated allele at the GWAS index SNP rs2514218 predicts decreased expression for this gene, on its surface a counterintuitive result.

We thus examined the receptor’s genetic association with expression in greater detail. The dopamine receptor D2 generates two principal isoforms, D2L (long) and D2S (short) via alternative splicing of exon 6 (*20–22*) with different localization and function (Fig. 3A). D2L has postsynaptic receptor function whereas D2S has autoreceptor function, is present primarily in presynaptic terminals of dopaminergic neurons, and participates in a negative feedback circuit that regulates production and release of dopamine from terminals (*23, 24*) (Fig. 3A).

**Fig. 3:**
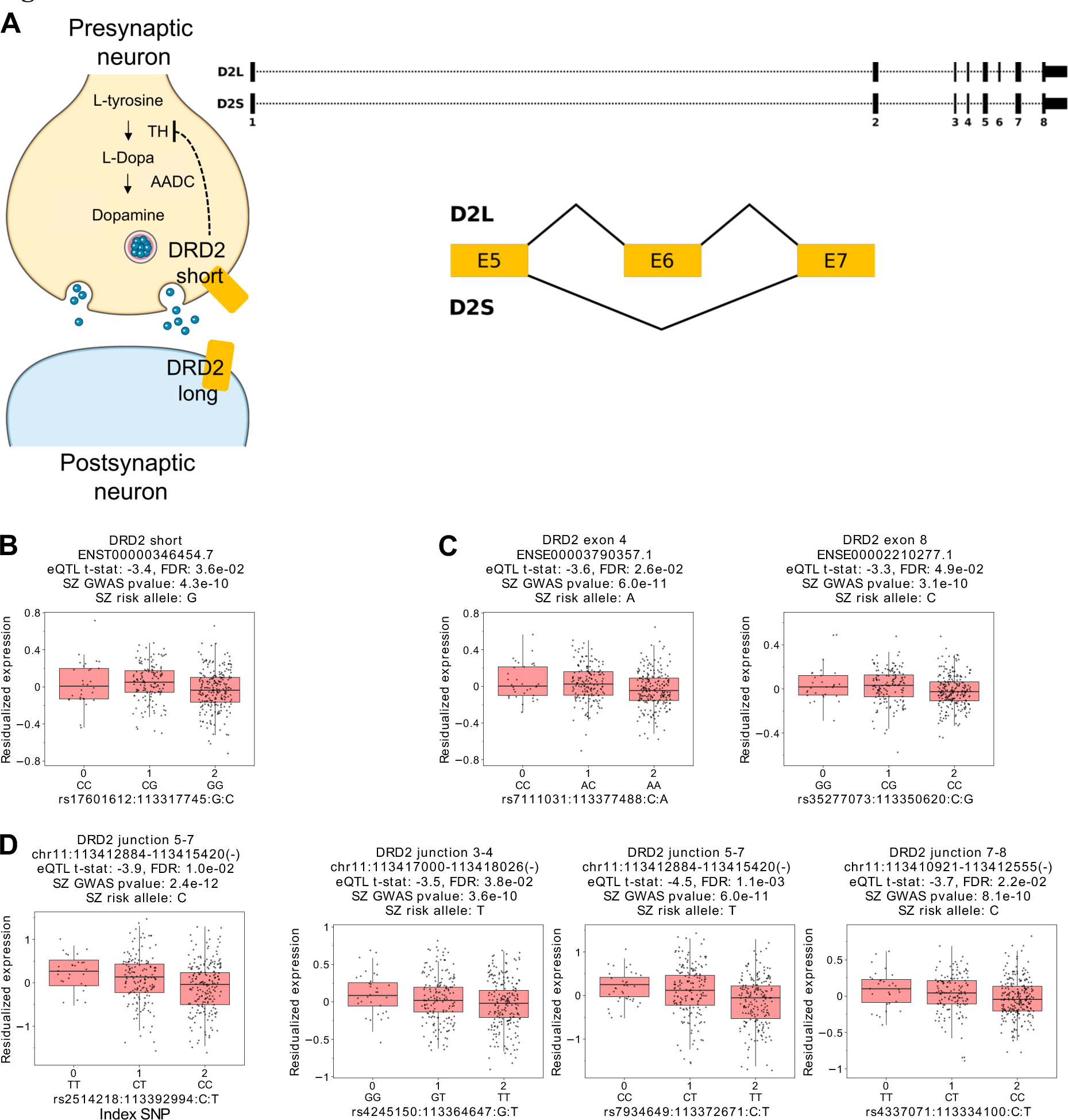
Genetic regulation of dopamine receptor D2 associated with short isoform (D2S) in the caudate nucleus. **(A)** Schematic of DRD2 isoforms and mechanism of action. **(B)** Transcript-level eQTL analysis identifies SNPs associated with schizophrenia risk and expression changes only for D2S. **(C)** Exon-level eQTL analysis identifies SNPs associated with schizophrenia risk and expression changes for exons shared between short (D2S) and long (D2L) isoforms. **(D)** Junction-level analysis identifies SNPs associated with schizophrenia risk and DE specific to D2S. (eQTL FDR < 0.05, GWAS P value < 5e-8).

On the transcript level, we identified eQTLs with SNPs associated with schizophrenia risk for the short isoform of *DRD2*, while no significant eQTLs were found in GWAS-significant SNPs for the long isoform (Fig. 3B). As with the *DRD2* gene level analysis, eQTL analysis indicates decreased expression of the short isoform is associated to increased schizophrenia risk (Fig. 3B).

To further validate these results, we performed eQTL analysis for individual *DRD2* exons, which are shown in Fig. 3C. We found eQTLs for GWAS-significant SNPs only for exon 4 and 8, which are present in both long and short isoforms (Fig. 3C and Fig. S7). Similar to the full gene and transcript models, reduced expression is associated with schizophrenia risk.

As there are no exons specific to D2S, we performed additional analysis of eQTL for individual exon-exon junctions, which do show specificity. Here, we found exclusive association of D2S with schizophrenia risk as the D2S-specific junction between exons 5 and 7 showed association with schizophrenia risk SNPs (Fig. 3D and Fig. S9). These results converge on the conclusion that reduced expression of the short isoform of *DRD2* in the caudate nucleus is associated with genetic risk for schizophrenia. In prefrontal cortex and hippocampus, where *DRD2* expression, particularly of the D2S isoform, is very low in abundance (Fig. S10), we did not find significant eQTLs with GWAS associations (*8*).

Interestingly, one exon of *ANKK1*, a genomic neighbor of *DRD2*, also has a significant eQTL association with index SNP rs2514218 (FDR = 8.4*10^-3^, Fig. S8A). We thus compared its expression with *DRD2* junction 5-7 to identify possible independent associations. As shown in Fig. S8B there is no significant correlation between the expression of this *ANKK1* exon with *DRD2* junction 5-7 conditional on the genotype at SNP rs2514218, suggesting they are independently regulated, factoring out the genetic influence of this locus.

We compared our *DRD2* results with eQTLs from the GTEx caudate nucleus dataset (*16*), which has a smaller number of samples (194 samples with genotypes) than our dataset. At the gene level analysis GTEx did not identify eQTL associations with *DRD2*. Because GTEx does not provide eQTL calls for exon-exon junctions, we applied our eQTL calling methodology on caudate exon-exon junction RNA-Seq read counts and covariates provided by GTEx. Although the association of *DRD2* junction 5-7 and schizophrenia GWAS index SNP rs2514218 was found to be not statistically significant (p=0.72), the direction of effect was the same as in our data. To determine if the smaller sample size might explain why this association does not reach statistical significance in GTEx, we performed 10 random samplings of our data (n=444) to GTEx sample size (n=194) and calculated eQTLs for each of them. For only 2 out of 10 samplings, SNP rs2514218 had a statistically significant association *DRD2* junction 5-7 (FDR < 0.05), suggesting the smaller sample size in GTEx could explain, at least in part, the difference to our dataset.

### Transcriptome-wide association study in caudate identifies new genes associated with schizophrenia risk

Leveraging our transcriptional and genetic datasets with schizophrenia GWAS summary statistics (*3*), we sought to prioritize candidate schizophrenia risk genes by applying transcriptome-wide association study (TWAS) methodology (*25*). We identified 5256 genes, 9735 transcripts, 39467 exons and 14226 junctions with heritable expression (heritability p-value < 0.01). From these features, we identified 489 genes, 933 transcripts, 3907 exons and 1419 junctions with significant TWAS association for schizophrenia (FDR < 0.05) (Table S5 and Data S2).

We examined the TWAS results for associations with *DRD2* and found two of its genomic features to be heritable. One is exon 2 of isoform *DRD2-006* (ENSE00002234274.1), which has a non-significant TWAS association (p-value=0.65, FDR=1.0). Interestingly, the other heritable feature is *DRD2* junction 5-7 (chr11:113,412,884-113,415,420, belonging specifically to the short isoform), which has a nominally significant TWAS association (p-value=0.007, FDR=0.067), further supporting the association specifically of *DRD2* short autoreceptor isoform with genetic risk for schizophrenia and downregulation being the operant directionality.

Next, we applied the hypergeometric test to identify Gene Ontology (GO) terms enriched among TWAS-significant genes. We found significant enrichment (FDR < 0.05) for Golgi-associated vesicle, endoplasmic reticulum membrane, MHC protein complex and antigen processing and presentation (Fig. S12). These results are somewhat divergent from GO term enrichment analyses on TWAS gene sets based on gene expression in cortical regions which have emphasized synaptic function and neurodevelopmental processes (*3, 6*).

Interestingly, and consistent with the GO analyses, the comparison among TWAS genes for caudate, DLPFC, and hippocampus also revealed that a number of TWAS genes were only significant for caudate, while others were shared across tissues as shown, respectively, in red and blue in the Manhattan plot in Fig. 4C. Comparing the caudate nucleus TWAS results with those of hippocampus and DLPFC (*6*), we observed considerable overlap among the three brain regions for significant gene-level TWAS associations as well as overlap of heritable genes (Fig. 4A). Moreover, we also found a significant enrichment with significant schizophrenia TWAS associations (*26*) (Fisher’s Exact Test, p-value < 0.01; Data S3). For each pair of brain regions, the vast majority of genes that have significant TWAS association have concordant directionality of association, as shown in Fig. 4B, which is also observed between DLPFC and hippocampus (*6*). This observation is in line with the eQTL directionality comparison with other studies (Table S2-3), also suggesting each SNP-gene pair has an intrinsic genotype and expression directionality relationship that is independent of brain region and cell types that distinguish regions. For example, *C12orf65*, a mitochondria related gene, is a highly significant TWAS gene in caudate with increased expression associated with schizophrenia risk and is also shared among the two other tissues. We found that 48 of the 70 overlapping TWAS significant genes shared across tissues did not reach GWAS significance in the clinical GWAS study (Data S3). For these top genes, *CORO7* (coronin 7), plays an essential role in maintenance of Golgi apparatus morphology. We found *GLG1* (Golgi Glycoprotein 1) shared between caudate and hippocampus, and *JKAMP,* which might regulate the duration of MAPK8 activity in response to various stress stimuli, shared between caudate and DLPFC.

**Fig. 4:**
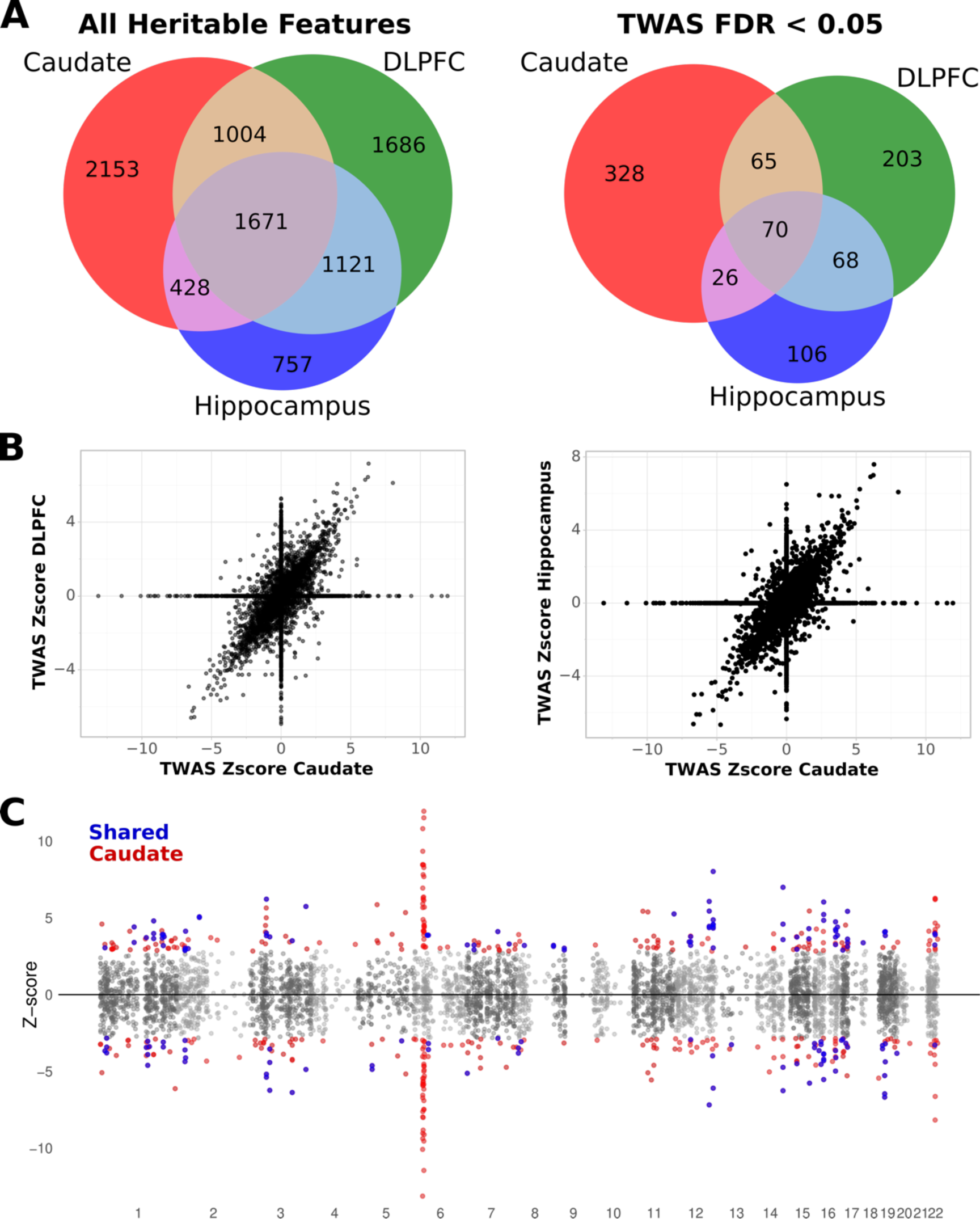
The majority of TWAS associations have same direction of effect across brain regions. **(A)** Comparison of TWAS associations, all heritable genes (left) significant genes (FDR < 0.05; right) between caudate nucleus, DLPFC, and hippocampus. **(B)** The vast majority of heritable genes have concordant directionality between brain regions. **(C)** Manhattan plot of schizophrenia TWAS associations for the caudate nucleus. Each point represents an individual heritable gene physical position on the x-axis and signed Z-score for each association on the y- axis. Blue points are significant TWAS associations (FDR < 0.05) shared between caudate, DLPFC, and hippocampus. Red points are caudate significant TWAS associations.

Remarkably, however, we found 252 TWAS genes unique to caudate compared with other schizophrenia TWAS analyses (*6, 7, 27*), where 148 of these genes did not reach GWAS significance in the clinical GWAS sample (Table S6). Of these *MIAT*, also known as Gomafu, has been linked to schizophrenia in other analyses (*28–30*). Interestingly, previous work has shown that *MIAT* positively regulates *GDNF* (*31*) and our differential expression results (below) found that both *MIAT* and *GDNF* were significantly upregulated in schizophrenia (Data S4).

These region selective TWAS findings underscore that the mechanisms of genetic risk for schizophrenia are not solely represented in one brain region or functional circuit, but implicate distributed brain systems that mediate diverse information processing streams.

### Differential expression in the caudate nucleus in schizophrenia

Despite the caudate nucleus having been associated with schizophrenia and being a likely principal target of antipsychotic medication, no large-scale analysis in the caudate nucleus has identified differentially expressed RNA features in patients with schizophrenia compared to neurotypical individuals. Here, we analyzed RNA-Seq data from 394 individuals of age 17 and older, 154 of them diagnosed with schizophrenia and 240 of them neurotypical controls. After quantifying genes, transcripts, exons, and junctions from the RNA-Seq reads, we performed differential expression analysis using limma-voom in the R environment, adjusting for sex, age, population stratification and RNA quality metrics, including RNA degradation metrics obtained with the qSVA methodology (*32*) taking into account degradation experiments performed on caudate nucleus samples.

We observed extensive differential gene expression for schizophrenia (2699 genes at FDR < 0.05) with *GDNF-AS1* (glial cell derived neurotrophic factor antisense RNA 1) and *TH* (tyrosine hydroxylase) as the top up- and down-regulated genes, respectively (Fig. 5A). As shown in the KEGG pathway map of the dopaminergic signaling pathway, *TH* – the rate limiting enzyme in dopamine production – and the dopamine receptors D2 and D3 were differentially expressed (Fig. S13). While we identified cis-eQTLs for D2 as noted above, there were no significant eQTLs (FDR < 0.05) for D3, D4, or TH. A summary of differentially expressed (DE) features can be found in Table S7.

**Fig. 5:**
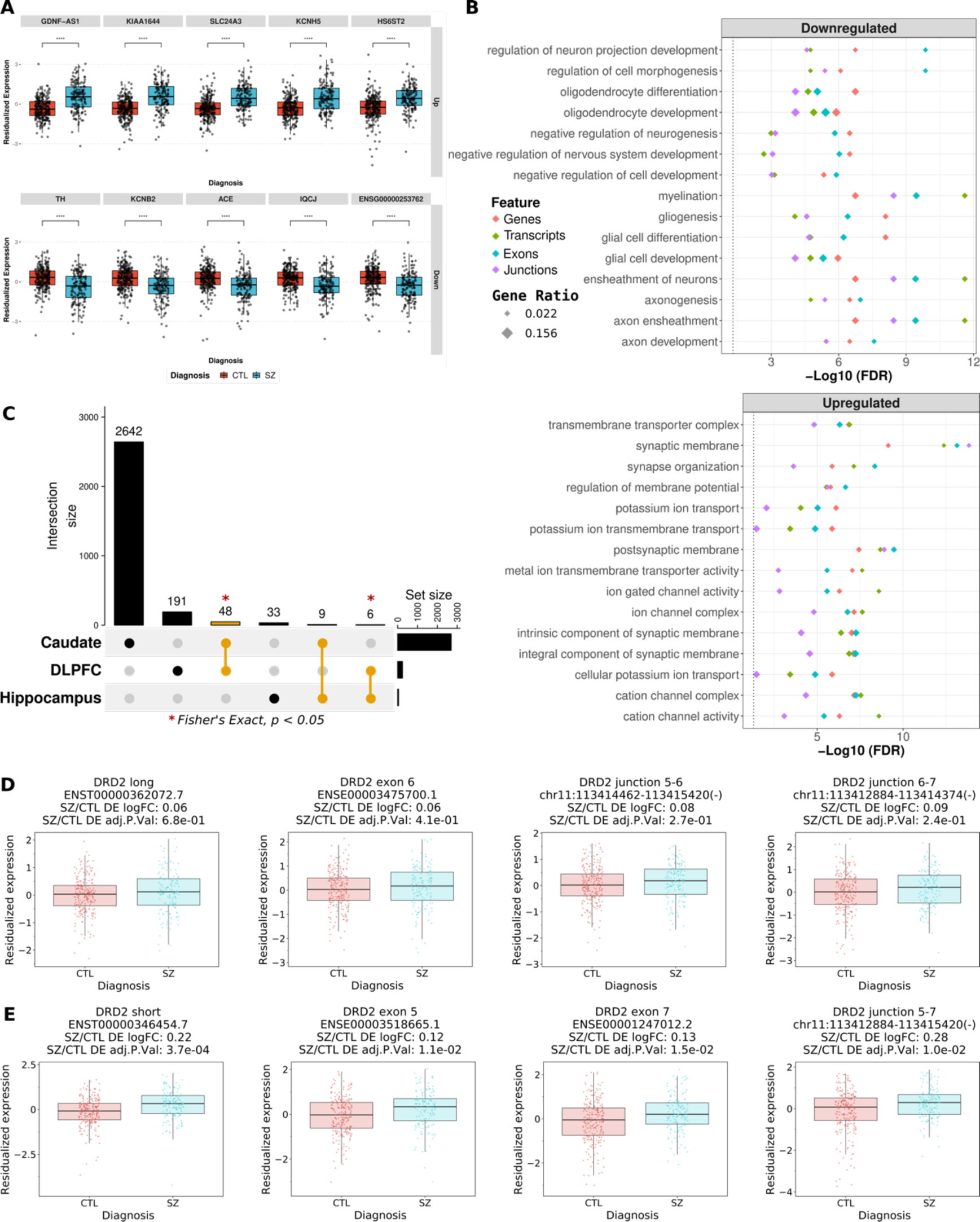
Widespread upregulation of neuronal signaling and downregulation of neural differentiation & development in the schizophrenia caudate nucleus. **(A)** Boxplots of the top 5 up- and downregulated genes in the caudate nucleus of patients compared with controls, ordered by p-value from left to right. Control (red), Schizophrenia (blue). **(B)** Top 15 up- and downregulated GO enriched terms shared across all features. **(C)** Upset plot comparing differentially expressed genes of caudate (FDR < 0.05) to DLPFC and hippocampus (*6*) (FDR < 0.05) showing brain region-specific differential expression. * statistically significant pairwise overlap of DE genes (Fisher’s exact test p-value < 0.05). Boxplot of differential expression analysis on the transcript, exon, and junction levels (**D**) specific to DRD2 long isoform, or (**E**) associated with DRD2 short isoform.

To identify biological themes associated with differentially expressed genes in schizophrenia, we performed the hypergeometric test and gene set enrichment analysis for term enrichment against the GO database. The upregulated features are enriched for synapse organization and ion transport whereas the downregulated features are enriched for gliogenesis, myelination, and negative regulation of neurogenesis (Fig. 5B). These results, which notably diverge from those related to genetic risk, suggest, perhaps not surprisingly, that in postmortem analysis of schizophrenia brain, the disease, and its consequences, including treatment and lifestyle changes, may have a major impact on different structural and functional properties of the caudate nucleus.

We next compared differentially expressed genes in schizophrenia in caudate with that of DLPFC and hippocampus in BrainSeq samples (Fig. 5C). The caudate nucleus has substantially more DE genes (2699 DEGs, FDR < 0.05) compared with DLPFC and hippocampus (245 and 48 DEGs, respectively (*6*)). While the majority of DEGs are region-specific and there is remarkably no DEG overlap for all three brain regions, there is statistically significant pairwise overlaps between caudate and DLPFC (p=4.2*10^-4^, Fisher’s exact test) and between DLPFC and hippocampus (p=1.2*10^-5^, Fisher’s exact test). There is also a significant positive pairwise correlation for DE t-statistics (Spearman p-value < 0.001; ρ = 0.24 and 0.15 for caudate comparison with DLPFC and hippocampus respectively; Fig. S14). It is further noteworthy that among the genes that are DE in two brain regions, several have discordant direction of effect for schizophrenia (Fig. S15), highlighting the importance of studying multiple brain regions when searching for targets for drug development.

Interestingly, the *DRD2* long isoform did not show differential expression in schizophrenic individuals compared to neurotypical controls (Fig. 5D), whereas the short isoform is upregulated in the caudate nucleus of schizophrenic individuals. Consistent with this, for exons 2, 3, 4, 5, 7 and 8, which are present in both long and short isoforms, we observe a similar increase in expression (log2 fold change 0.13, FDR < 0.015; Fig. 5D-E and Fig. S7) in schizophrenic individuals, whereas for exon 6, which is only present in the long isoform, the difference in expression (log2 fold change 0.06; Fig. 5D) is not statistically significant (FDR = 0.41). Furthermore, only the junction associated with D2S and not junctions specific to D2L (5-6, 6-7) were upregulated in schizophrenia individuals (Fig. 5E and Fig. S9). These data suggest opposing associations of trait (i.e., downregulation) and state (i.e., upregulation) with expression of D2S.

### Integration of GWAS, DE, and eQTL identifies 46 genes associated with both risk and clinical state

Observable expression differences between cases and controls originate from a combination of genetic and environmental factors, likely potential consequences of chronicity and treatment, as well as technical factors and study biases. While correction for confounding factors can lead to statistical associations that are more closely related to causal relationships, observational studies alone are not sufficient to characterize the causal structure leading to disease (*33*). eQTL and TWAS analyses help tease out the genetic contribution to gene expression and link it with disease risk through integration with GWAS. By combining eQTL and differential expression, we can identify, for each brain region, expressed features that are associated both with disease risk as well as clinical state and interrogate their direction of effect to inform the direction of potential therapeutic modulation.

For genes, transcripts, exons, and junctions, we combined the cis-eQTL associations (SNP-feature pairs at FDR<0.05) with the differential expression data in the caudate nucleus (schizophrenia vs control adjusted p-value < 0.05) with all GWAS-significant risk SNPs for schizophrenia (index and proxy SNPs with p-value < 5e-8) provided by the latest schizophrenia GWAS (*3*). The intersection of differentially expressed genes and genes with eQTLs in GWAS-significant risk SNPs identified 46 genes as shown in Fig. 6A with genes also overlapping with significant TWAS associations shown in bold. Out of the 46 genes reported, 23 have differential expression directionality concordant with the direction of schizophrenia risk according to GWAS, and 23 have discordant directionality. Four examples of genes in this intersection are C4A, *CACNA1I*, *CNNM2*, and *HIRIP3* (Fig. 6B). For instance, *CACNA1I*, which encodes a low-voltage-activated calcium channel, has decreased expression associated with schizophrenia risk according to eQTL analysis and observed decreased expression in affected patients.

**Fig. 6:**
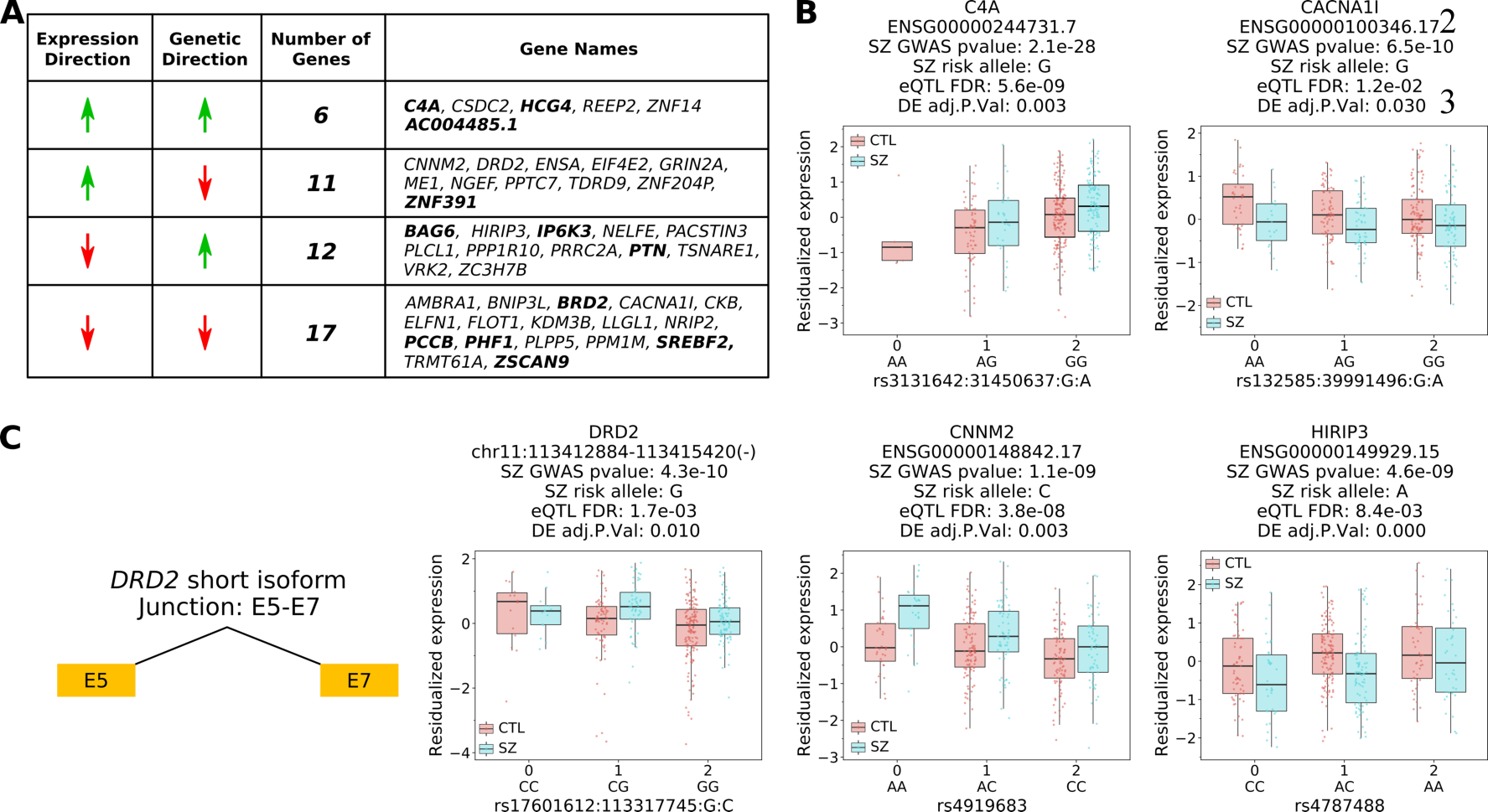
Integration of GWAS, DE, and eQTL identifies 46 genes associated with both disease risk and clinical state. **(A)** Summary of the intersection between GWAS, DE, and eQTL. Genes in bold are also present within TWAS significant (FDR < 0.05) associated genes. **(B)** Four examples of genes associated with differential expression directionality concordant with the direction of schizophrenia risk according to GWAS (top) and discordant with schizophrenia risk (bottom). **(C)** Junction specific to DRD2 short isoform, exon 5 to exon 7, associated with schizophrenia risk.

Additionally, *CACNA1I* has been previously associated with schizophrenia in the Chinese population (*34, 35*) as well as in GWAS (*3*) and rare missense mutations in *CACNA1I* disrupting its activity were found in schizophrenic individuals (*36*). Our data suggest, however, that the therapeutic action targeting CACNA1I would be agonism, rather than antagonism which has previously been explored (*37*).

Interestingly, as noted above, on the junction level integration analysis, we identified the D2S specific junction (5–7) (Fig. 6C) as showing discordant associations with illness risk and illness state, likely because of environmental factors and consequences of the disease and its treatment. Indeed, antipsychotic drugs upregulate DRD2 abundance. Plots and summary information for all genes, transcripts, exons, and junctions in the GWAS/DE/eQTL intersection are summarized in Tables S8-10 and Fig. S16-27. Full information for each feature is available in Data S5.

### Learning a caudate nucleus gene expression network with deep neural networks

To gain novel insights about gene expression relationships in the caudate, we created Gene Networks with Variational Autoencoders (GNVAE, Fig. 7A), a new method based on deep neural networks to learn biological networks from gene expression data. GNVAE is a manifold learning-based method that uses a disentangling variational autoencoder (*38, 39*) to obtain a compressed representation of each gene’s expression pattern into a low-dimension vector of latent variables. By using learned representations of expression patterns to build a gene network, GNVAE focuses on expression modes that are recurrent among genes and tends to capture meaningful biological themes. Autoencoders are neural networks that are trained to reconstruct their inputs at the output layer. By using a low-dimensional bottleneck layer, autoencoders learn a compressed, non-linear representation of the data that usually captures meaningful properties of the data. Disentangling variational autoencoders have a loss function that encourages the latent variables to be statistically independent of each other. In our approach, we train the autoencoder considering each gene as a training example and its expression values across individuals as features. After training the autoencoder, GNVAE uses the learned representation vectors to compute distances between all pairs of genes, forming a distance matrix. At this point, we can use the distance matrix directly to identify neighbors of genes of interest in the representation space. Alternatively, we can identify modules of genes with similar representation. GNVAE computes a neighborhood graph from the distance matrix and applies the Leiden clustering algorithm (*40*) to identify gene modules.

**Fig. 7:**
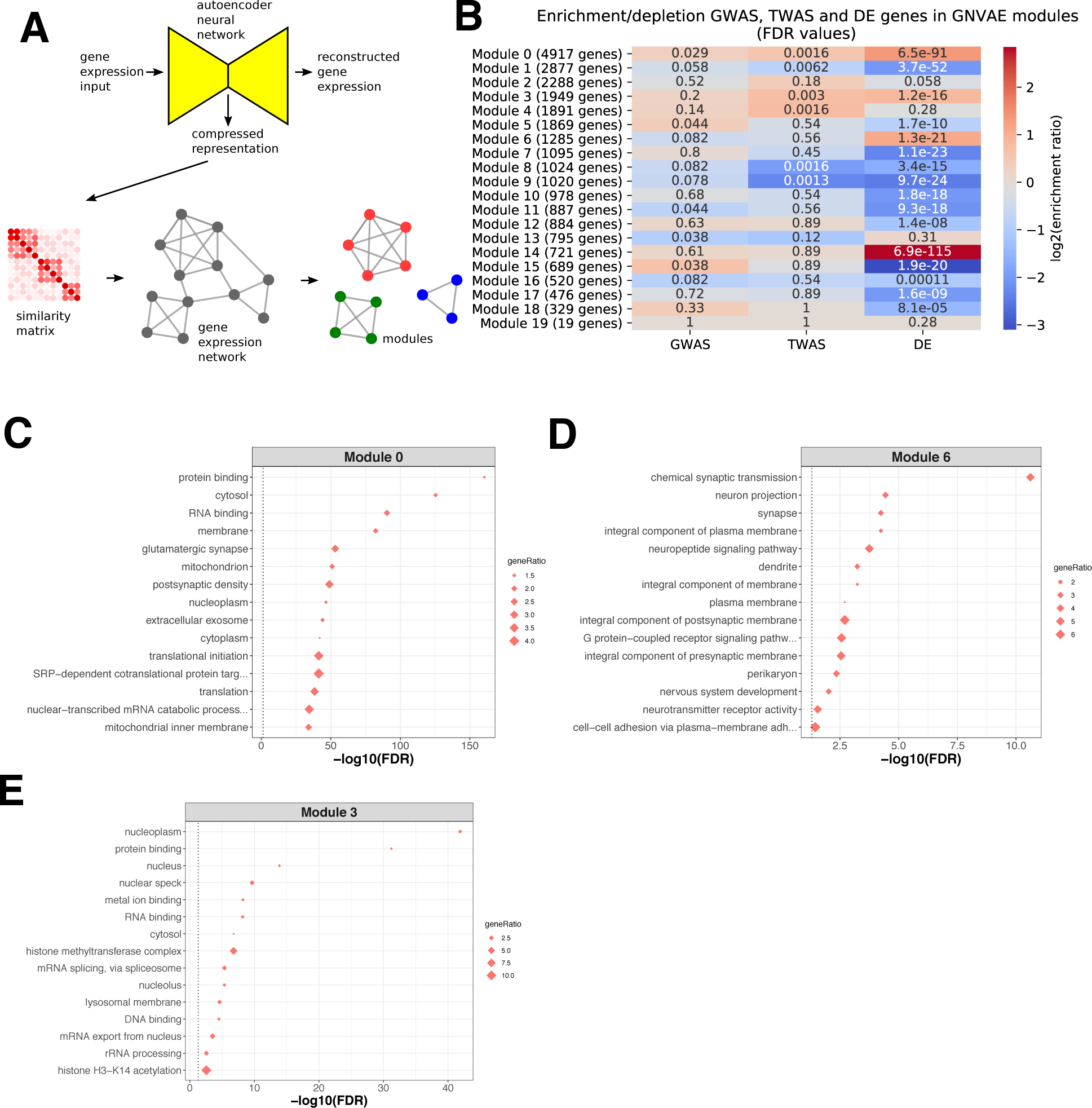
Learning a caudate nucleus gene expression network with deep neural networks. **(A)** Overview of GNVAE pipeline. **(B)** Enrichment analysis (FDR: hypergeometric test p-values corrected for multiple hypotheses testing with Benjamini-Hochberg procedure). **(C)** Top 15 enriched Gene Ontology terms for Module 0, which contains the *DRD2* gene and *DRD2* junctions 5-6 and 6-7. **(D)** Top 15 enriched GO terms for Module 6, which contains *DRD2* junctions 5-7. **(E)** Top 15 enriched GO terms for Module 3, which contains the *SETD1A* gene.

We applied GNVAE to the set of 394 adult schizophrenia and control caudate nucleus samples. We obtained a distance matrix, a neighborhood graph and 20 modules, and for each module we calculated Gene Ontology term enrichment (Fig. 7 and Data S6) and tested if the module is enriched or depleted in genes located in GWAS-significant loci, TWAS-significant genes and differentially expressed genes (Fig 7B). Several modules show highly significant enrichment or depletion for genes differentially expressed in schizophrenia, and some modules show enrichment or depletion for GWAS and TWAS genes, suggesting specific expression patterns are shared among a significant portion of TWAS, GWAS and DE genes. Interestingly, *DRD2* junction 5-7 (from the presynaptic autoreceptor isoform) was attributed to a different module than *DRD2* junctions 5-6 and 6-7 (from the postsynaptic isoform), perhaps reflecting their distinct biological roles and anatomic separation. *DRD2* junctions 5-6 and 6-7 were attributed to module 0 (Fig. 7C), which is enriched in a broad range of GO terms, including translation initiation, protein and RNA binding and terms related to the synapse. Module 0 is significantly enriched in GWAS, TWAS and DE genes. The *DRD2* junction 5-7 (from the short isoform) was attributed to module 6 (Fig. 7D), which is enriched for GO terms related to the synapse, including chemical synaptic transmission and neurotransmitter receptor activity, as well as neuronal projection and axon. Interestingly, this module is strongly enriched for glutamatergic synapse. Module 6 is also significantly enriched in DE genes. Interestingly, *SETD1A*, a histone methyltransferase gene with rare mutations leading to schizophrenia-like phenotypes, was attributed to module 3 (Fig 7E), which is enriched in TWAS and DE genes and is enriched in GO terms including protein binding and histone methyltransferase complex. Of the other three modules that are enriched for TWAS genes, module 4 is enriched for transcription regulation and DNA binding, module 8 is not enriched for any GO terms and module 9 is enriched for miRNA gene silencing (Data S6).

In contrast, when we applied WGCNA (Weighted Gene Co expression Network Analysis) on the same samples, we found primarily significant enrichment for only DEG for schizophrenia across 20 of the 22 modules with only two modules associated with GWAS genes (Fig. S28 and Data S7). Interestingly, the separation of the short and long *DRD2* junctions was replicated in the WGCNA modules. Here, the *DRD2* junctions 5-6 and 6-7 were in the purple module, which did not show any GO enrichment. Unlike with GNVAE, the purple module was only significantly enriched for DE genes. The *DRD2* junction 5-7 was in the cyan module, where GO terms associated with the synapse similar to GNVAE module 6 were enriched (Fig. S29). The cyan module was also only enriched for DE genes.

Collectively, these data suggest that expression representations captured by GNVAE tend to place genes in biologically meaningful neighborhoods, which can provide insight into potential interactions if these genes are targeted for therapeutic intervention. Further, that GNVAE modules show enrichment for both trait and state factors suggests that insights may emerge from this approach that could be missed in traditional WCGNA analyses.

## Discussion

We have profiled the genetic and transcriptional landscapes of the caudate nucleus with respect to schizophrenia in the largest caudate dataset to date. We annotated genetic regulation of gene expression across four genomic features (gene, transcript, exon, and exon-exon junction), finding millions of statistically significant *cis*-eQTLs, and thousands with SNPs associated with schizophrenia risk (GWAS p-value < 5e-8). When we compared these eQTLs with other tissues from mostly the same brains from BrainSeq Phase II (DLPFC and hippocampus), and also GTEx Caudate, and CMC DLPFC, we found both region specific and region non-specific genetically regulated SNP-gene associations. We also find an intrinsic variant to gene expression directionality relationship independent of brain region, such that 99% of the significant eQTLs that are present in multiple regions affect gene expression in the same direction.

To identify significant genetically regulated expression to SZ trait associations, we performed TWAS to integrate gene expression, genetic variation, and schizophrenia risk for caudate nucleus across multiple features. We identify novel candidate genomic features outside of GWAS significant loci that may have causal relationship with schizophrenia, and distinguish region-specific and shared associations across DLPFC, hippocampus, and caudate. For example, *JKAMP* - a JNK1/MAPK8 associated membrane protein - was identified as shared between the three brain regions; moreover, it was also detected in a TWAS of the cerebral cortex (*7*). In contrast, *MIAT*, which has prior connections with schizophrenia but is outside of GWAS significance, is a caudate specific TWAS association. These divergent regional data highlight the importance of a multiple brain region approach in deciphering the underlying mechanisms of schizophrenia risk.

We identified 2699 genes in caudate differentially expressed between patients with schizophrenia and neurotypical controls, which was substantially more than in the previous study of DLPFC and hippocampus largely from the same individuals (245 and 48 DEGs, respectively at FDR < 0.05). Interestingly, the top downregulated gene for caudate was *TH*, which encodes tyrosine hydroxylase, the rate-limiting enzyme in the biosynthesis of dopamine. Given that most of the patients in this study were on antipsychotic drugs at the time of their death, this finding of reduced DA presynaptic synthesis is consistent with evidence from animal studies that chronic treatment with DRD2 antagonist drugs induce a relative inactivation of DA neuronal activity (*41, 42*). Within our GO analysis of caudate, we found significant downregulation of oligodendrocytes, myelination, and axonogenesis, which is observed in morphological studies with a general decrease in the number of glial positive cells (*43, 44*). Recent work has highlighted this pattern of oligodendrocyte deficits also in autism and in other CNS disorders with prominent neuronal involvement (*45*).

Integration of GWAS, eQTL and differential expression analysis identified 46 genes that are both genetically associated with schizophrenia risk and differentially expressed in the caudate nucleus with consistent directionality. These genes uniquely bear association both with risk and the illness state. Among these genes is *CACNA1I*, which encodes a voltage-gated calcium channel playing a role in sleep spindle activity (*46*), and associated with schizophrenia GWAS risk (*4*). Interestingly, the association of genetic risk and illness state with this gene is for decreased expression, suggesting that therapeutic targeting would involve increasing activity of this channel (*37*).

We developed GNVAE, a new approach to learn biological networks from gene expression data using deep neural networks. The gene expression representations captured by GNVAE tend to place genes in biologically meaningful neighborhoods and also reveal modules enriched for both trait and state associated genes, which can be used as a resource to identify potential interactions for genes to be targeted for therapeutic intervention.

### DRD2 and Schizophrenia

The caudate nucleus is rich in DRD2 receptors and has been a focus of studies of the DA system in schizophrenia for decades, using both postmortem analyses and in vivo radioreceptor imaging (*17, 47*). It has generally been assumed that the DA system is overactive and that, in particular, expression of the DRD2 receptor is increased, potentially facilitating increased DA signaling (*17*). However, evidence from our eQTL analysis reveals that decreased expression of specifically the short isoform of dopamine D2 receptor in the caudate is associated with genetic risk for schizophrenia. No such association was found for the long isoform of DRD2. In contrast, differential expression analysis indicates increased expression of D2S in the caudate nucleus of schizophrenic patients, with again no signal in D2L, in agreement with an earlier report that did not have power for statistical significance (*48*). The genetic state of risk may be obscured in patient samples because of environmental factors, including developmental and disease- associated compensatory effects especially antipsychotic medication. DRD2 antagonists have been shown to lead to increased abundance of DRD2 receptors on the cell membrane, presumably because they block internalization of the DRD2 receptor after binding of DA (*17*).

At least in our postmortem human brain samples, this presumed effect is only seen presynaptically. While it is unclear whether and how this presynaptic effect might contribute to the antipsychotic actions of DRD2 antagonists, concomitant blockade of postsynaptic D2L reduces striatal DA signaling, the presumed basis of antipsychotic action and bias to parkinsonian side effects.

A question that arises is whether the association with the DRD2 short isoform is based specifically on expression of this isoform on dopaminergic neurons that project into the caudate nucleus or whether it is based on expression of this isoform on intrinsic caudate cells or perhaps even on other inputs, such as cortical glutamatergic terminals. A recent study has found evidence of ubiquitous local protein synthesis in presynaptic terminals (*49*), which could implicate the presence of D2 short mRNA in the caudate nucleus having originated in dopaminergic neurons projecting from the nigra.

In summary, we provide a comprehensive genetic and transcriptional analysis of the caudate nucleus with respect to schizophrenia, with indications of novel genetic associations and potential therapeutic targets. We identify a potential mechanism of the DA link with schizophrenia involving presynaptic autoreceptor regulation of DA release, suggesting that psychosis risk involves relatively compromised regulation of release, which in the presence of events that lead to increased DA neuronal activity, would bias towards increased synaptic DA. It is tempting to speculate that individuals so genetically affected under stress, when DA activity is increased, fail to appropriately modulate this activity at the synapse and are susceptible to sustained increased DA signaling when the context is no longer appropriate to reinforce stimuli converging on striatal neurons. We further speculate that the development of drugs targeting select presynaptic components of the dopamine autoregulation system might open new avenues in the treatment of psychosis.

## Materials and Methods

### Human postmortem brain tissue acquisition

Human postmortem brain tissue was collected at several sites for this study. A large number of samples were obtained at the Clinical Brain Disorders Branch (CBDB) at National Institute of Mental Health (NIMH) from the Northern Virginia and District of Columbia Medical Examiners’ Office, according to NIH Institutional Review Board guidelines (Protocol #90-M- 0142). These samples were transferred to the Lieber Institute for Brain Development (LIBD) under an MTA with the NIMH. Additional samples were collected at the LIBD according to a protocol approved by the Institutional Review Board of the State of Maryland Department of Health and Mental Hygiene (#12-24) and the Western Institutional Review Board (#20111080). Additional fetal, child, and adolescent brain tissue samples were provided by the National Institute of Child Health and Human Development Brain and Tissue Bank for Developmental Disorders via Material Transfer Agreements (NO1-HD-4-3368 and NO1-HD-4-3383) approved by Institutional Review Board of the University of Maryland.

Audiotaped informed consent to study brain tissue was obtained from the legal next-of-kin on every case collected at NIMH and LIBD. Details of the donation process and specimens handling are described previously (*50*). After next-of-kin provided audiotaped informed consent to brain donation, a standardized 36-item telephone screening interview was conducted, (the Lieber Institute for Brain Development Autopsy Questionnaire), to gather additional demographic, clinical, psychiatric history, substance abuse history, treatment, medical, and social history. A psychiatric narrative summary was written for every donor, to include data from multiple sources, including the Autopsy Questionnaire, medical examiner documents (investigative reports, autopsy reports, and toxicology testing), macroscopic and microscopic neuropathological examinations of the brain, as well as extensive psychiatric, detoxification, and medical record reviews, and/or supplemental family informant interviews using the MINI (Mini International Neuropsychiatric Interview). Two board-certified psychiatrists independently reviewed every case to arrive at DSM-5 lifetime psychiatric and substance use disorder diagnoses, including [schizophrenia and bipolar disorder, as well as substance abuse disorders], and if for any reason agreement was not reached between the two reviewers, a third board-certified psychiatrist was consulted.

All donors were free from significant neuropathology, including cerebrovascular accidents and neurodegenerative diseases. Each subject was diagnosed retrospectively by two board-certified psychiatrists, according to the criteria in the DSM-IV. Brain specimens from the CBDB were transferred from the NIMH to the LIBD under a Material Transfer Agreement. Available postmortem samples were selected based on RNA quality (RNA integrity number ≥ 5).

The toxicological analysis was performed in each case. The non-psychiatric non-neurological controls had no known history of significant psychiatric or neurological illnesses, including substance abuse. Positive toxicology was exclusionary for control subjects but not for patients with psychiatric disorders.

### Subject details

In total, 444 caudate postmortem brain samples were used in this study. The demographic data are summarized in Table S11. In brief, the caudate cohort contains 154 subjects with schizophrenia, 44 subjects with bipolar disorder and 246 non-psychiatric controls. Data S8 includes individual level demographic information.

### Human postmortem brain processing and dissections

The caudate nucleus was dissected out, pulverized, and stored at -80 °C. Briefly, after removal from the calvarium brains examined, photographed, weighed, and then the brainstem and cerebellum were removed via transection just above the quadrigeminal plate. The circle of Willis was dissected from the ventral surface of the brain, and the pineal gland was removed. The hemispheres were separated along the midline, and then each hemisphere was cut into approximately 1 cm thick coronal slabs from the frontal pole to the occipital pole. The cerebellar hemispheres were sectioned along the midline through the vermis, and then each hemisphere was cut horizontally into two equals blocks. The brainstem was sectioned into two midbrain blocks, two pontine blocks, two medullary blocks, and one block of the upper cervical spine, cut perpendicularly to the long axis of the brainstem. Slabs and blocks were flash frozen in a slurry of dry ice and isopentane, and then stored in zip lock bags inside labeled cardboard boxes at -80 °C until retrieval for caudate dissection.

The caudate nucleus was dissected from the slab containing the caudate and putamen at the level of the nucleus accumbens. The caudate was dissected from the dorsal third of the caudate nucleus, lateral to the lateral ventricle, to make certain that the caudate dissections did not impinge upon the nucleus accumbens. Dissections were done under visual guidance using a hand-held dental drill, on a tray over dry ice. Approximately 250 mg of caudate were moved per subject before pulverization. Tissue was kept frozen at all times throughout the brain dissection and pulverization steps.

### Genotype data processing

Genotype data processing was performed as previously described (*8*). Briefly, genotyping with Illumina BeadChips was carried out using DNA extracted from cerebellar tissue according to the manufacturer’s instructions. Genotype data was processed and normalized with crlmm (*59*), a R/Bioconductor package, separately by platform. Genotype prephasing and imputation was conducted separately by platform on observed genotypes with low quality and rare variants removed using IMPUTE2 (v2.3.2) (*51*) and ShapeIT v2 (v2.r837) (*52*) with imputation reference set from the full 1000 Human Genomes Project Phase 3 dataset (*53*) using genome build hg19.

We retained common SNPs (MAF > 5%) that were present in the majority of samples (missingness < 10%) that were in Hardy Weinberg equilibrium (at p > 1x10^-6^) using Plink (v1.90) (*54*). We then identified linkage disequilibrium (LD)-independent SNPs to use for population stratification of samples with multidimensional scaling (MDS). The first component separated samples by ethnicity. Variants were mapped from v142 dbSNP database (*55*) on hg19 to v149 on hg38, and liftOver (*56*) utilized to map chromosome locations. This processing and quality control steps resulted in 6,882,303 common variants in this dataset of 487 individuals, which was used for downstream analysis.

### RNA sequencing

Samples were sequenced as previously described (*6*). Briefly, sequencing libraries were prepared from 300 ng of total RNA using the TruSeq Stranded Total RNA Library Preparation kit with Ribo-Zero Gold ribosomal RNA depletion. For quality control, synthetic External RNA Controls Consortium (ERCC) RNA Mix 1 was spiked into each sample. These paired-end, strand-specific libraries were sequenced on an Illumina HiSeq 3000 at the LIBD Sequencing Facility. We generated FASTQ files using the Illumina Real Time Analysis module by performing image analysis, base calling, and the BCL Converter (CASAVA v1.8.2). The reads were aligned to the hg38/GRCh38 human genome (GENCODE release 25, GRCh38.p7) using HISAT2 (v2.0.4) (*57*) and Salmon (v0.7.2) (*58*) using the reference transcriptome to initially guide alignment based on annotated transcripts. The synthetic ERCC transcripts were quantified with Kallisto (v0.43.0) (*59*).

### RNA data processing

Counts were generated as previously described (*6*). Briefly, sorted BAM files from HISAT2 alignments were generated and indexed using SAMtools (v1.6; HTSlib v1.6). Alignment quality was assessed using RSeQC (v2.6.4) (*60*). The transcriptomes were characterized using four genomic features: 1) genes, 2) exons, 3) transcripts, and 4) exon-exon junctions. For transcripts, estimated counts were extracted for Salmon files for downstream differential expression analysis.

1. We generated gene counts using the SubRead utility featureCounts (v1.5.0-p3) (*61*) for paired end, reversed stranded read counting.
2. We also generated exon counts using featureCounts for paired end, reversed stranded read counting.
3. We generated transcript counts and TPM estimates using Salmon.
4. We extracted exon-exon splice junctions from BAM files filtered for primary alignments using regtools (v0.1.0) (*62*) and bed_to_juncs script from TopHat2 (*63*).

### Quality control and sample selection

Quality control of samples was determined as previously described (*6*). Briefly, samples were checked for four quality control measures: 1) ERCC concentrations, 2) genome alignment rate (>70%), 3) gene assignment rate (> 20%), and 4) mitochondrial mapping rate (< 6%). N samples were dropped for poor quality control based on the above measures resulting in 464 samples after quality control. Next, we select samples for age (>13) for a final number of 444 samples.

### Degradation data generation

The qSVA algorithm uses data from a separate RNA-Seq assay measuring RNA degradation in brain tissue (*32*). Aliquots of 100 mg pulverized caudate nucleus tissue from 5 individuals were left on dry ice and placed at room temperature until reaching the respective time interval, at which point the tissue was placed back onto dry ice. The four time intervals tested were 0, 15, 30, and 60 min, with the 0-minute aliquot remaining on dry ice for the entirety of the experiment. RNA extraction began immediately after the end of the final time interval, and RiboZero RNA-Seq libraries were prepared for each time point and each individual. From the RNA-Seq data, the set of 1000 expressed regions (*64*) most affected by RNA degradation was determined. Then the expression at these 1000 regions for the caudate samples was calculated to form the caudate nucleus degradation matrix, from which the top 12 principal components (PCs) are selected using the BE algorithm (*65*) while considering diagnosis status, age at time of death, sex, mitochondrial mapping rate, rRNA mapping rate, total assigned reads to gene proportion, and the first five ancestry PCs. These 12 PCs are referred to as quality surrogate variables (qSVs) and used as adjustment variables in differential expression analysis.

### Expression normalization

To normalize expression for each genomic feature, we first filtered out low expressing counts via filterByExpr from edgeR R/Bioconductor package (*66, 67*). Following filtering, we normalized counts for RNA composition using TMM an edgeR utility. For differential expression analysis, we accounted for sample variation by fitting a model across each of the genetic features as a function of SZ diagnosis adjusting for age, sex, ancestry (SNP PCs 1-5), and RNA quality (RIN – RNA Integrity Number, mitochondria mapping rate, gene assignment rate, genome mapping rate, rRNA mapping rate, ERCC error rate, and qSVA (*32*)), followed by applying the utility voom from the limma R/Bioconductor package (*68, 69*).

### eQTL expression feature-level filtering

We filtered lowly expressed features for eQTL analysis based on BrainSeq Phase II (PMID 31174959). Briefly, we used cutoffs for mean expression across all 444 samples: genes (RPKM > 0.2), transcripts (TPM > 0.4), exons (RPKM > 0.2), and junctions (RP10M > 0.4).

### Identification of cis-eQTLs

*Cis*-eQTL mapping was performed as previously described (*6*). Briefly, we identified eQTLs within caudate nucleus using Matrix eQTL (*70*) for all samples age > 13 with log2 transformed RPKM for genes and exons, RP10M for exon-exon junctions, and TPM for transcripts adjusted for diagnosis, sex, SNP PCs (1–5), and expression PCs (Equation 1) determined via the num.sv function from sva R/Bioconductor package (*71*) with vfilter parameter set to 50,000. The mapping window parameter was set to 0.5 Mb up- and downstream gene body. False discovery rate was calculated within the program with noFDRsaveMemory set to false using linear mode, modelLINEAR, and empty numeric vector for error covariance.

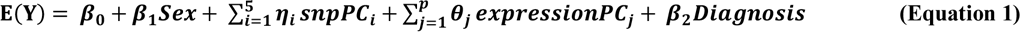

To identify eQTLs that have different effects by brain region, we ran a similar pairwise analysis but limiting diagnosis to control and schizophrenia diagnosis and using modelLINEAR_CROSS mode (Equation 2).

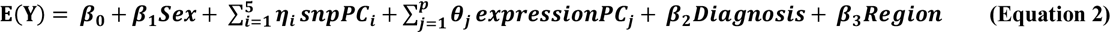

### Schizophrenia GWAS risk SNPs

We downloaded the list of index SNPs and meta-analysis of high-quality imputed SNPs determined by the Psychiatric Genomics Consortium (CLOZUK+PGC2) (Pardiñas 2018). From these lists we identified the CLOZUK+PGC2 SNPs in the BrainSeq SNPs. To this end, we converted the PGC2 GWAS SNPs from hg19 to hg38 using pyliftover. Following conversion, we merged our SNPs with the PGC2 GWAS SNPs on hg38 coordinates and matched alleles.

### Differential expression analysis

We used eBayes function from limma to identify differentially expressed features from voom normalized counts. We adjust for: age, sex, ancestry (top 5 genotype principal components) and several RNA-Seq sample quality measures: fraction of reads mapping to the genome, fraction of reads mapping to mitochondria, fraction of reads mapping to ribosomal RNA, fraction of reads assigned to genes, RNA integrity number (RIN), total ERCC deviation from expected counts and top 12 quality surrogate variables (qSVs, to account for RNA degradation (*32*)), using the model described in Equation 3. The number of qSVs, K=12 for the caudate dataset, was calculated using the BE algorithm (*65*) implemented in the SVA Bioconductor package. For comparison with the CommonMind Consortium dataset, we downloaded open access differential expression summary results and matched by gene IDs.

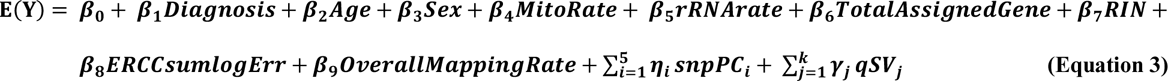

### Gene term enrichment and pathway analyses

For differential expression gene term enrichment analysis, we utilized enrichment analysis (enrichGO), gene set enrichment analysis (gseaGO), and directional comparison enrichment (compareCluster) with Entrez IDs for genes, transcripts, exons, and exon-exon junctions using functions from clusterProfiler (*72*), a R/Bioconductor package. In addition to gene term enrichment analysis, we also conducted pathway analysis for differential expression results using pathview (*73*), a R/Bioconductor package. Parameters for all functions can be found within the corresponding jupyter notebooks (see section **Data and Code Availability** for details).

### Venn diagrams and upset plots

We generated venn diagrams using python venn package for unweighted four feature overlaps and matplotlib-venn package for weighted three tissue overlaps. Upset plots were generated using the R UpSetR package.

### Expression residualization

We generated residualized data using voom normalized counts and a modified version of the residuals function from limma. To this end, we created a null model, Equation 4, without variable of interest (e.g., diagnosis), fit the null model using lmFit from limma, and regressed out covariates using the fitted model coefficients. Following residualization, we transformed the data with a z-score standardization. All boxplots used residualized expression.

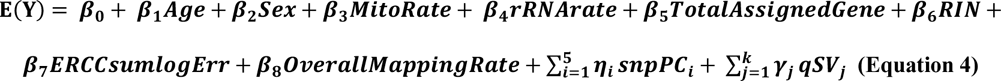

### TWAS analysis

For transcriptome-wide association study analysis, we first adapted the LD reference files provided by the FUSION TWAS software (*25*) and the GWAS summary statistics SNPs from PGC2 and the Walters Group Data Repository (*3*) from hg19 to hg38 using the port_to_hg38.R script (https://github.com/LieberInstitute/brainseq_phase2/tree/master/twas (*6*)) modified to perform LD and summary statistics conversion separately. Following conversion, we computed feature weights using the example script provided by the FUSION TWAS software modified to run in parallel with our data and FUSION.compute_weights.R (FUSION TWAS software; gemma v0.98.1) with slight modifications to run with multiple threads and gcta v1.92beta.

Summary information for the feature weights were generated using FUSION.profile_wgt.R (FUSION TWAS software) and a python script was used to extract weight positions for downstream analysis. After computing functional weights, we applied FUSION.assoc_test.R and FUSION.post_process.R to generate TWAS association and calculate functional GWAS associations. The TWAS p-values were adjusted for multiple testing using the Benjamini-Hochberg procedure implemented in the statsmodels Python package.

### CommonMind Consortium and Genotype-Tissue Expression replication

For CommonMind DE and eQTL replication, we download differential expression and eQTL result from Synapse (https://www.synapse.org/), syn6183936 and syn4622659. For eQTL replication we used caudate (brain caudate basal ganglia) from GTEx v7, which is supported by the Common Fund of the Office of the Director of the National Institutes of Health, and by NCI, NHGRI, NHLBI, NIDA, NIMH, and NINDS. We obtain eQTL data from the GTEx Portal (https://gtexportal.org/home/datasets). For SNP-gene comparisons of eQTLs, we matched converted SNP IDs across datasets.

### GTEx dopamine receptor D2 cis-eQTL analysis replication

For dopamine receptor D2 (DRD2) eQTL analysis replication, we utilized GTEx v8, extracted SNPs from chromosome 11 of the whole genome sequencing VCF using plink, and subset for the brain caudate basal ganglia. *Cis*-eQTL analysis was performed using Matrix eQTL as described above (**cis-eQTL analysis**) with expression adjusted for GTEx covariates (PCR, platform, sex, SNP PCs (1–5), and PEER (*74*) inferred covariates). Significant *DRD2* eQTLs were determined using p-value cutoff determined via BrainSeq caudate Matrix eQTL analysis at FDR < 0.05.

### Subsampling analysis for comparison of *DRD2* junction 5-7 eQTLs with GTEx

For subsampling of the BrainSeq consortium caudate nucleus eQTL analysis, we randomly sampled 194 individuals from the 444 caudate nucleus samples and performed eQTL analysis (Equation 1) for *DRD2*genomic features (gene, transcripts, exons, and junctions). We performed this 10 times using random sampling of the 444 individuals. To determine significant eQTL associates for GWAS index SNP (rs2514218) and DRD2 junction 5-7, we used the nominal p-value associated with significant eQTL (FDR < 0.05) from the full caudate nucleus (n = 444). As we used a subset of genes, we mapped nominal p-values from the permutation back to the full eQTL nominal p-values to determine FDR.

### Learning a caudate nucleus gene expression network with deep neural networks

We adapted the disentangling autoencoder code from https://github.com/YannDubs/disentangling-vae, which was originally designed for image datasets, to tabular form (for gene expression data) by replacing the convolutional layers with fully connected layers. We used a neural network architecture with 394, 128, 8, 128, 394 neurons in each layer, respectively, with dimension 8 in the bottleneck layer. We use the caudate nucleus gene expression matrix expressed in log2 RPKM. For autoencoder training, we consider each gene as a training example in which the features are the expression values across our cohort of individuals. We perform 10-fold cross validation to verify that reconstruction error in training set and in test set have similar values, indicating there is no overfitting (Fig. S30). We then retrain the autoencoder with the full dataset and apply to each gene to obtain their representation vectors.

We compute a similarity matrix based on the Euclidean distance between the representations of genes, using as similarity score the inverse of squared Euclidean distance. Using the similarity scores, we compute k-neighborhood graph (with k=8) and apply the Leiden clustering algorithm (*40*) to identify modules. For each module, we perform Gene Ontology enrichment analysis with the GOATOOLS Python package (*75*) using hypergeometric tests. We use the enriched GO terms (FDR < 0.05) to generate word clouds using the wordcloud Python package (https://github.com/amueller/word_cloud), using font size proportional to -log(p-value).

### WGCNA analysis

To compare GNVAE with traditional network analysis, we performed signed network WGCNA analysis using the caudate nucleus gene expression matrix expressed in log2 CPM to generate the co-expression network with control and schizophrenia samples. The co-expression network was made using Pearson correlation values with 391 samples and 22972 genes. The Scale-Free Topology and connectivity were evaluated as shown in Fig. S31.

### Data and code availability

Raw and processed data are available from http://erwinpaquolalab.libd.org/caudate_eqtl/. Code and jupyter notebooks are available through GitHub at https://github.com/paquolalab/LieberInstituteBrainSeqPhase3CaudateSchizophrenia.

### Additional resources

Similar to the BrainSeq Phase II release (*6*), we created an eQTL browser available at (http://erwinpaquolalab.libd.org/caudate_eqtl/) that enables exploring the eQTL SNP-feature pairs for caudate nucleus and brain region dependent results comparing the caudate with DLPFC and hippocampus.

## Data Availability

Raw and processed data are available from http://erwinpaquolalab.libd.org/caudate_eqtl/. Code
and jupyter notebooks are available through GitHub at
https://github.com/paquolalab/LieberInstituteBrainSeqPhase3CaudateSchizophrenia

https://github.com/paquolalab/LieberInstituteBrainSeqPhase3CaudateSchizophrenia

## Acknowledgments

The authors would like to extend their appreciation to the Offices of the Chief Medical Examiner of Washington DC, Northern Virginia, and Maryland for the provision of brain tissue used in this study. Appreciation is also extended to Dr. Llewellyn B. Bigelow and members of the LIBD Neuropathology Section for their work in assembling and curating the clinical and demographic information and organizing the Human Brain Tissue Repository of the Lieber Institute. Finally, the authors gratefully acknowledge the families that have donated this tissue to advance our understanding of psychiatric disorders. This work is supported by the Lieber Institute for Brain Development, the BrainSeq Consortium, the National Institutes of Health (NIH) T32 fellowship (T32MH015330) to K.J.M.B, and a NARSAD Young Investigator Grant from the Brain & Behavior Research Foundation to J.A.E.

## Author contributions

Conceptualization, K.J.M.B., the BrainSeq Consortium, J.A.E., D.R.W. and A.C.M.P; Methodology, K.J.M.B., J.A.E., A.E.J., D.R.W. and A.C.M.P.; Software, K.J.M.B., A.S.F., A.R.B., A.E.J., L.C.T., E.E.B., W.S.U. and A.C.M.P.; Formal Analysis, K.J.M.B, A.S.F., A.R.B., and A.C.M.P.; Investigation, J.H.S., R.T., and T.M.H.; Data Curation, A.D-S. and J.E.K.; Writing – Original Draft, K.J.M.B., J.A.E., D.R.W and A.C.M.P.; Writing – Review & Editing, K.J.M.B., A.E.J., L.C.T, T.M.H., J.A.E., D.R.W. and A.C.M.P.; Visualization, K.J.M.B., A.S.F., W.S.U., and A.C.M.P.; Supervision, J.A.E., D.R.W. and A.C.M.P.; Project Administration, the BrainSeq Consortium; Funding Acquisition, the BrainSeq Consortium, J.A.E., and D.R.W.

## Supplementary Materials

BrainSeq Consortium Members

Figures S1-S31

Tables S1-S10

Data Files S1-S8

## Supplementary Information

### BrainSeq Consortium Members

Members of the BrainSeq Consortium include Mitsuyuki Matsumoto, Takeshi Saito, Katsunori Tajinda, Daniel J. Hoeppner, David A. Collier, Karim Malki, Bradley B. Miller, Maura Furey, Derrek Hibar, Hartmuth Kolb, Michael Didriksen, Lasse Folkersen, Tony Kam-Thong, Dheeraj Malhotra, Joo Heon Shin, Andrew E. Jaffe, Rujuta Narurkar, Richard E. Straub, Amy Deep-Soboslay, Thomas M. Hyde, Joel E. Kleinman, and Daniel R. Weinberger.

### Supplementary Data

**Data S1.** eqtl_tables.tar.gz Compressed tar file with R variables containing eQTL results all features (genes, transcripts, exons, and exon-exon junctions) at FDR < 0.05, for caudate and for brain region/genotype interactions.

**Data S2.** BrainSeq_Phase3_Caudate_TWAS_associations_allFeatures.txt.gz Compressed text file of TWAS results for the caudate nucleus with the latest schizophrenia GWAS (PGC2 + CLOZUK) across all features.

**Data S3.** TWAS_gene_tissue_summary.csv Text file of TWAS genes summarized across caudate nucleus, DLPFC, and hippocampus.

**Data S4.** BrainSeq_Phase3_Caudate_DifferentialExpression_DxSZ_all.txt.gz Compressed text file of differential expression results for the caudate nucleus of schizophrenia versus neurotypical controls across all features.

**Data S5.** BrainSeq_Phase3_Caudate_GWAS_DE_eQTL_Integration.txt.gz Compressed text file of integrative analysis at GWAS p-value < 5e-8, DE FDR < 0.05, and eQTL FDR < 0.05 for the caudate nucleus across all features.

**Data S6.** GNVAE_output.tar.gz Compressed tar file containing GNVAE output: table of latent variables for all genes, module membership tables, GO enrichment analysis tables and GO word clouds.

**Data S7.** WGCNA_output.tar.gz Compressed tar file containing WGCNA output: eigengenes, module membership tables, and GO enrichment analysis tables.

**Data S8.** caudate_phenotypes.csv Text file of demographic information for caudate nucleus samples.

**Fig. S1.**
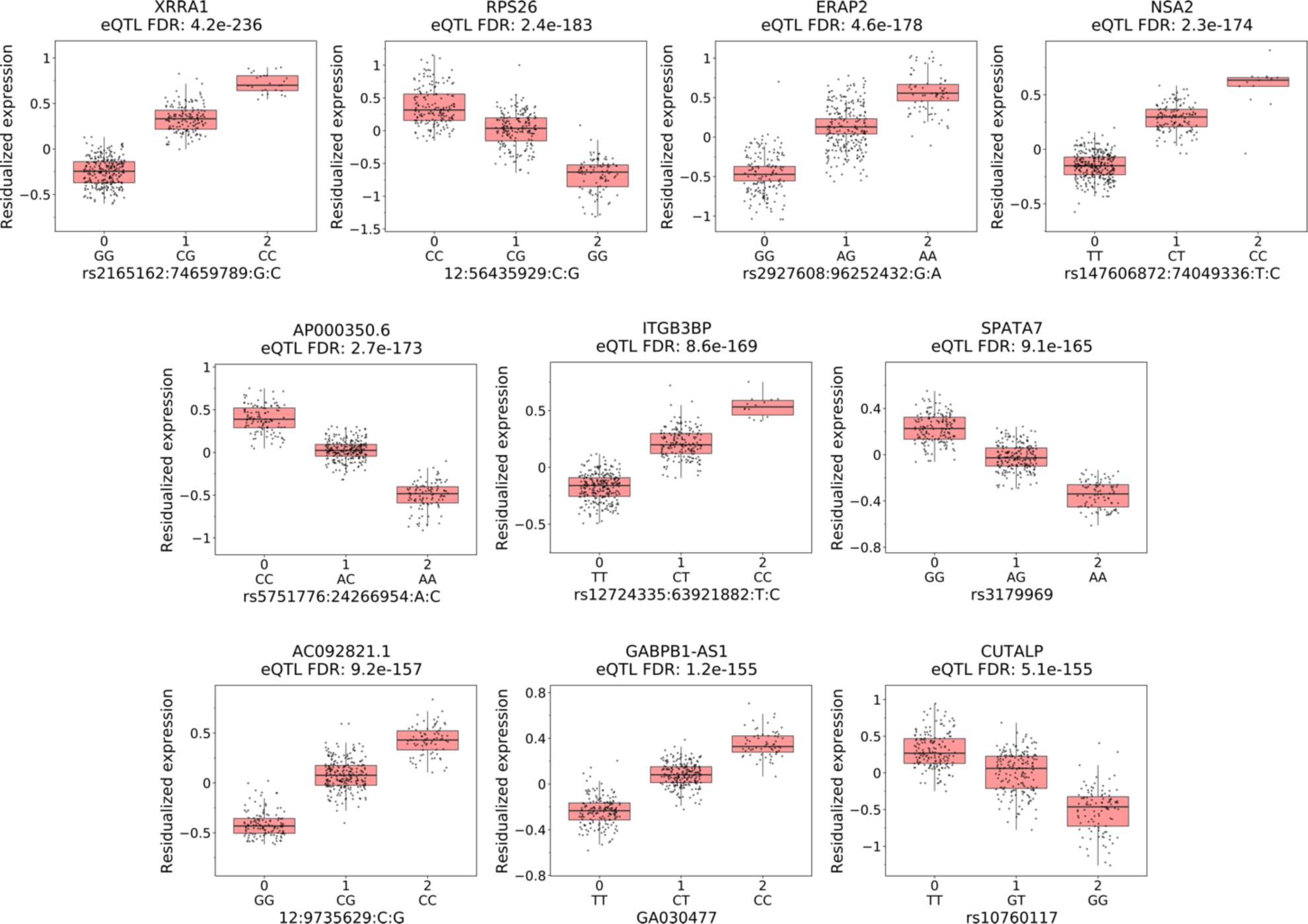
Example boxplots of the ten most significant by FDR corrected p-value *cis*-eQTL associated with unique genes (eGenes) showing changes in the caudate nucleus residualized (y-axis) gene expression with SNP genotypes (x-axis). Plots organized by most significant (FDR) top left. Gene names and FDR p-values annotated for each boxplot.

**Fig. S2.**
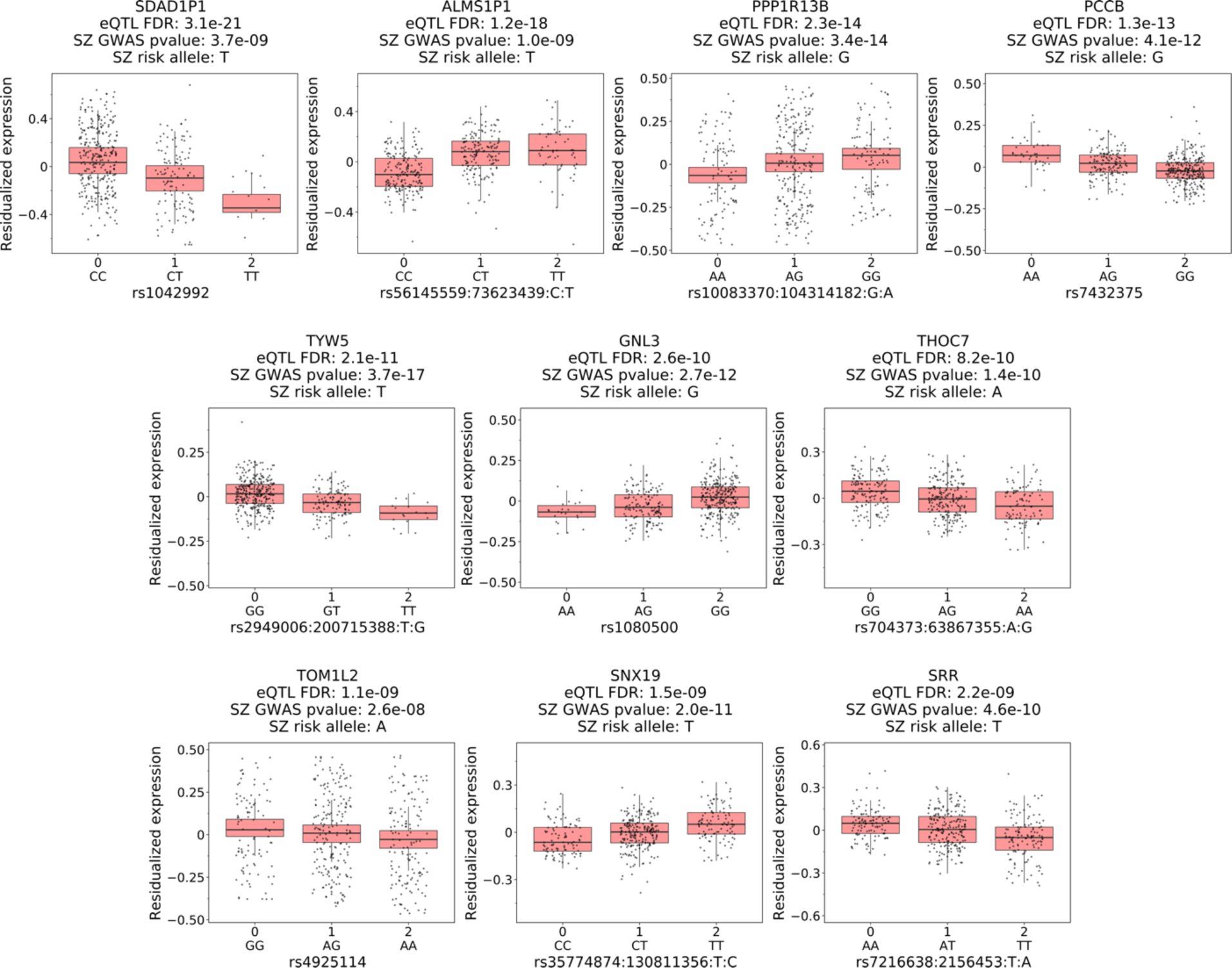
Boxplots of the ten most significant by FDR corrected p-value *cis*-eQTL with index SNPs associated with schizophrenia (SZ) risk for unique genes (eGenes) in the caudate nucleus. Boxplots organized by most significant eQTL FDR show changes in residualized (y-axis) gene expression with SNP genotypes (x-axis). Genotypes organized with increase in schizophrenia risk (left to right). Schizophrenia risk alleles and p-value determined from PGC2+CLOZUK schizophrenia GWAS. Plots organized by most significant (FDR) top left. Gene names and FDR p-values annotated for each boxplot.

**Fig. S3.**
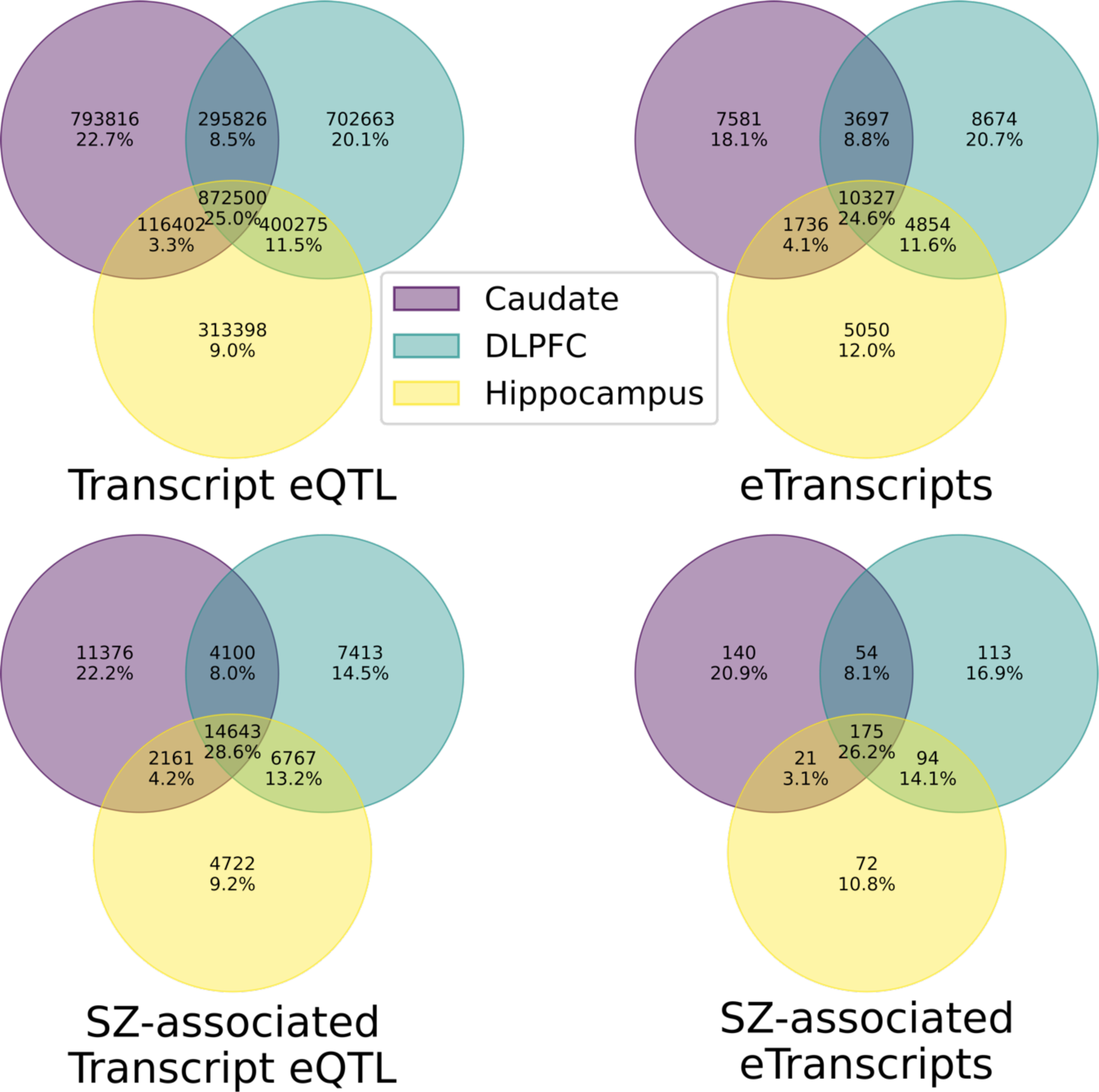
More than 40% of transcript eQTL and eQTL associated with unique transcripts (eTranscripts) including those associated with schizophrenia (SZ) risk are shared with at least two brain regions. Venn diagrams showing percent and number of overlapping significant (FDR < 0.01) transcript eQTL and eTranscripts (top) and those with SNPs associated with schizophrenia risk (PCG2+CLOZUK GWAS p-value < 5e-8; bottom) across the caudate nucleus, DLPFC, and hippocampus.

**Fig. S4.**
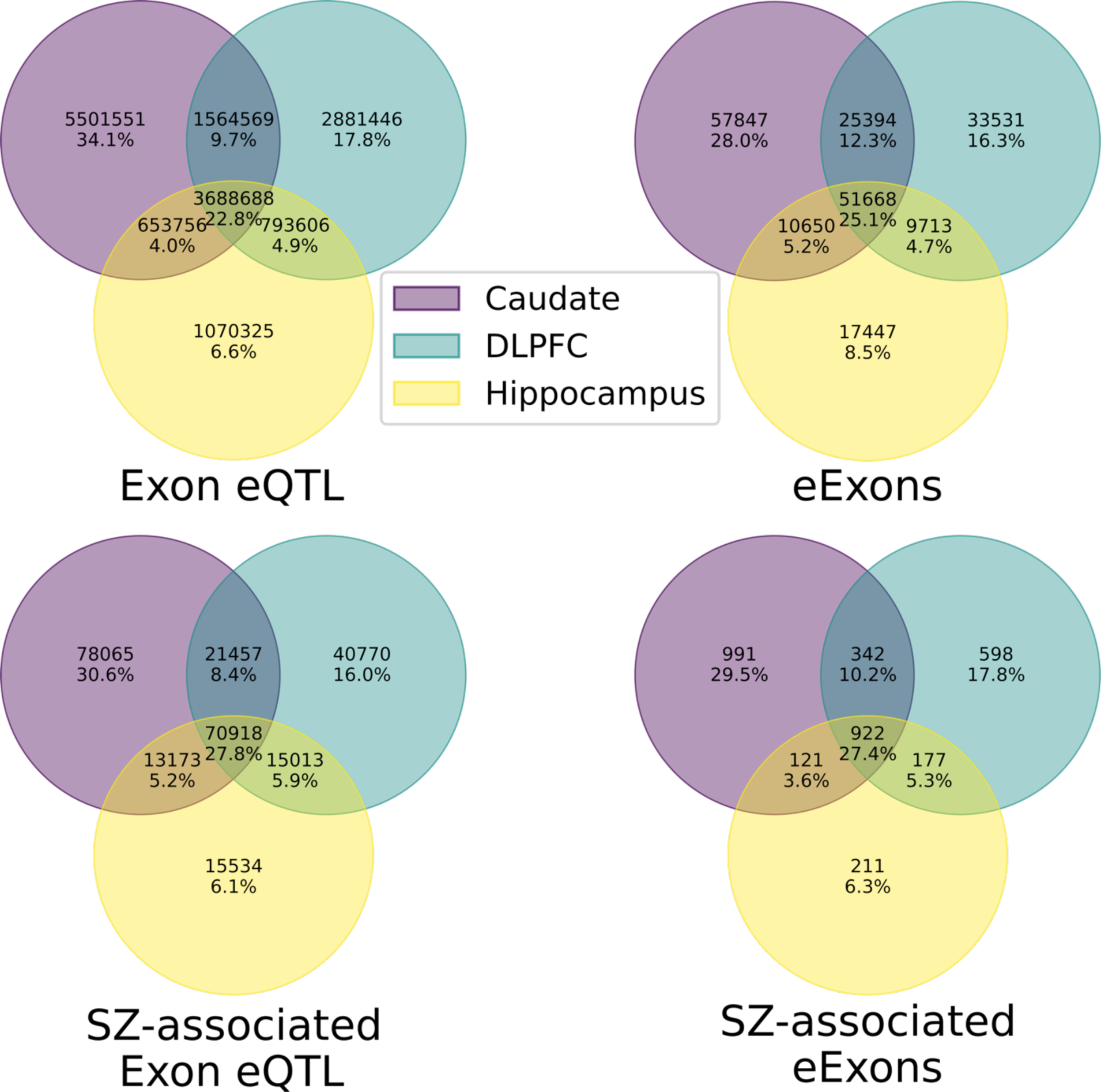
More than 40% of exon eQTL and eQTL associated with unique exons (eExons) including those associated with schizophrenia (SZ) risk are shared with at least two brain regions. Venn diagrams showing percent and number of overlapping significant (FDR < 0.01) exon eQTL and eExons (top) and those with SNPs associated with schizophrenia risk (PCG2+CLOZUK GWAS p-value < 5e-8; bottom) across the caudate nucleus, DLPFC, and hippocampus.

**Fig. S5.**
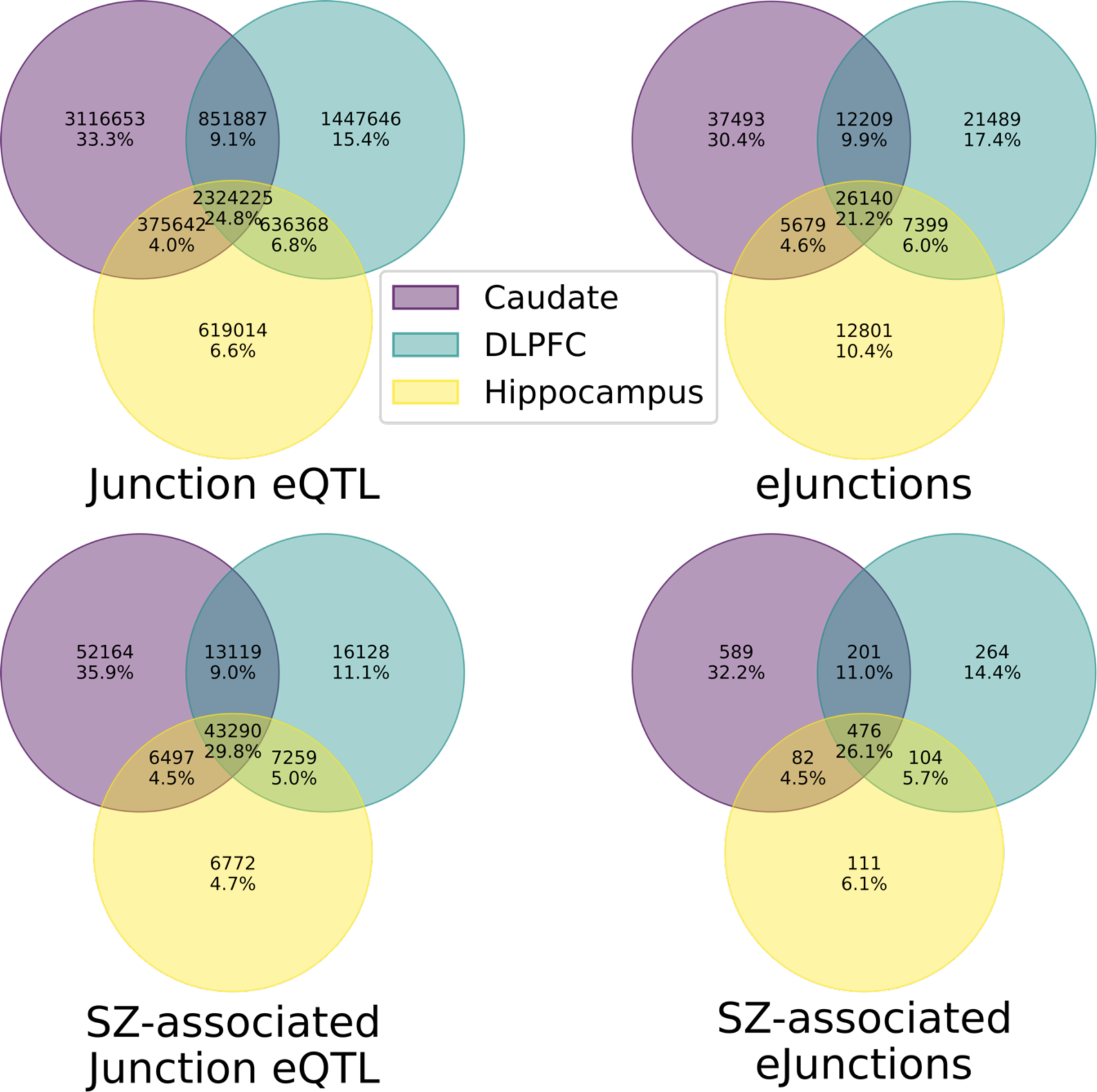
More than 40% of junction eQTL and eQTL associated with unique junctions (eJunctions) including those associated with schizophrenia (SZ) risk are shared with at least two brain regions. Venn diagrams showing percent and number of overlapping significant (FDR < 0.01) junction eQTL and eJunctions (top) and those with SNPs associated with schizophrenia risk (PCG2+CLOZUK GWAS p-value < 5e-8; bottom) across the caudate nucleus, DLPFC, and hippocampus.

**Fig. S6.**
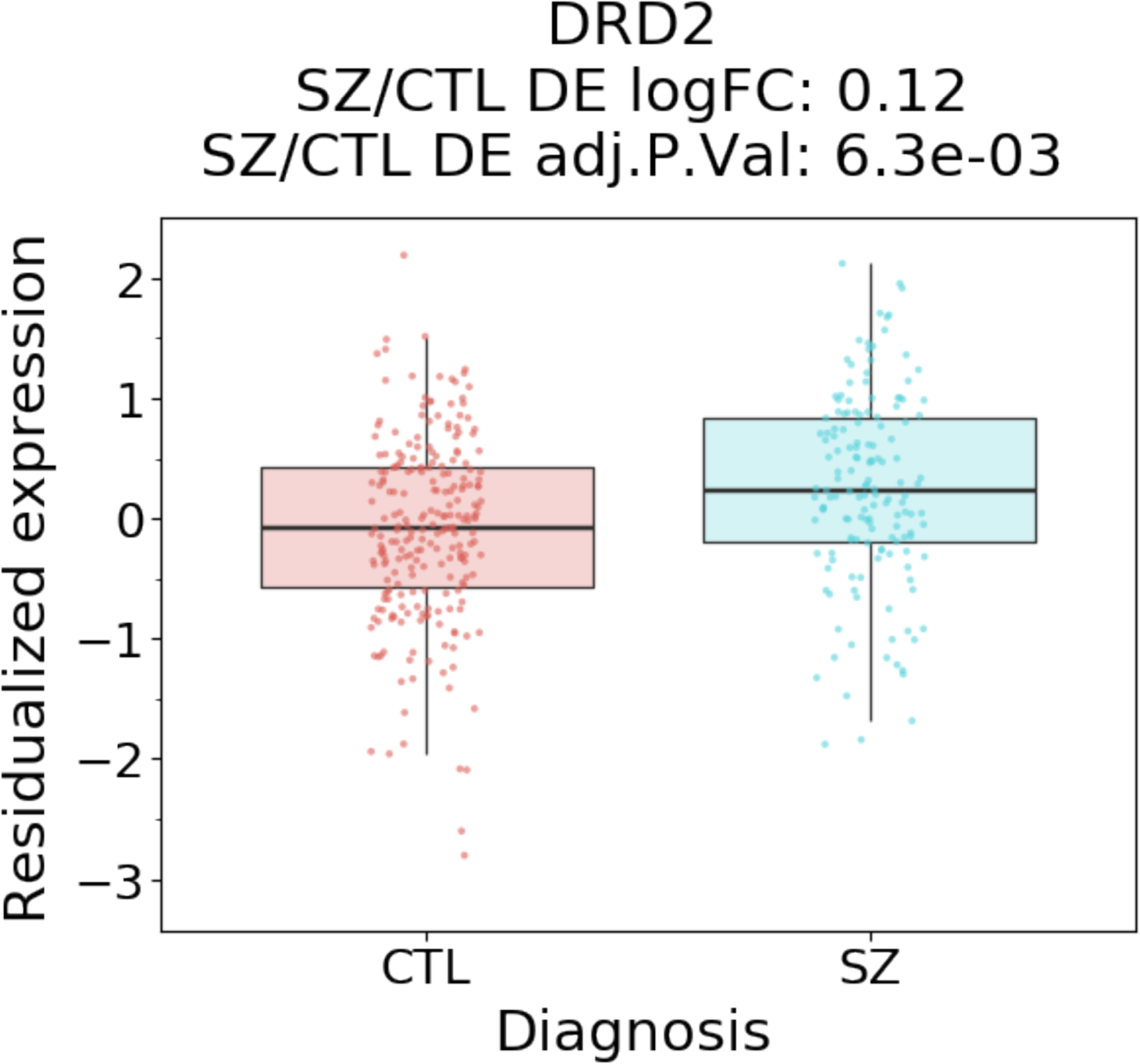
Dopamine receptor D2 (DRD2) significantly upregulated in schizophrenia (SZ) compared to neurotypical controls (CTL) in the caudate nucleus. Boxplot of DRD2 residualized gene expression (y-axis) comparing diagnosis (x-axis) in the caudate nucleus.

**Fig. S7.**
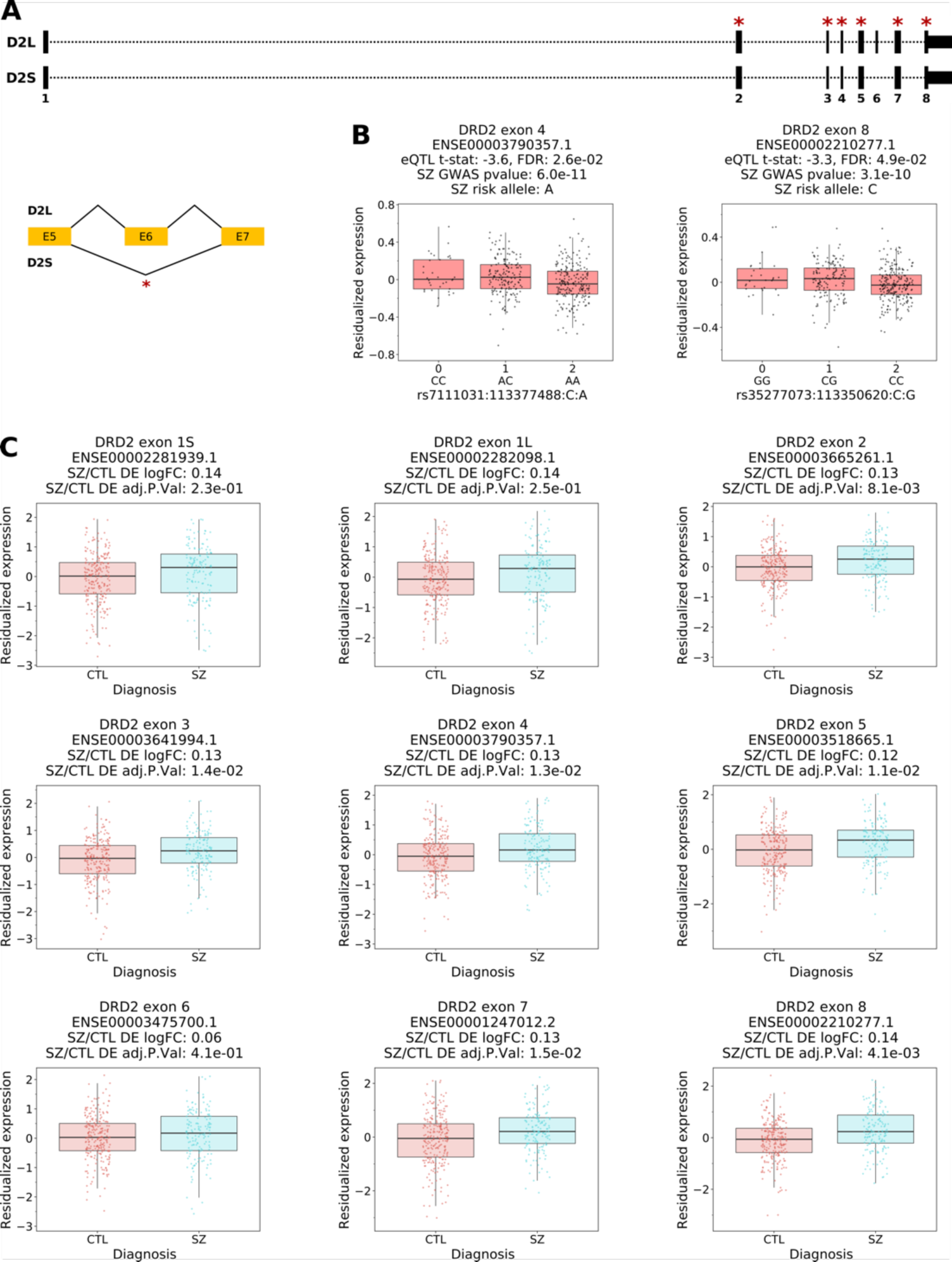
Decreased expression of exons shared between the dopamine receptor D2 (DRD2) short (D2S) and long (D2L) isoforms are associated with increased schizophrenia (SZ) risk (PGC2+CLOZUK GWAS p-value < 5e-8) with significant upregulation in schizophrenia (adjusted p-value < 0.05) in the caudate nucleus. A. *DRD2* annotation of D2S and D2L with exons and D2S specific exon-exon junction that are significantly upregulated (adjusted p-value < 0.05) denoted with a red asterisk. **B.** Boxplots of DRD2 exon eQTL with SNP associated with schizophrenia risk (GWAS p-value < 5e-8). **C.** Boxplots of differential expression for all DRD2 exons. SZ – schizophrenia, CTL – neurotypical controls, logFC – log2 transformation of fold change, DE – differential expression, and t-stat – t-statistic.

**Fig. S8.**
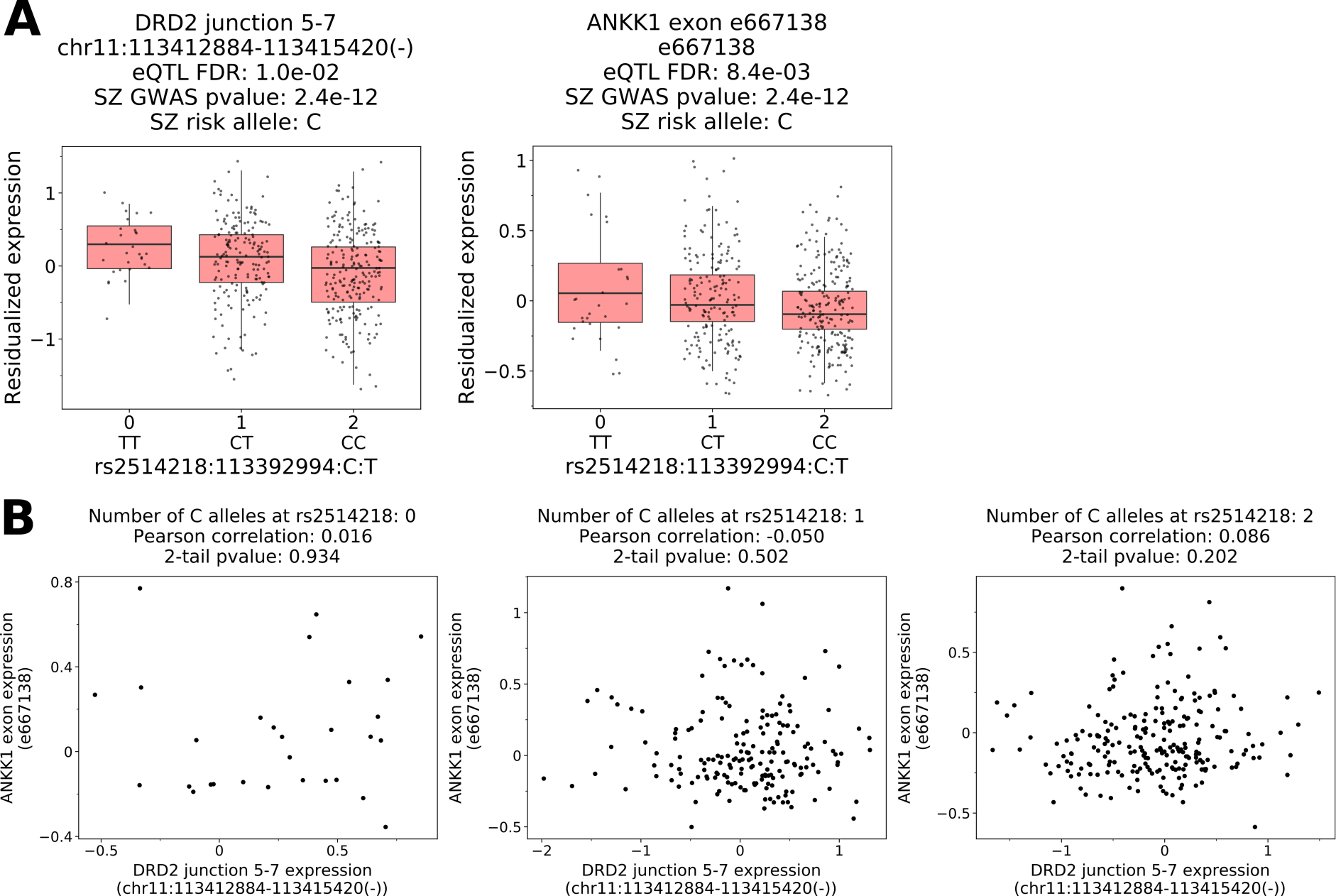
Expression of *ANKK1* exon e667138 and *DRD2* junction 5-7 eQTLs are uncorrelated, conditional on the genotype at SZ risk SNP rs2514218. A. Boxplot of *DRD2* junction 5-7 and *ANKK1* exon (e667138) showing eQTL association to SNP rs2514218. **B.** Scatter plot of *ANKK1* exon expression and *DRD2* junction 5-7 expression shows no correlation for all genotypes at SNP rs2514218.

**Fig. S9.**
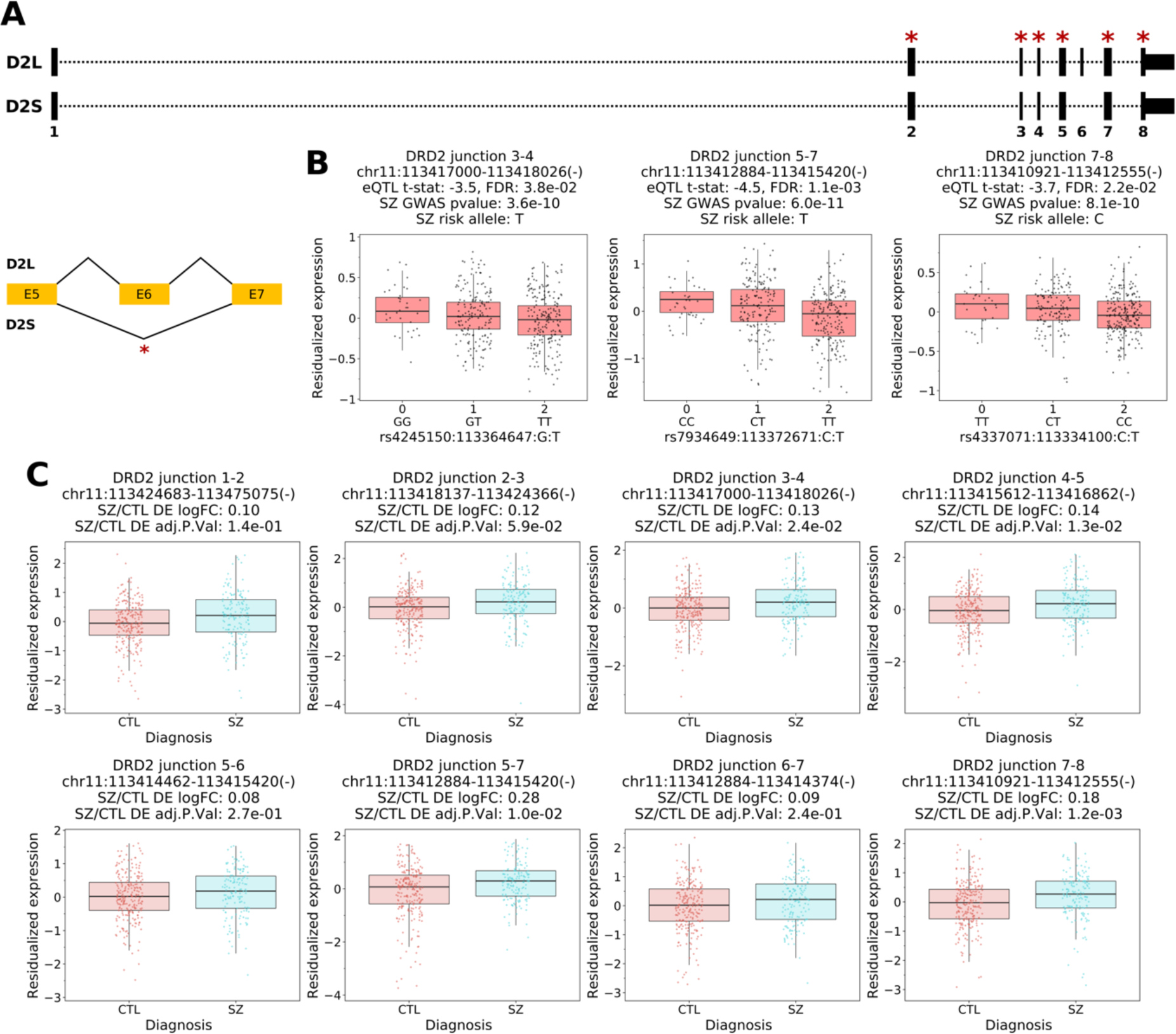
Decreased expression of junctions associated with the dopamine receptor D2 (DRD2) short isoform (D2S) and not the long (D2L) isoform is associated with increased schizophrenia (SZ) risk (PGC2+CLOZUK GWAS p-value < 5e-8) and significant upregulation in schizophrenia (adjusted p-value < 0.05) in the caudate nucleus. A. *DRD2* annotation of D2S and D2L with exons and D2S specific exon-exon junction that are significantly upregulated (adjusted p-value < 0.05) denoted with a red asterisk. **B.** Boxplots of DRD2 junctions eQTL with SNP associated with schizophrenia risk (GWAS p-value < 5e-8). **C.** Boxplots of differential expression for all DRD2 junctions. SZ – schizophrenia, CTL – neurotypical controls, logFC – log2 transformation of fold change, DE – differential expression, and t-stat – t-statistic.

**Fig. S10.**
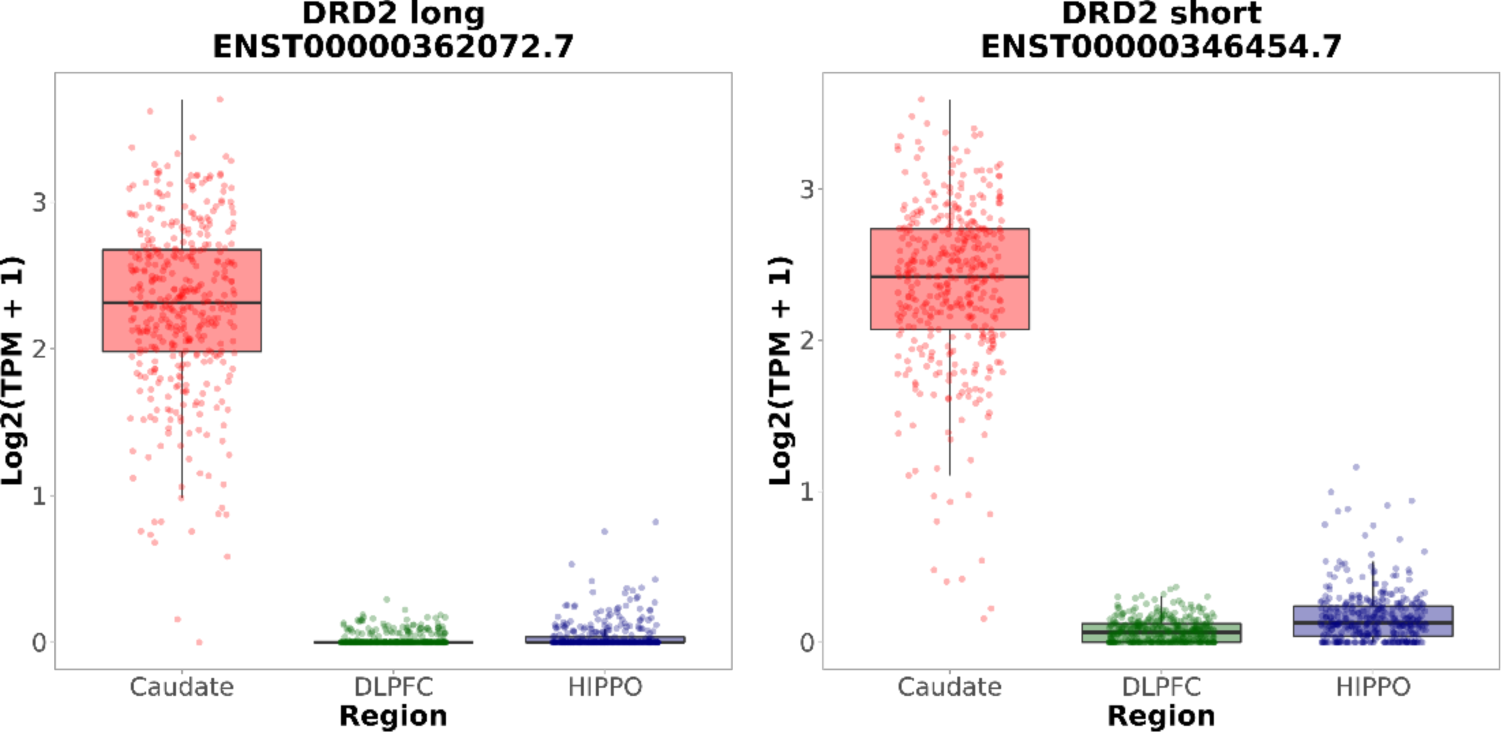
The short and long isoforms of the dopamine receptor D2 (DRD2) are abundantly expressed in the caudate nucleus but not the DLPFC nor the hippocampus. Boxplot of *DRD2* long and short expression across caudate nucleus, dorsolateral prefrontal cortex (DLPFC), and hippocampus (HIPPO).

**Fig. S11.**
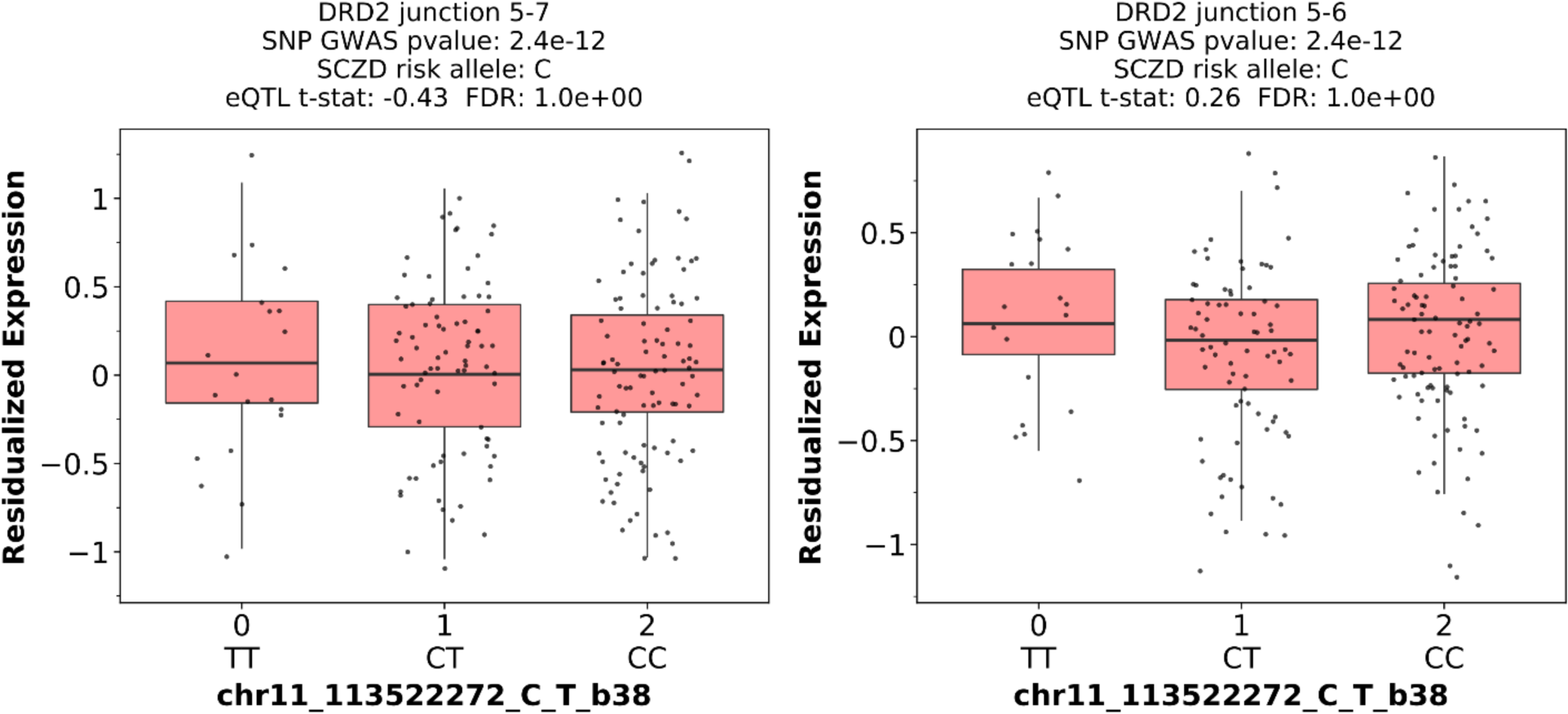
Expression of DRD2 exon junction 5-7 in GTEx v8 caudate nucleus samples as a function of schizophrenia (SCZD) risk allele dosage at the index SNP rs2514218 (GTEx SNP ID chr11_113522272_C_T_b38). Even though there is not statistically significant eQTL association in GTEx samples, the direction of association is the same as in our data (Fig 3).

**Fig. S12.**
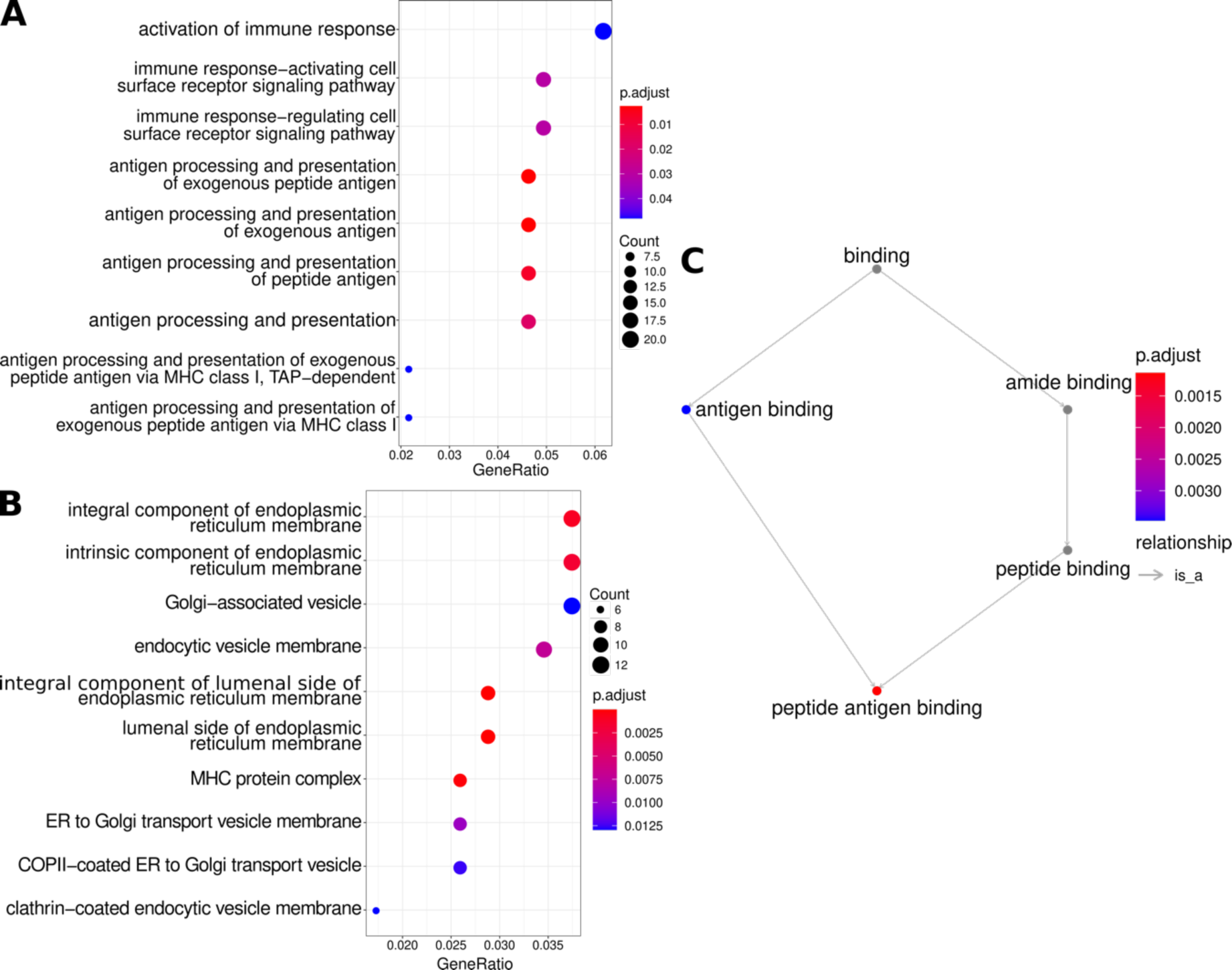
Enrichment of immune related pathways for significant TWAS associations (FDR < 0.05) in the caudate nucleus. Gene ontology enrichment analysis dotplots of **A.** cellular components and **B**. biological processes, and **C**. GO network of molecular function.

**Fig. S13.**
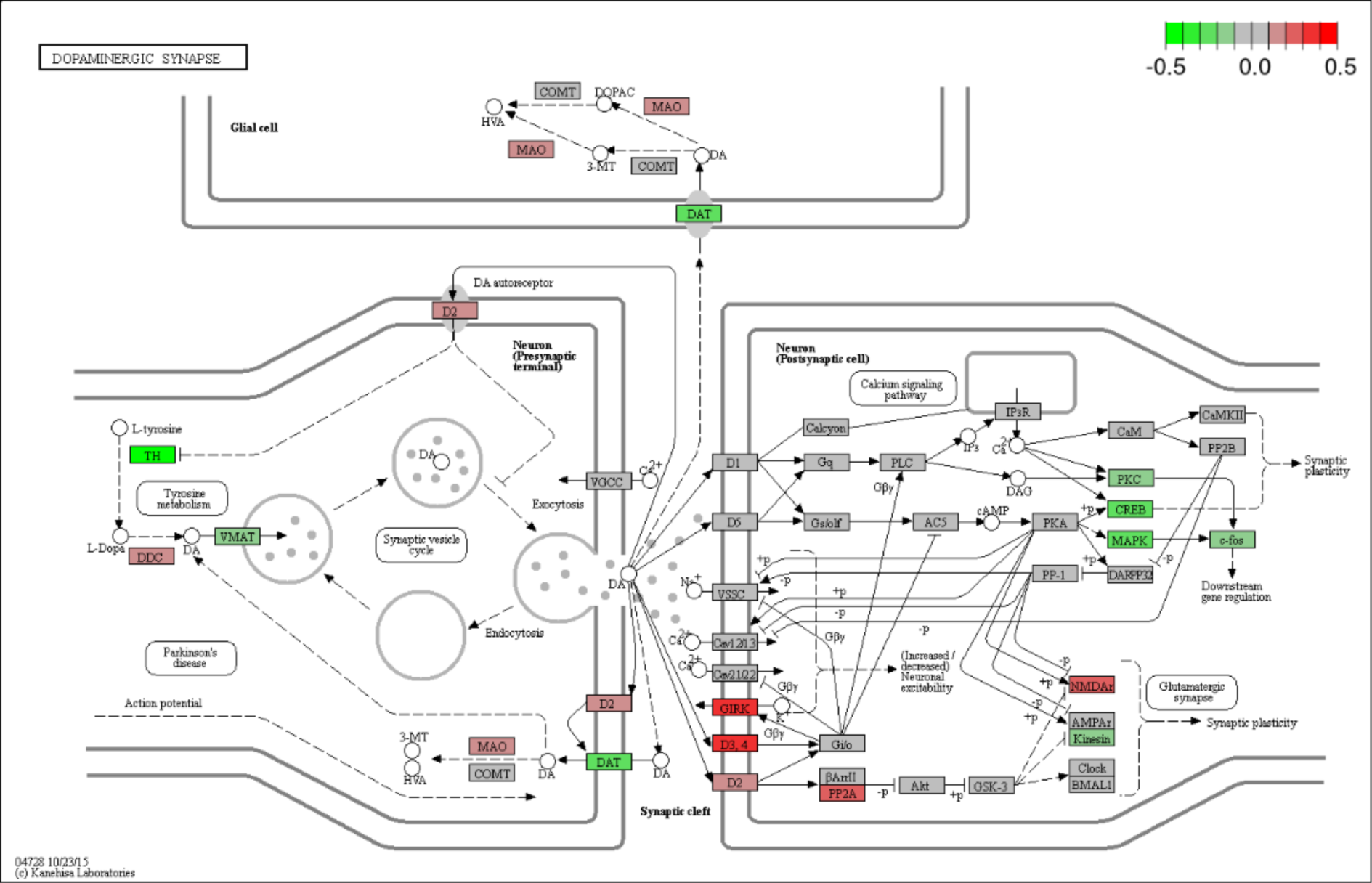
Dopaminergic synapses KEGG pathway demonstrating upregulation of dopamine receptors and downregulation of tyrosine hydroxylase from caudate nucleus differential expression results. Bar scale legend is log2 fold change in SZ vs control (red means higher expression in SZ). KEGG ID: hsa04728.

**Fig. S14.**
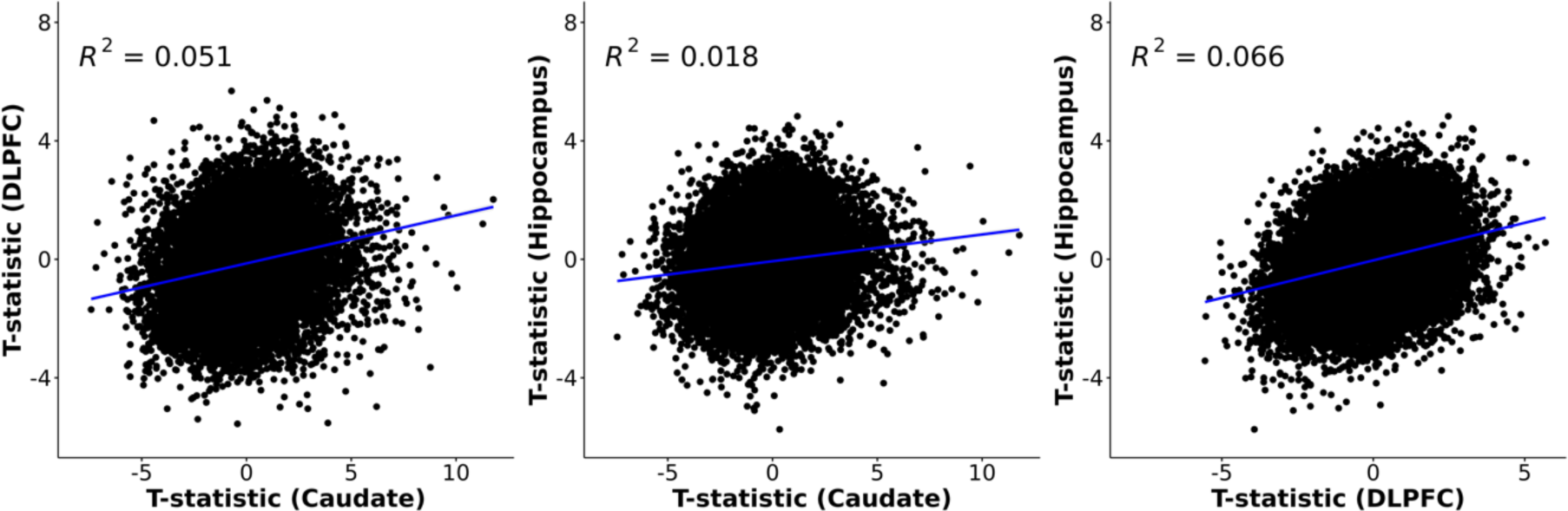
Differential expression analysis (schizophrenia vs control) shows small but positive correlation of t-statistics for caudate compared to DLPFC and hippocampus (Spearman, p-value < 0.001).

**Fig. S15.**
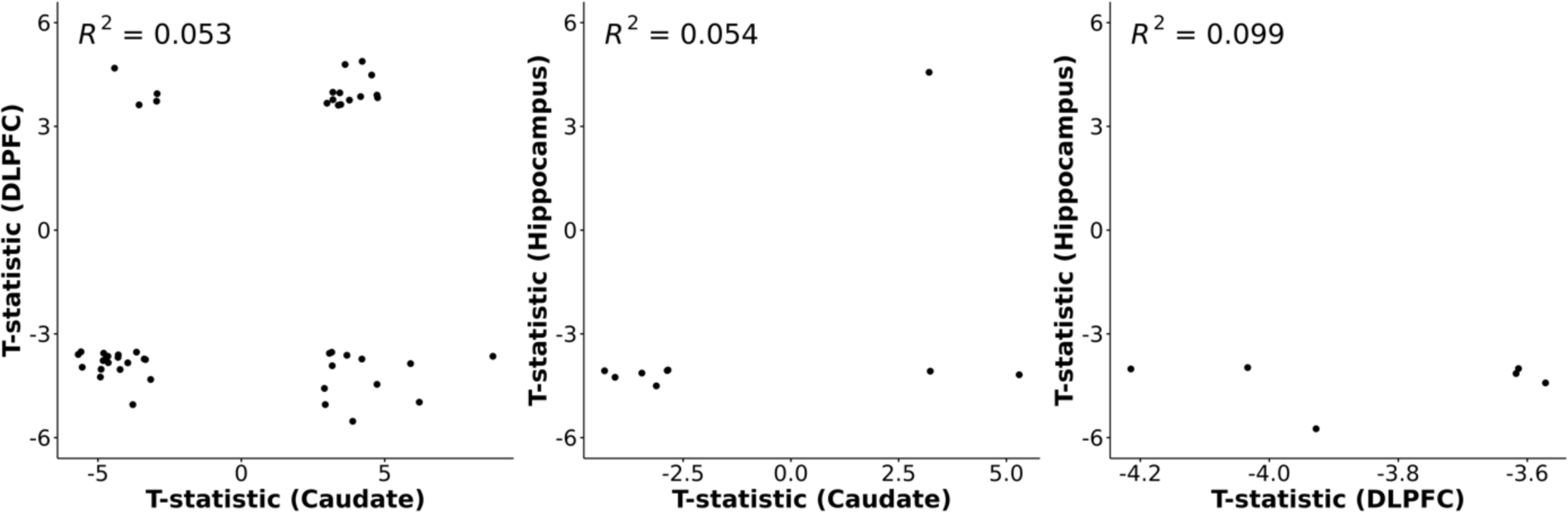
Comparison of t-statistic for significantly differentially expressed genes in three brain regions. Significantly differentially expressed genes (FDR < 0.05, schizophrenia vs control) show both concordant and discordant direction of effect (t-statistic) in the caudate nucleus compared with the DLPFC or hippocampus.

**Fig. S16.**
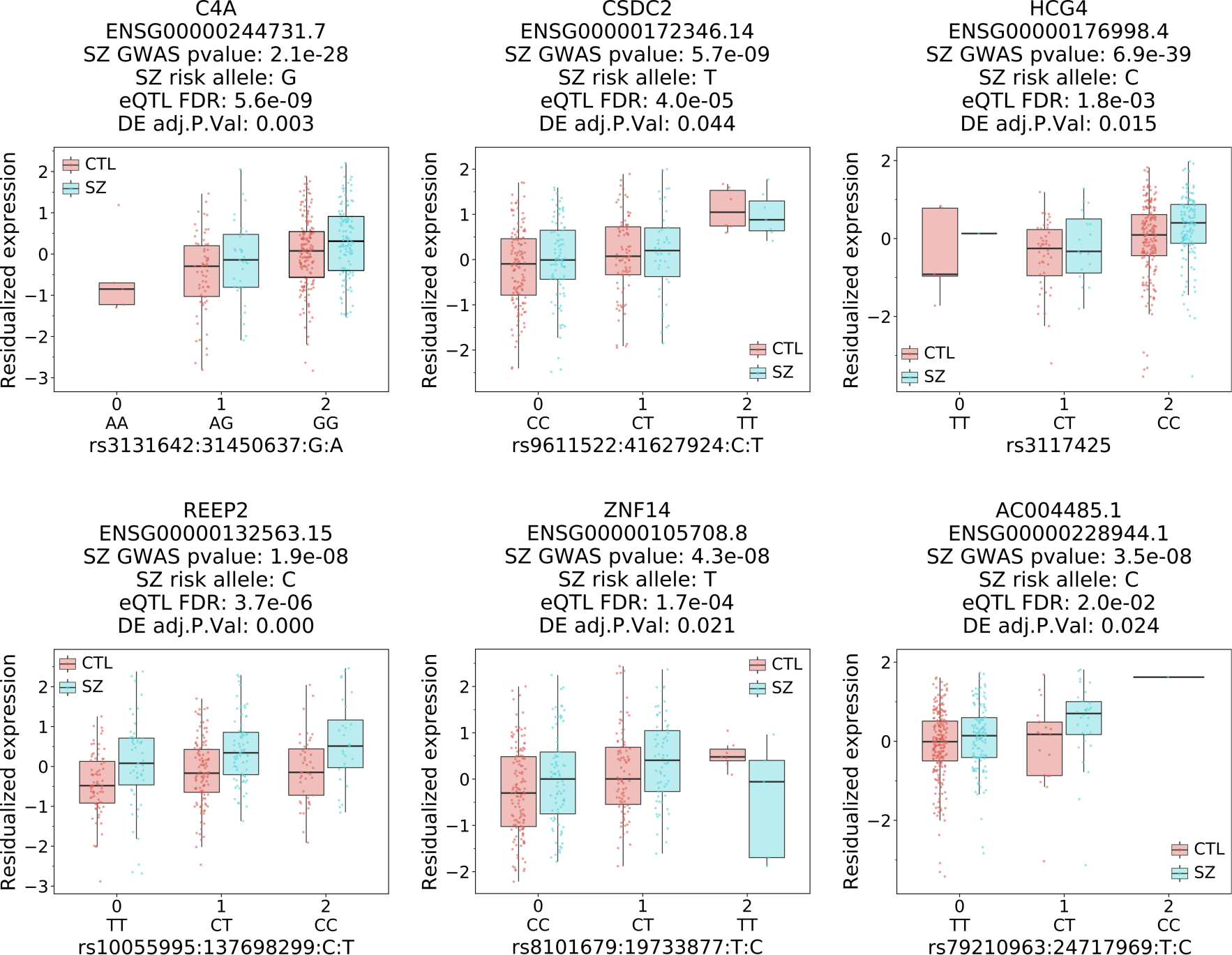
Six genes show same direction of effect of an increase in schizophrenia risk with an increase in gene expression as well as upregulation in schizophrenia compared to neurotypical controls. Boxplots of residualized gene expression in the caudate nucleus for unique genes based on gene-SNP pairs that are associated with schizophrenia risk (GWAS p- value < 5e-8, eQTL FDR < 0.05), significantly differentially expressed (FDR < 0.05) and have concordant directionality.

**Fig. S17.**
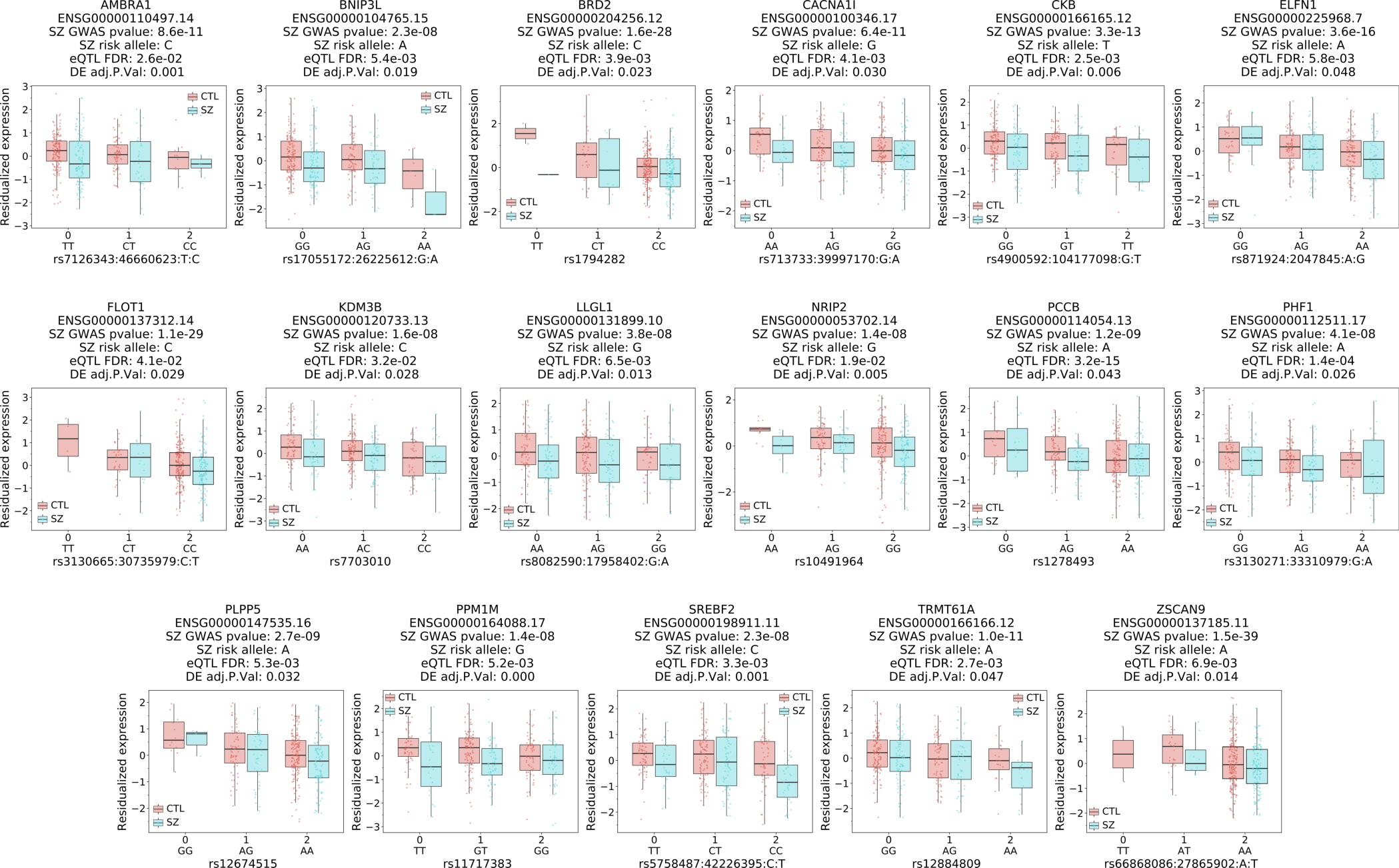
Seventeen genes show same direction of effect of an increase in schizophrenia risk with a decrease in gene expression as well as downregulation in schizophrenia compared to neurotypical controls. Boxplots of residualized gene expression in the caudate nucleus for unique genes based on gene-SNP pairs associated with schizophrenia risk (GWAS p-value < 5e- 8, eQTL FDR < 0.05), significant differential expression (FDR < 0.05) and have concordant directionality.

**Fig. S18.**
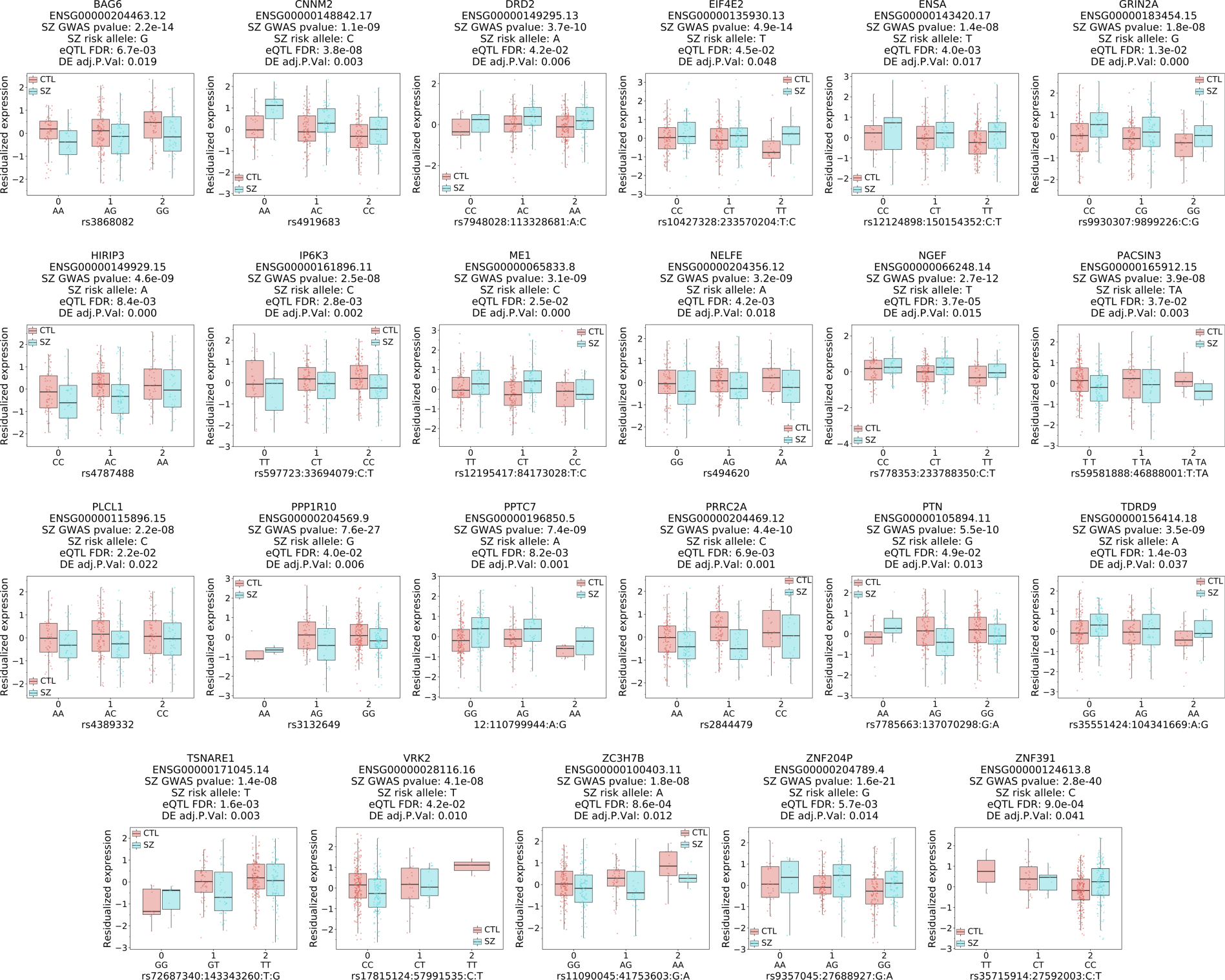
Twenty-three genes show different direction of effect including *DRD2*. Boxplots of residualized gene expression in the caudate nucleus for unique genes based on gene-SNP pairs associated with schizophrenia risk (GWAS p-value < 5e-8, eQTL FDR < 0.05), significant differential expression (FDR < 0.05) and have discordant directionality.

**Fig. S19.**
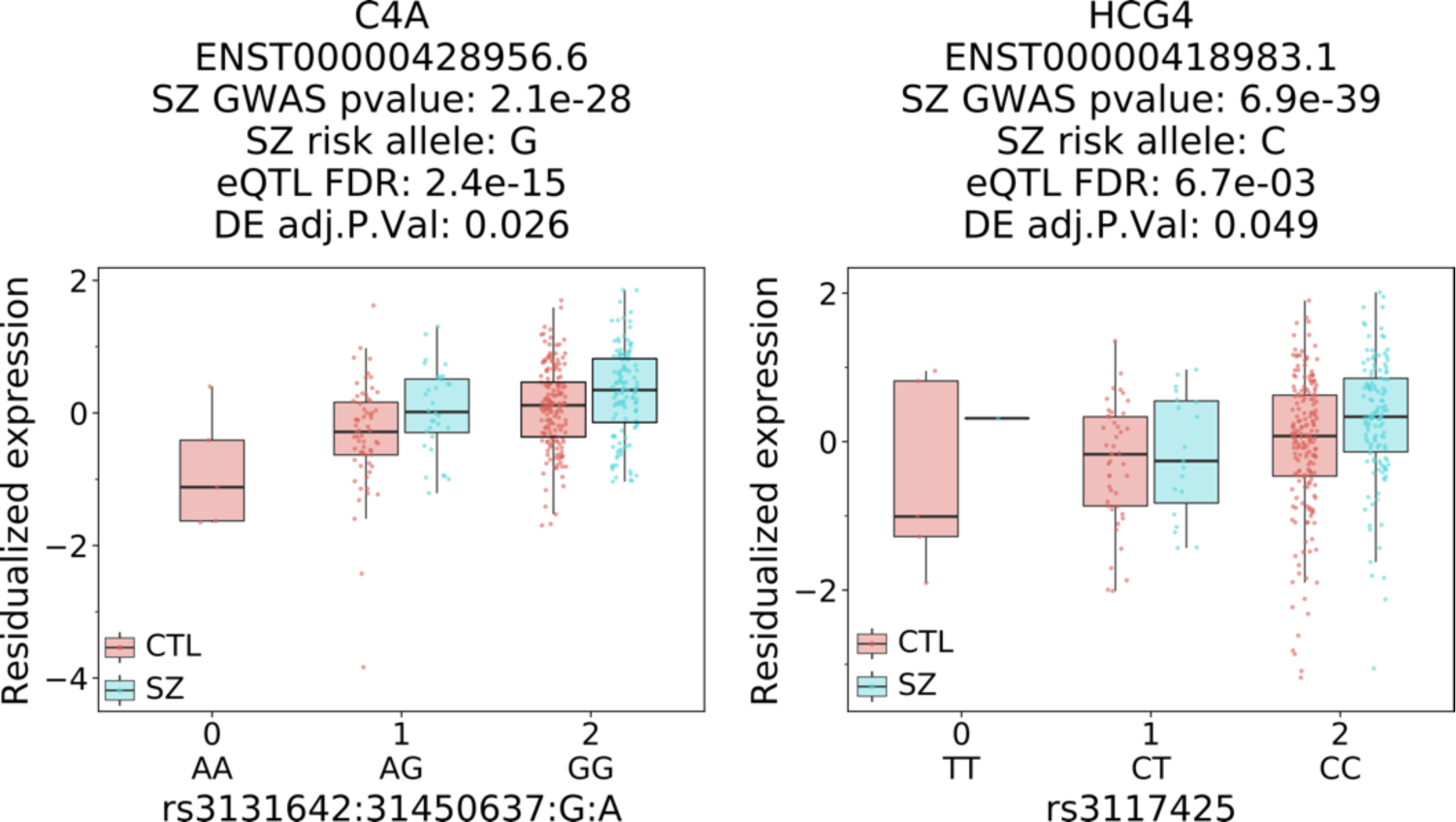
Two transcripts show same direction of effect of an increase in schizophrenia risk with an increase in gene expression as well as upregulation in schizophrenia compared to neurotypical controls. Boxplots of residualized expression in the caudate nucleus for unique transcripts based on transcript-SNP pairs associated with schizophrenia risk (GWAS p-value < 5e-8, eQTL FDR < 0.05), significant differential expression (FDR < 0.05) and have concordant directionality.

**Fig. S20.**
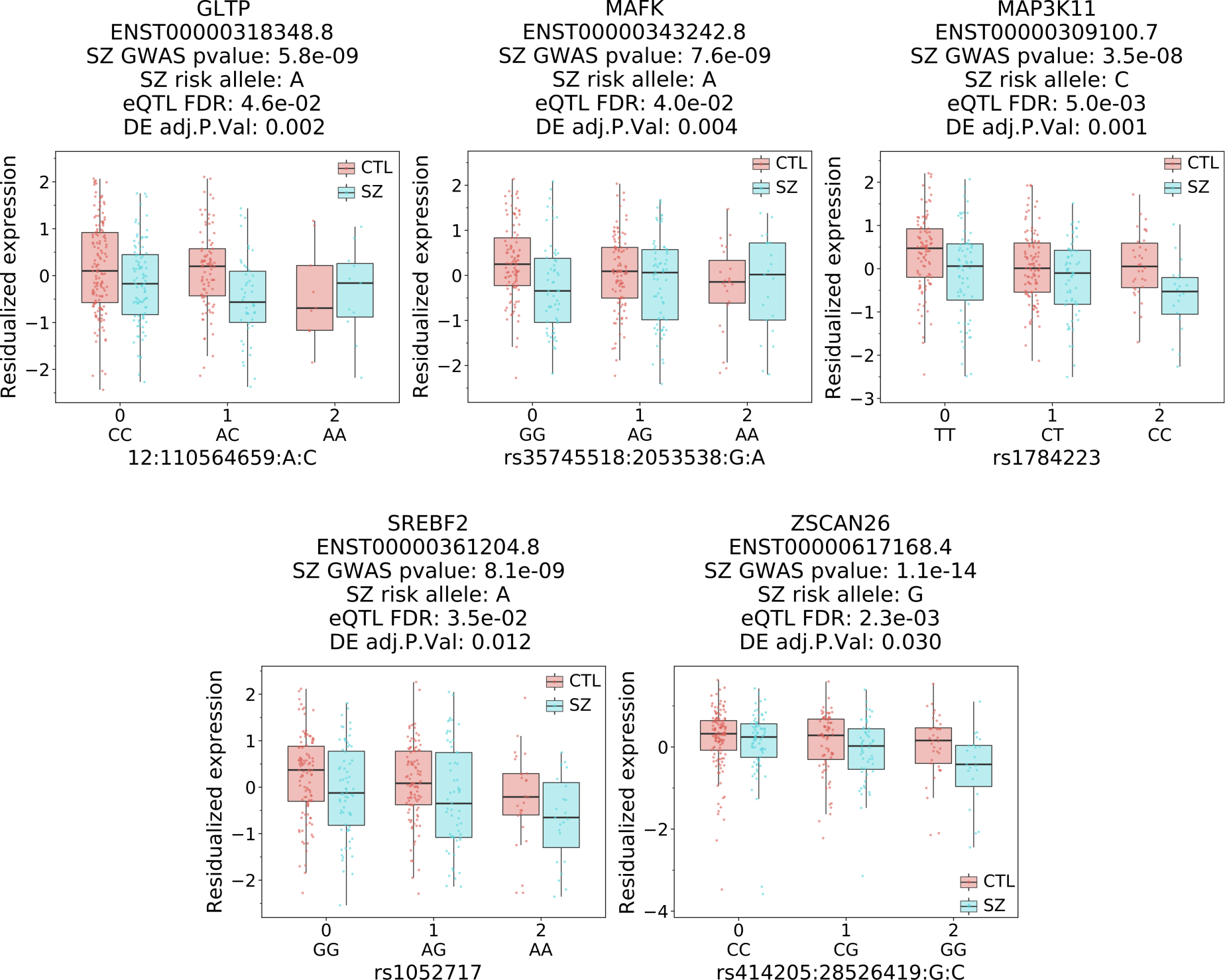
Five transcripts show same direction of effect of an increase in schizophrenia risk with a decrease in gene expression as well as downregulation in schizophrenia compared to neurotypical controls. Boxplots of residualized expression in the caudate nucleus for unique transcripts based on gene-SNP pairs associated with schizophrenia risk (GWAS p-value < 5e-8, eQTL FDR < 0.05), significant differential expression (FDR < 0.05) and have concordant directionality.

**Fig. S21.**
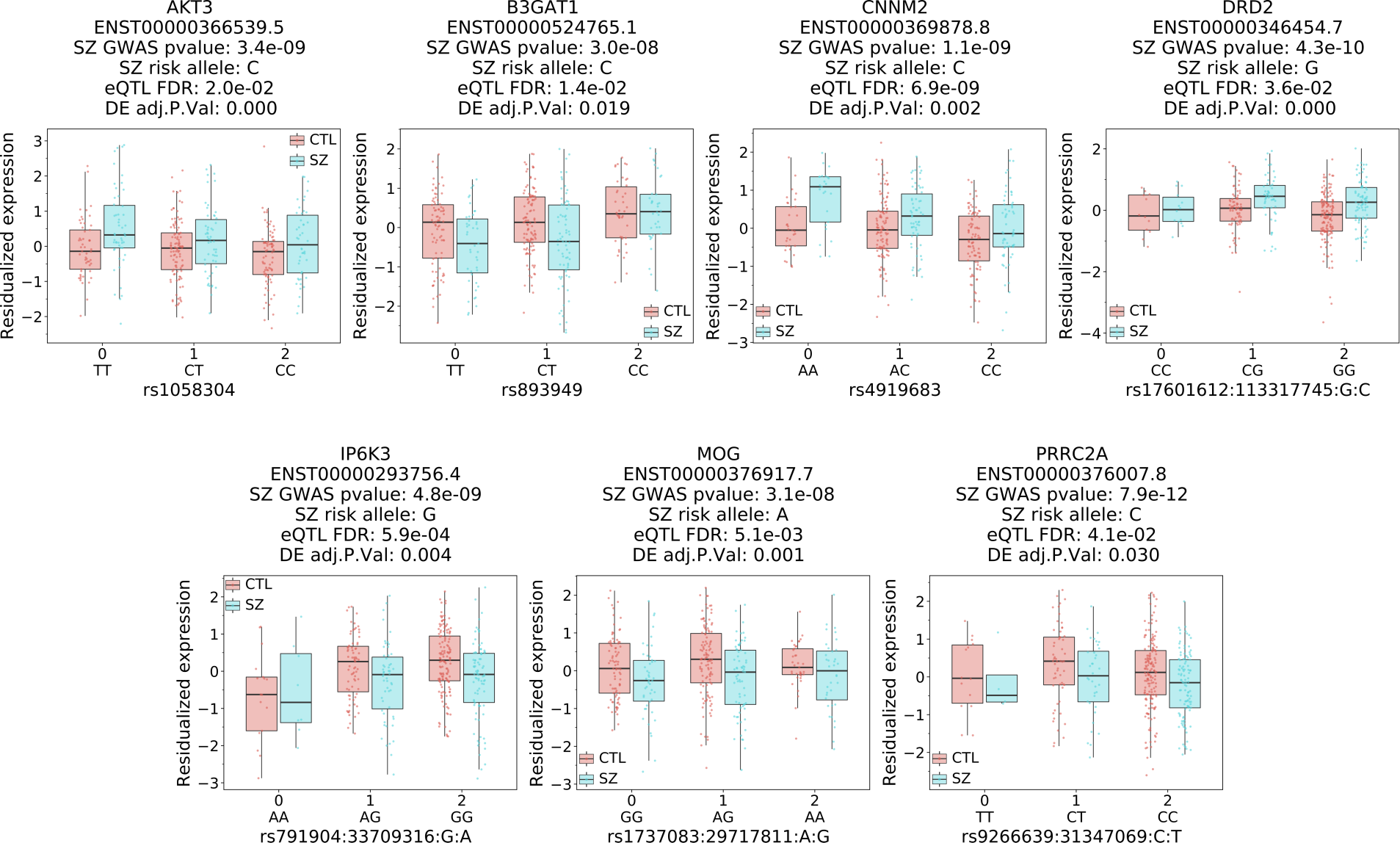
Seven transcripts show different direction of effect including *DRD2*. Boxplots of residualized expression in the caudate nucleus for unique transcripts based on transcript-SNP pairs associated with schizophrenia risk (GWAS p-value < 5e-8, eQTL FDR < 0.05), significant differential expression (FDR < 0.05) and have discordant directionality.

**Fig. S22.**
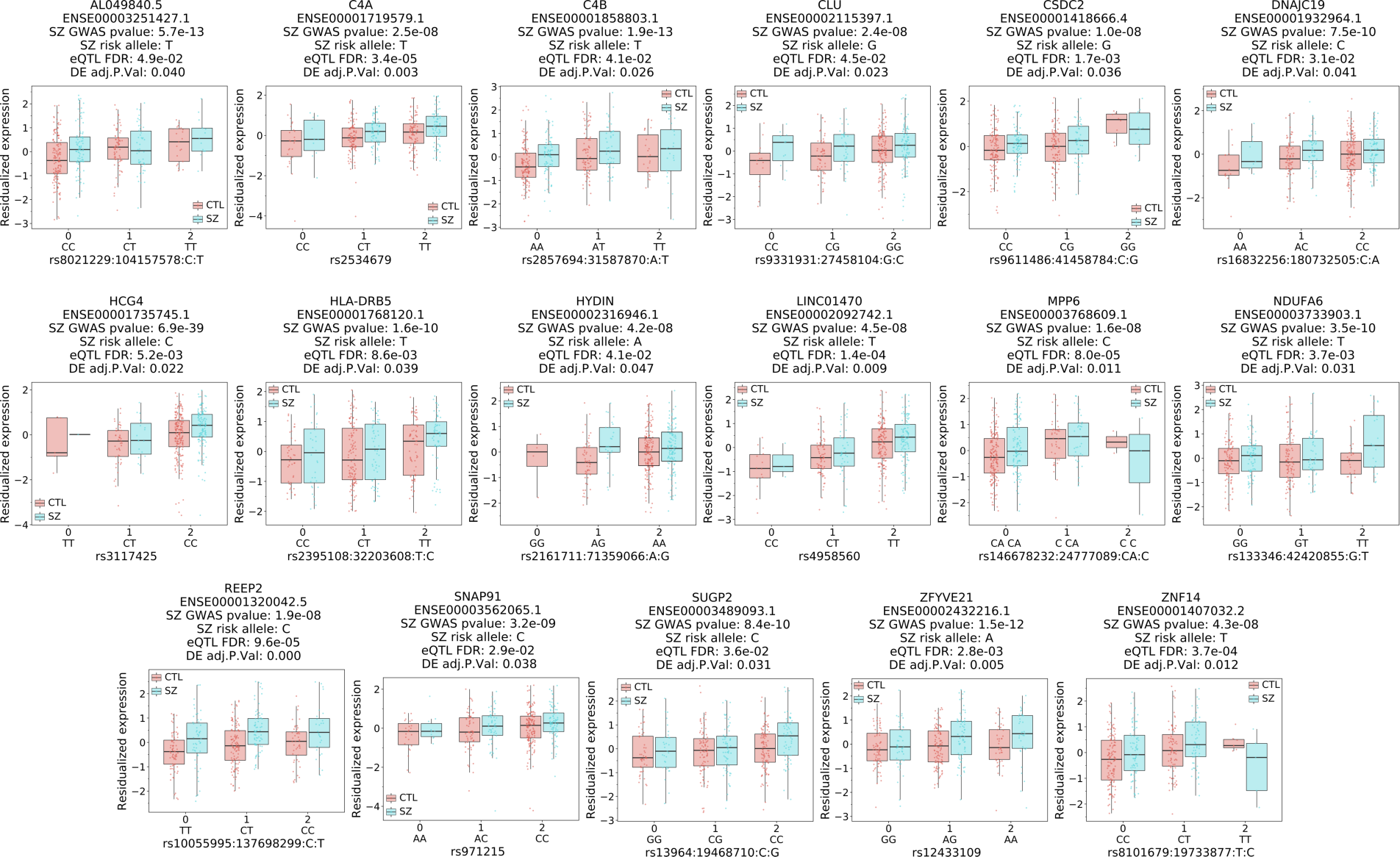
Seventeen genes show same direction of effect of an increase in schizophrenia risk with an increase in exon expression as well as upregulation in schizophrenia compared to neurotypical controls. Representative boxplots of residualized expression in the caudate nucleus for unique genes with at least one exon with a SNP associated with schizophrenia risk (GWAS p-value < 5e-8, eQTL FDR < 0.05), significant differential expression (FDR < 0.05) and have concordant directionality.

**Fig. S23.**
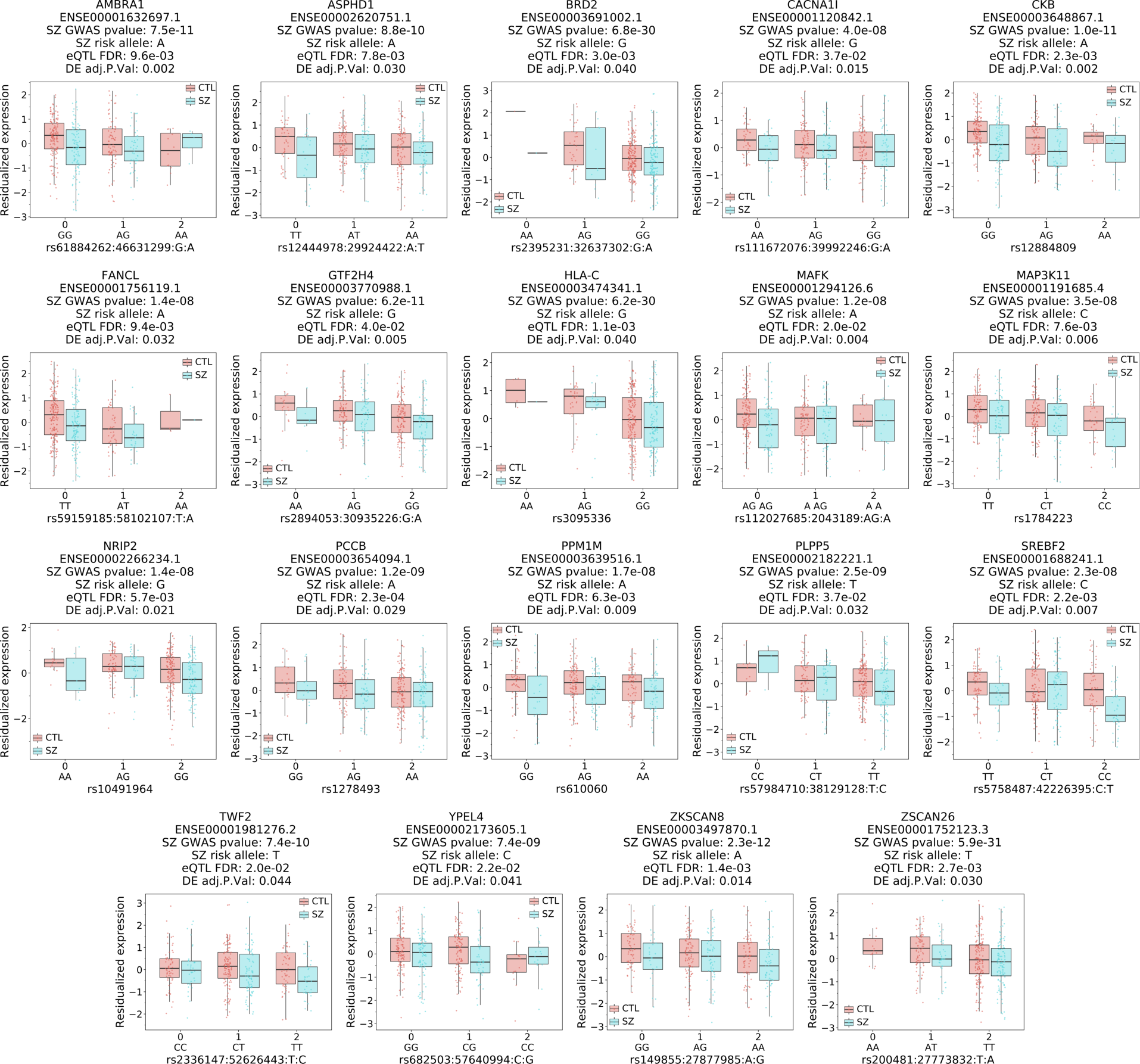
Nineteen genes show same direction of effect of an increase in schizophrenia risk with a decrease in exon expression as well as downregulation in schizophrenia compared to neurotypical controls. Representative boxplots of residualized expression in the caudate nucleus for unique genes with at least one exon with a SNP associated with schizophrenia risk (GWAS p-value < 5e-8, eQTL FDR < 0.05), significant differential expression (FDR < 0.05) and have concordant directionality.

**Fig. S24.**
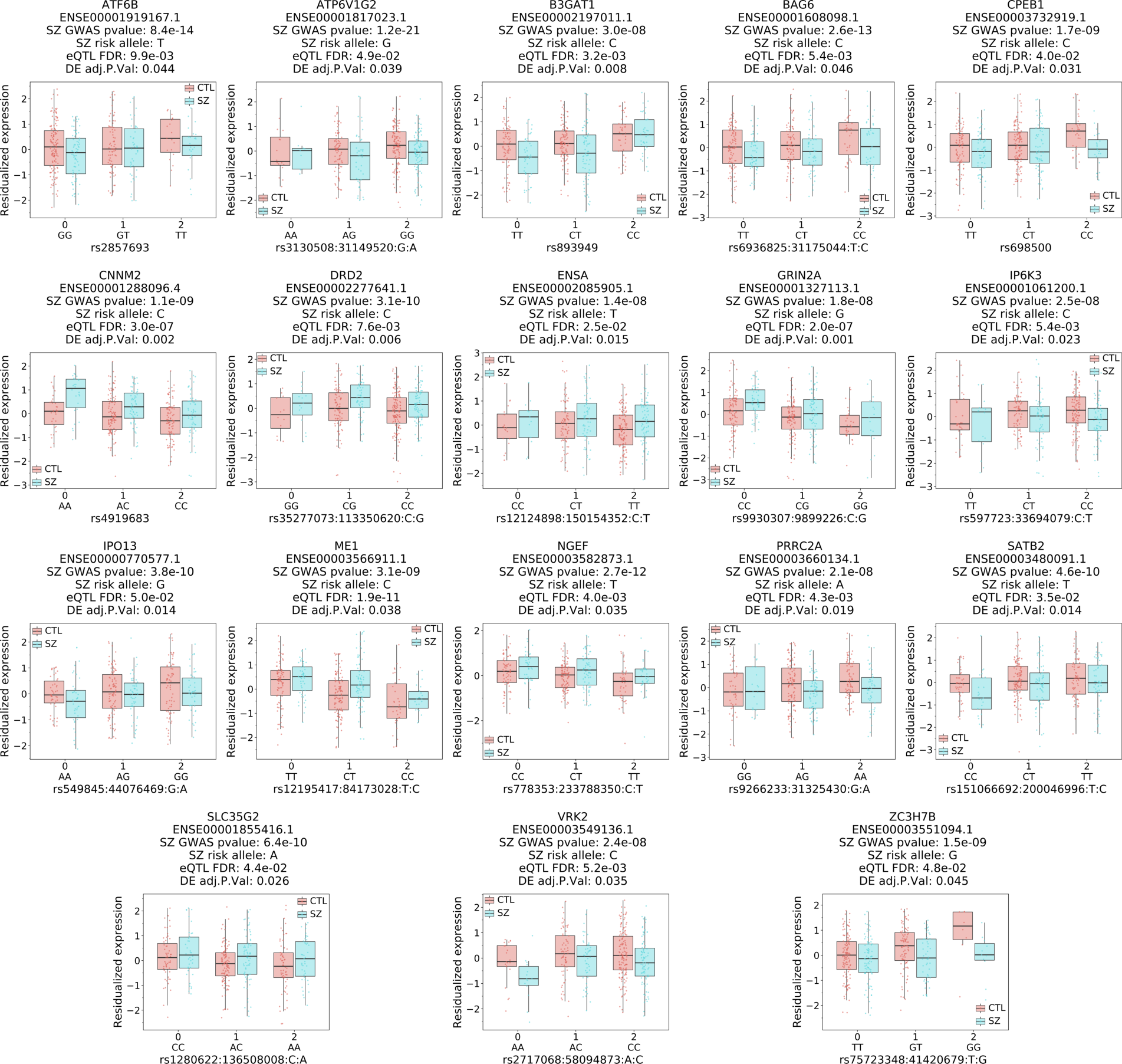
Eighteen genes show different direction of effect for exon-SNP associations including *DRD2*. Representative boxplots of residualized gene expression in the caudate nucleus for unique genes with at least one exon with a SNP associated with schizophrenia risk (GWAS p- value < 5e-8, eQTL FDR < 0.05), significant differential expression (FDR < 0.05) and have discordant directionality.

**Fig. S25.**
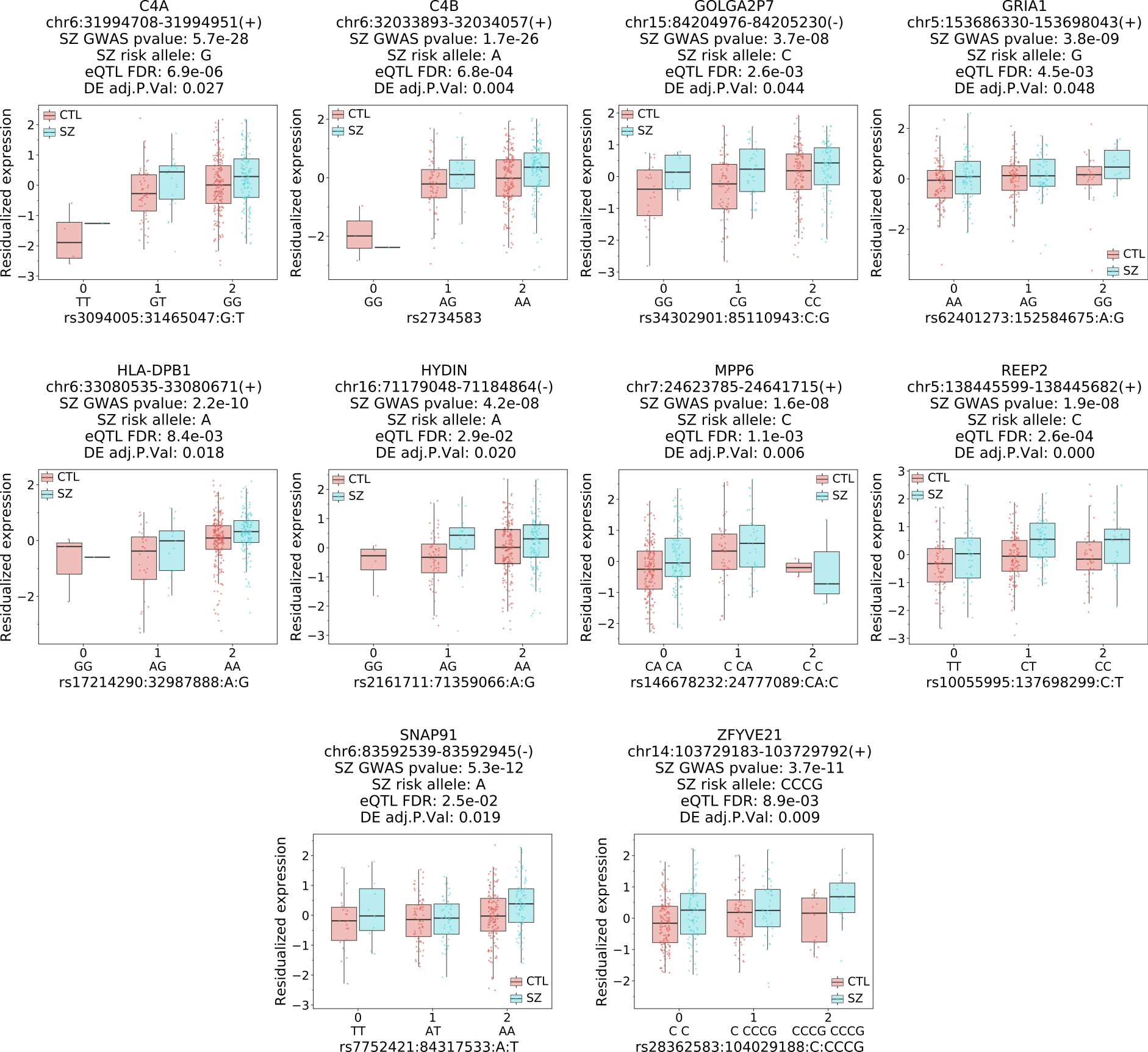
Ten unique genes show same direction of effect of an increase in schizophrenia risk with an increase in exon-exon junction expression as well as upregulation in schizophrenia compared to neurotypical controls. Representative boxplots of residualized expression in the caudate nucleus for unique genes with at least one junction with a SNP associated with schizophrenia risk (GWAS p-value < 5e-8, eQTL FDR < 0.05), significant differential expression (FDR < 0.05) and have concordant directionality.

**Fig. S26.**
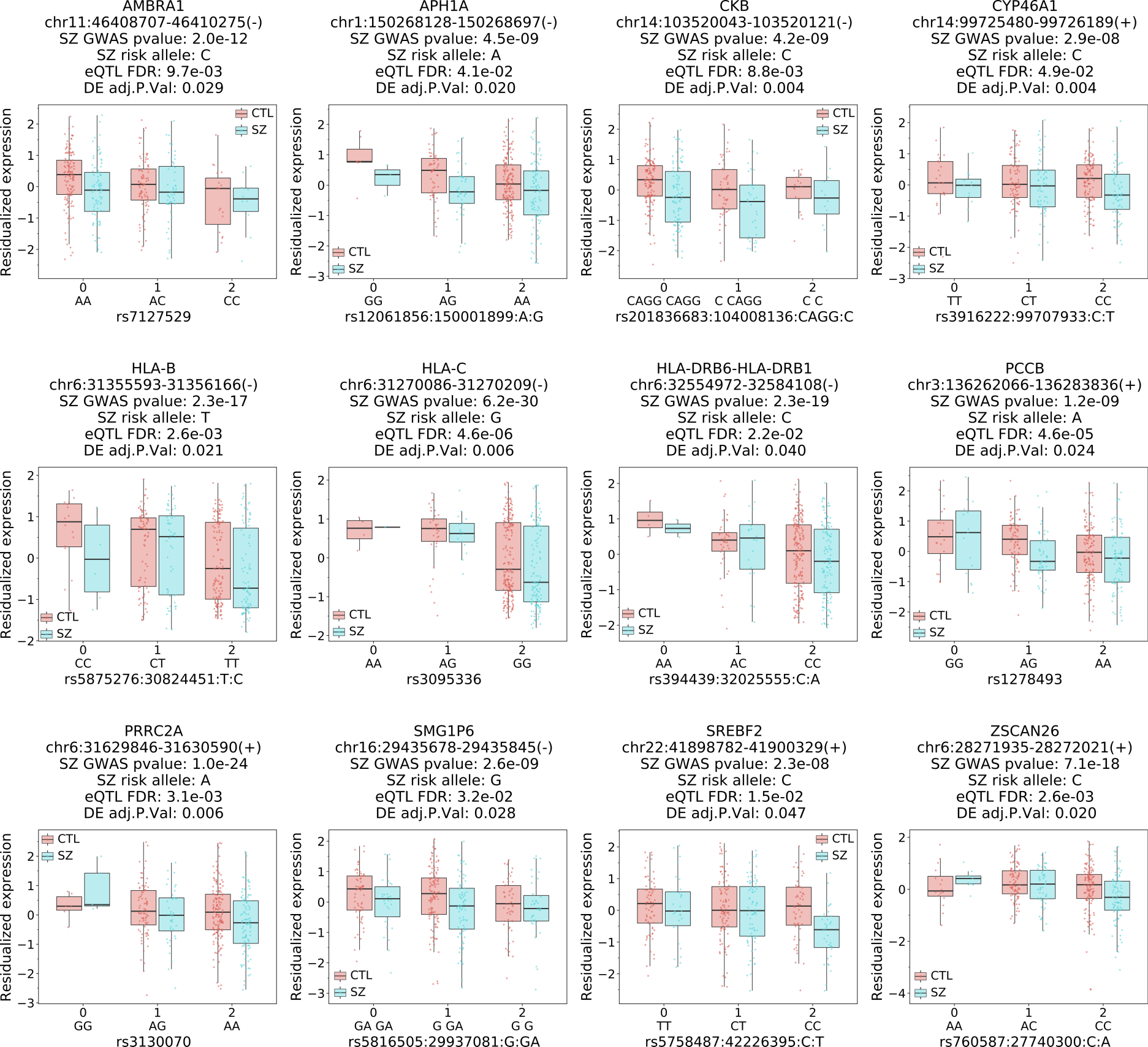
Twelve unique genes show same direction of effect of an increase in schizophrenia risk with a decrease in exon-exon junction expression as well as downregulation in schizophrenia compared to neurotypical controls. Representative boxplots of residualized expression in the caudate nucleus for unique genes with at least one junction with a SNP associated with schizophrenia risk (GWAS p-value < 5e-8, eQTL FDR < 0.05), significant differential expression (FDR < 0.05) and have concordant directionality.

**Fig. S27.**
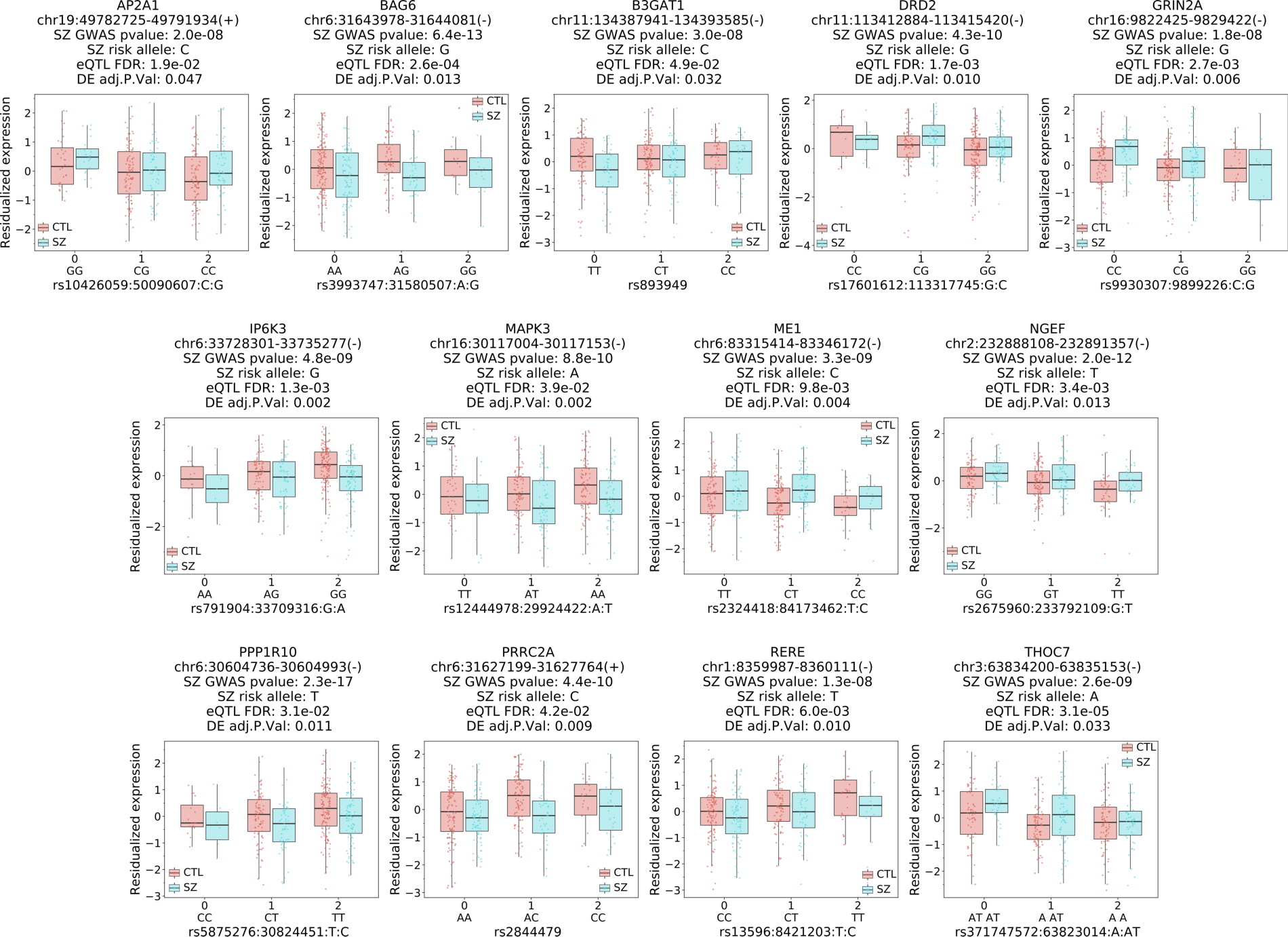
Thirteen genes show different direction of effect for SNP-junction associations including *DRD2*. Representative boxplots of residualized gene expression in the caudate nucleus for unique genes with at least one junction with a SNP associated with schizophrenia risk (GWAS p- value < 5e-8, eQTL FDR < 0.05), significant differential expression (FDR < 0.05) and have discordant directionality.

**Fig. S28.**
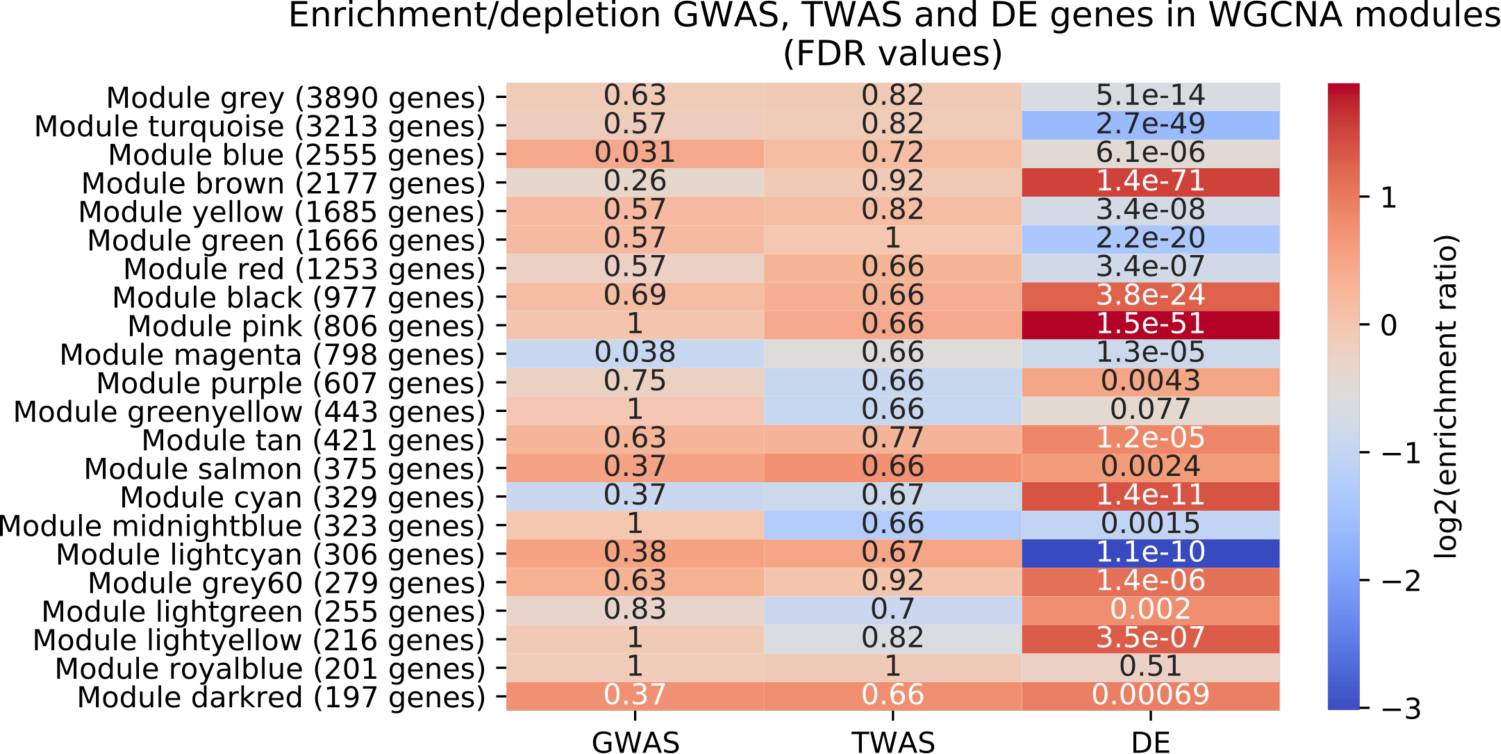
*DRD2* short (junction 5-7; cyan) and long isoforms (junction 5-6, 6-7; purple) in separate modules. Heatmap of WGCNA modules showing significant enrichment (hypergeometric test) for schizophrenia DEGs across 20 of 22 modules but virtually no enrichment for GWAS or TWAS genes.

**Fig. S29.**
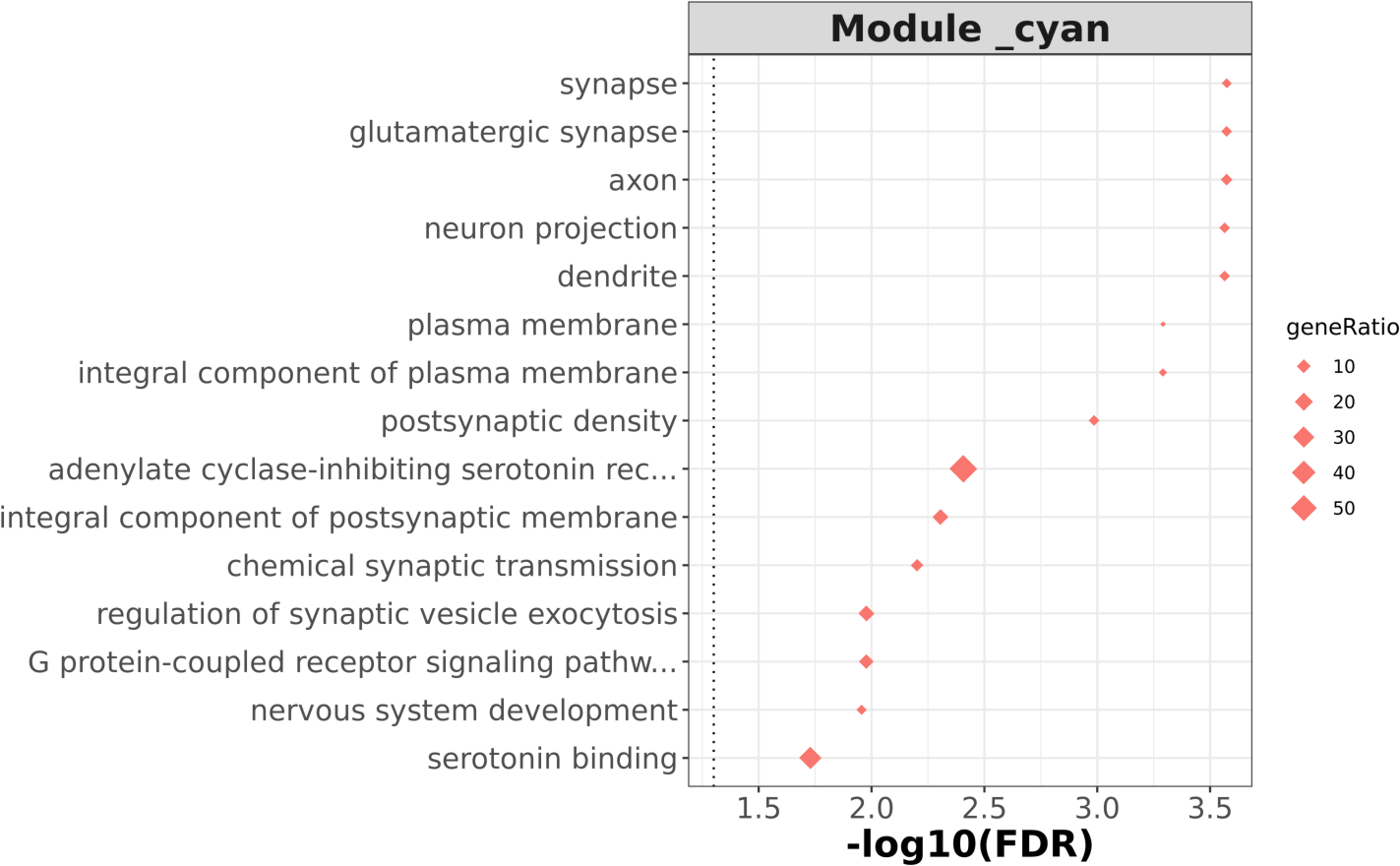
*T*op 15 enriched GO terms for *DRD2* short (junction 5-7) cyan module shows significant enrichment for synapse.

**Fig. S30.**
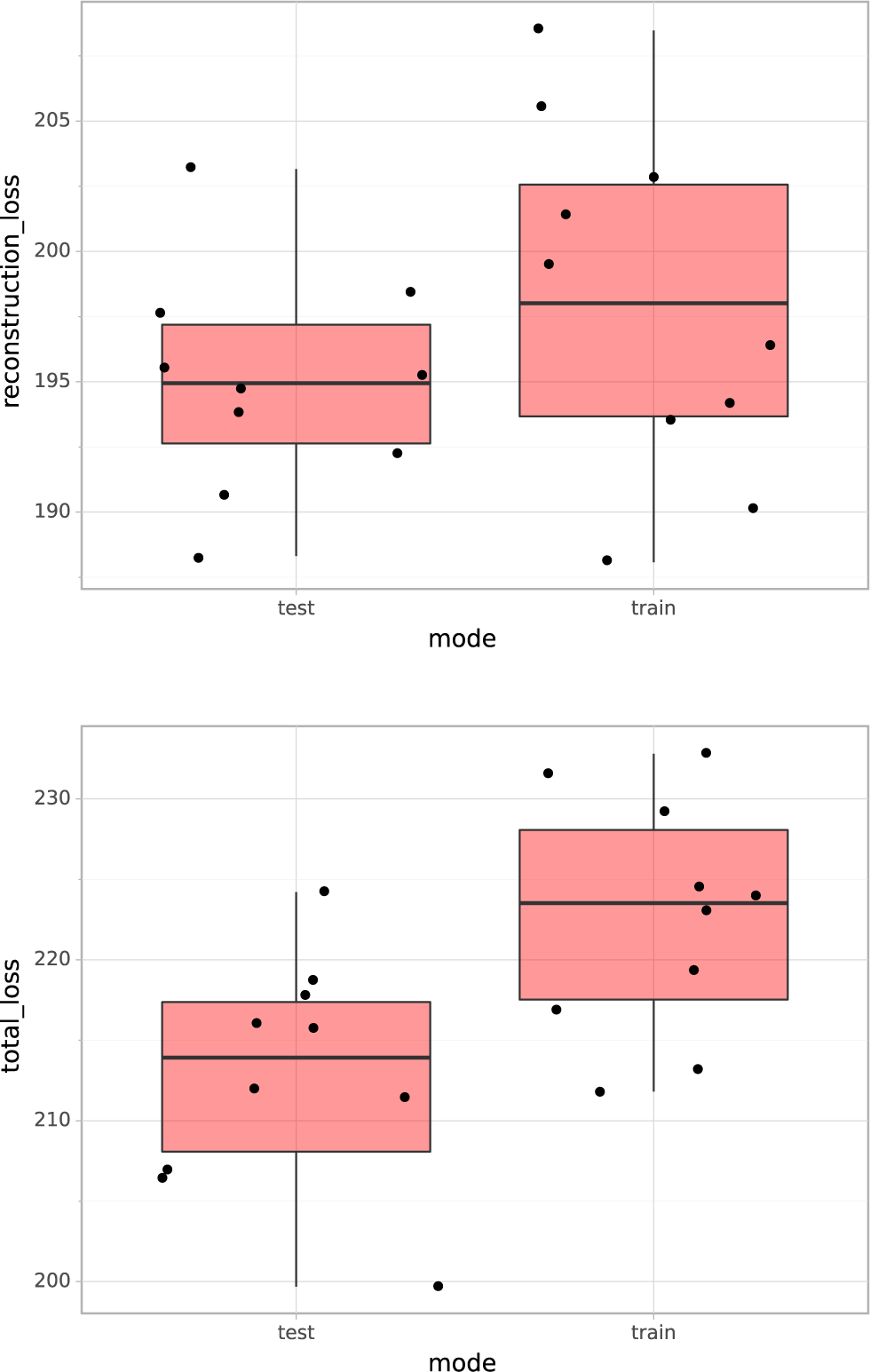
Cross-validation analysis of GNVAE. We perform 10-fold cross-validation on the GNVAE and examine reconstruction loss (top) and total loss (bottom) on the training and test set. Each point corresponds to a different train/test split of the data. The fact that the losses are not smaller in training set than in test set indicates the autoencoder is not overfitting the training examples.

**Fig. S31.**
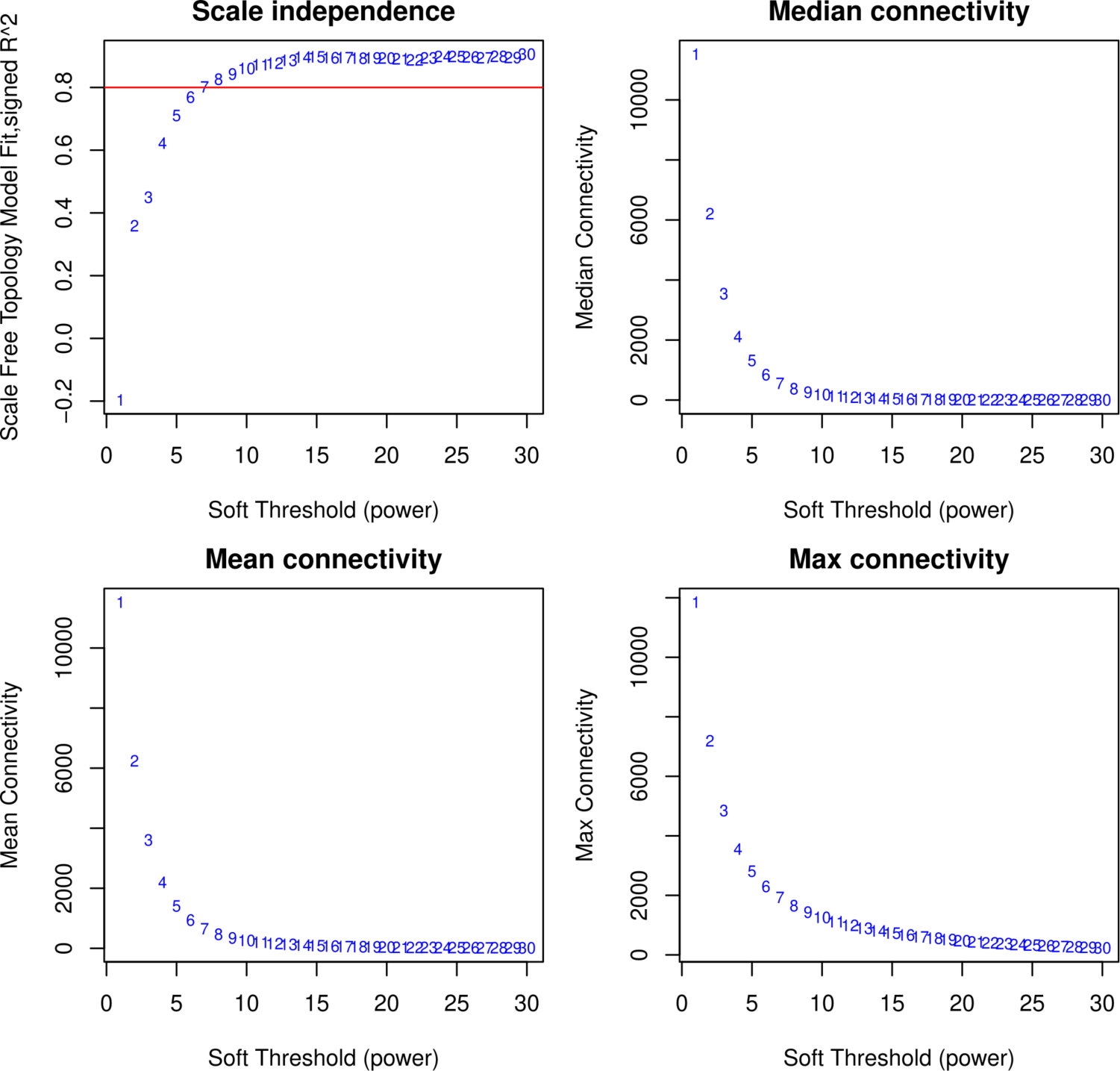
Scale-Free Topology and connectivity metrics for WGCNA analysis in the caudate nucleus.

**Table S1.**
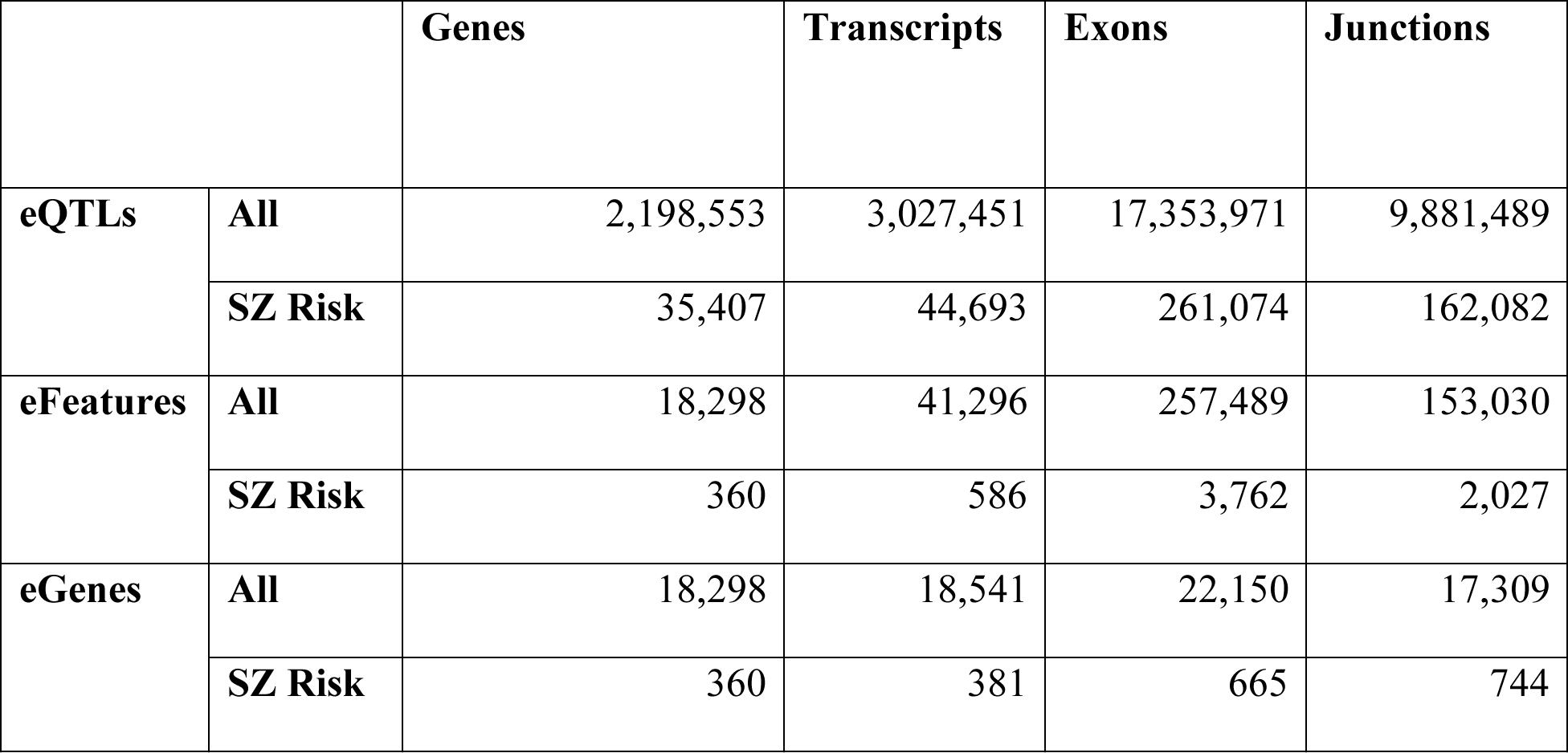
Summary of eQTLs (FDR < 0.05), unique features, and unique genes for all eQTLs and those with GWAS-significant SNPs (GWAS p-value < 5e-8) according to the PGC2 + CLOZUK GWAS. eQTLs: number of SNP-feature associations (eQTLs) found in the caudate nucleus, for each of the features: genes transcripts, exons, and junctions. eFeatures: number of unique features that have eQTL associations. eGenes: number of unique genes for each eFeature.

**Table S2.** Directionality comparison of gene-level cis-eQTLs across multiple studies. One eQTL is defined as a SNP-gene pair with significant association (FDR<0.01 for BrainSeq). The total number of gene-level cis-eQTLs in caudate is 1,506,125 (FDR < 0.01). An eQTL matches across two studies if both their SNP ID and gene ID match.

|  | **Total**<br>**significant**<br>**eQTLs** | **Number of**<br>**eQTLs**<br>**matching**<br>**caudate**<br>**eQTLs** | **Percent of**<br>**total caudate**<br>**eQTLs** | **Percent of**<br>**matching**<br>**eQTLs that**<br>**are**<br>**directionality**<br>**concordant** | **eQTL**<br>**calling**<br>**method** |
| --- | --- | --- | --- | --- | --- |
| **BrainSeq Phase**<br>**II DLPFC** | 1,577,963 | 862,718 | 57.3 | 99.6 | Matrix eQTL |
| **BrainSeq Phase**<br>**II Hippocampus** | 1,075,733 | 697,167 | 46.3 | 99.7 | Matrix eQTL |

**Table S3.** Directionality comparison of gene-level cis-eQTLs across the CommonMind Consortium (CMC) and caudate nucleus from GTEx (Genotype-Tissue Expression). One eQTL is defined as a SNP- gene pair with significant association (FDR<0.05 for CMC, and GTEx). The total number of gene-level cis-eQTLs in caudate is 2,198,553 (FDR < 0.05). An eQTL matches across two studies if both their SNP ID and gene ID match.

|  | **Total**<br>**significant**<br>**eQTLs** | **Number of**<br>**eQTLs**<br>**matching**<br>**caudate eQTLs** | **Percent of**<br>**total**<br>**caudate**<br>**eQTLs** | **Percent of matching**<br>**eQTLs that are**<br>**directionality**<br>**concordant** | **eQTL**<br>**calling**<br>**method** |
| --- | --- | --- | --- | --- | --- |
| **GTEX v7**<br>**caudate** | 435,939 | 207,175 | 9.4 | 98.5 | FastQTL |
| **CMC**<br>**DLPFC** | 2,154,319 | 672,334 | 30.6 | 98.0 | Matrix<br>eQTL |

**Table S4.** Summary of BrainSeq samples region-dependent eQTLs (genotype-brain region interaction FDR < 0.05) across features for all eQTLs and those that have SNPs associated with schizophrenia risk (GWAS p-value < 5e-8) according to the PGC2+CLOZUK GWAS.

|  |  | **Gene eQTLs**<br>**(eGenes)** | **Transcript**<br>**eQTLs**<br>**(eTranscripts)** | **Exon eQTLs**<br>**(eExons)** | **Junction**<br>**eQTLs**<br>**(eJunctions)** |
| --- | --- | --- | --- | --- | --- |
| **Caudate and**<br>**DLPFC** | **All** | 354,474<br>(6,510) | 262,003<br>(6,892) | 1,222,125<br>(35,755) | 591,503<br>(17,982) |
|  | **SZ Risk** | 4,726 (79) | 3,12 (65) | 15,653 (383) | 4,273 (155) |
| **Caudate and**<br>**hippocampus** | **All** | 204,016<br>(4,710) | 118,733<br>(3,713) | 692,897<br>(23,175) | 397,816<br>(12,518) |
|  | **SZ Risk** | 2,750 (45) | 2,016 (40) | 9,833 (222) | 3,691 (100) |
| **DLPFC and**<br>**hippocampus** | **All** | 86,980<br>(2,247) | 37,026<br>(1,114) | 201,900<br>(6,545) | 101,781<br>(3,619) |
|  | **SZ Risk** | 622 (16) | 171 (7) | 1,292 (56) | 381 (25) |

**Table S5.** Transcriptome-wide association study (TWAS) feature summary for caudate nucleus across multiple genomic features (genes, transcripts, exons, exon-exon junctions). Heritable p-value cutoff set to 0.01.

|  | **Genes** | **Transcripts** | **Exons** | **Junctions** |
| --- | --- | --- | --- | --- |
| **Heritable features ($h^2$)** | 5256 | 9735 | 39467 | 14226 |
| **TWAS (FDR < 0.05)** | 489 | 933 | 3907 | 1419 |
| **TWAS Geneid (FDR < 0.05)** | 489 | 697 | 1012 | 589 |

**Table S6.** Summary of top 25 most significant TWAS associations (by TWAS p-value), but not significant in the PGC2 GWAS (GWAS p-value > 5e-8) specific to the caudate nucleus.

| GENE | SNP | CHR | TWAS FDR | GWAS P-value |
| --- | --- | --- | --- | --- |
| ANKRD45 | rs6695327 | 1 | 8.4e-5 | 1.3e-6 |
| LRRC4 | rs6954161:127739721:G:<br>T | 7 | 1.1e-4 | 3.6e-7 |
| KLHL21 | rs200448 | 1 | 2.0e-4 | 5.4e-8 |
| ENSG00000271858 | rs2624847 | 3 | 2.9e-4 | 1.6e-5 |
| STRC | rs11070410:44057045:A:<br>G | 15 | 2.9e-4 | 4.0e-6 |
| CEPT1 | rs1389724 | 1 | 4.2e-4 | 3.3e-7 |
| ANKRD36 | rs2271893 | 2 | 5.7e-4 | 2.1e-5 |
| ENSG00000257180 | rs12449216 | 16 | 7.2e-4 | 1.0e-6 |
| LINC00862 | rs10919967 | 1 | 1.1e-3 | 5.5e-6 |
| PPIL2 | rs5749696 | 22 | 1.2e-3 | 1.4e-5 |
| ENSG00000260517 | rs6565300:28993049:A:C | 16 | 1.3e-3 | 1.4e-3 |
| SYNDIG1L | rs8009761 | 14 | 1.3e-3 | 6.0e-5 |
| RAPH1 | rs16839296 | 2 | 1.4e-3 | 2.6e-4 |
| CTXN3 | rs1421750 | 5 | 1.9e-3 | 1.2e-5 |
| ADAM10 | rs653765 | 15 | 1.9e-3 | 1.6e-6 |
| ENSG00000236723 | rs17785382 | 1 | 2.1e-3 | 1.2e-6 |
| ENSG00000258695 | rs2269967 | 14 | 2.1e-3 | 1.2e-4 |
| ENSG00000231615 | rs12561796 | 1 | 2.5e-3 | 1.8e-6 |
| MFHAS1 | rs330943 | 8 | 2.6e-3 | 6.8e-5 |
| IFT57 | rs3804639 | 3 | 2.8e-3 | 3.9e-5 |
| TDRD6 | rs12215229 | 6 | 2.8e-3 | 5.1e-5 |
| DNPH1 | rs6938026 | 6 | 2.9e-3 | 6.4e-7 |
| MIAT | rs5752351:26990807:A:<br>G | 22 | 3.4e-03 | 2.1e-4 |
| GAP43 | rs6438256:115130198:C:<br>A | 3 | 3.5e-3 | 1.8e-4 |
| CDC42 | rs11582542 | 1 | 3.7e-3 | 4.2e-6 |

**Table S7.** Summary of differentially expressed features in schizophrenia versus control in the caudate nucleus (FDR < 0.05).

|  | Genes | Transcripts | Exons | Junctions |
| --- | --- | --- | --- | --- |
| DE Features | 2,699 | 1,908 | 22,816 | 7,470 |
| DE Geneid | 2,699 | 1,607 | 3,832 | 2,066 |

**Table S8.**
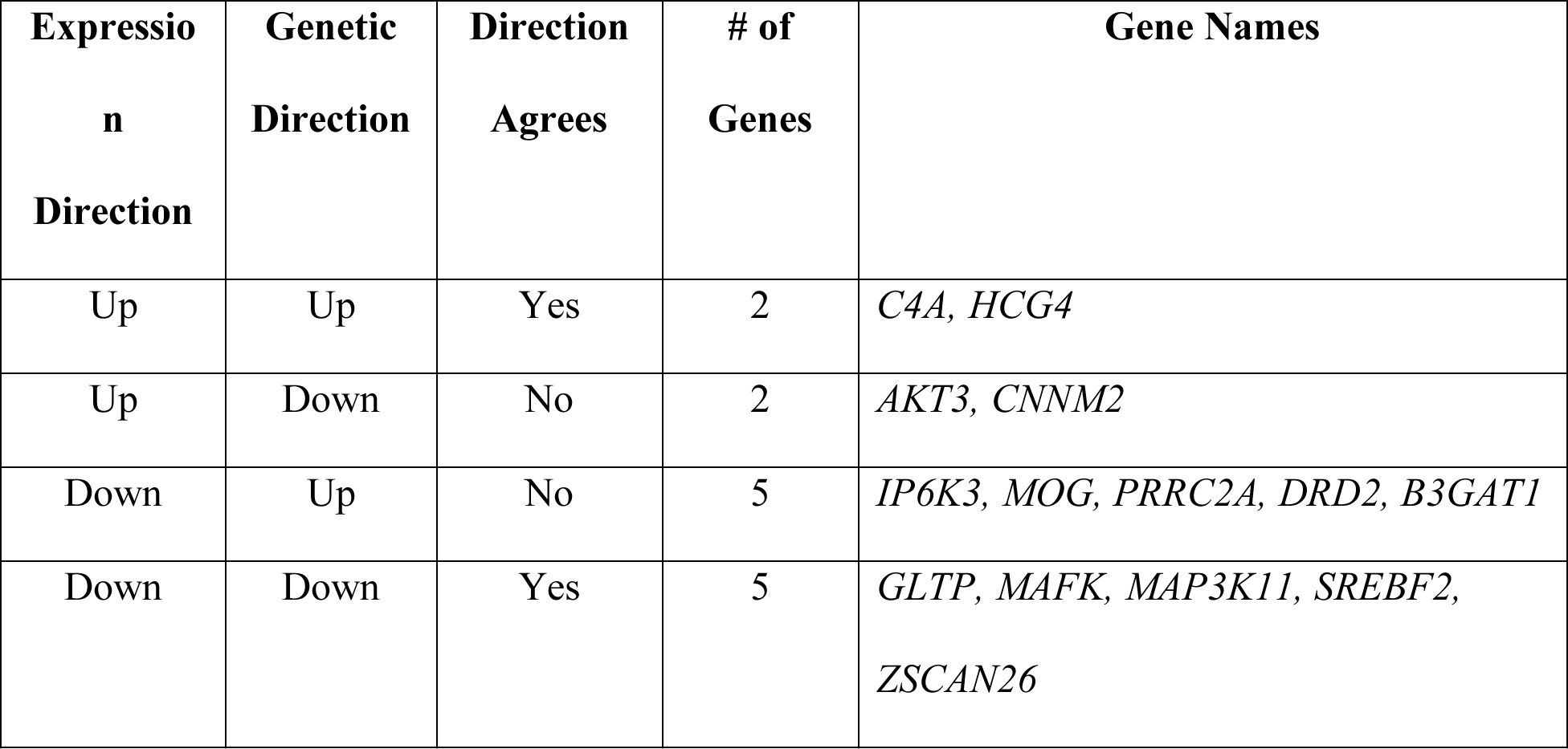
Summary of the intersection between GWAS (P-value < 5e-8), DE (FDR < 0.05), and eQTL (FDR < 0.05) for **transcripts**.

| **Expressio**<br>**n**<br>**Direction** | **Genetic**<br>**Direction** | **Direction**<br>**Agrees** | **# of**<br>**Genes** | **Gene Names** |
| --- | --- | --- | --- | --- |
| Up | Up | Yes | 2 | *C4A, HCG4* |
| Up | Down | No | 2 | *AKT3, CNNM2* |
| Down | Up | No | 5 | *IP6K3, MOG, PRRC2A, DRD2, B3GAT1* |
| Down | Down | Yes | 5 | *GLTP, MAFK, MAP3K11, SREBF2,*<br>*ZSCAN26* |

**Table S9.** Summary of the intersection between GWAS (P-value < 5e-8), DE (FDR < 0.05), and eQTL (FDR < 0.05) for **exons**.

| **Expression Direction** | **Genetic Direction** | **Direction Agrees** | **# of Genes** | **Gene Names** |
| --- | --- | --- | --- | --- |
| Up | Up | Yes | 17 | *AL049840.5, C4A, C4B, CLU, CSDC2, DNAJC19, HCG4, HLA-DRB5, HYDIN, LINC10470, MPP6, NDUFA6, REEP2, SNAP91, SUGP2, ZFYVE21, ZNF14* |
| Up | Down | No | 7 | *CNNM2, DRD2, ENSA, GRIN2A, ME1, NGEF, SLC35G2* |
| Down | Up | No | 11 | *ATF6B, ATP6V1G2, B3GAT1, BAG6, CPEB1, IP6K3, IP013, PRRC2A, SATB2, VRK2, ZC3H7B* |
| Down | Down | Yes | 19 | *AMBRA1, ASPHD1, BRD2, CACNA1I, CKB, FANCL, GTF2H4, HLA-C, MAFK, MAP3K11, NRIP2, PCCB, PPM1M, PLPP5, SREBF2, TWF2, YPEL4, ZSCAN26, ZKSCAN8* |

**Table S10.**
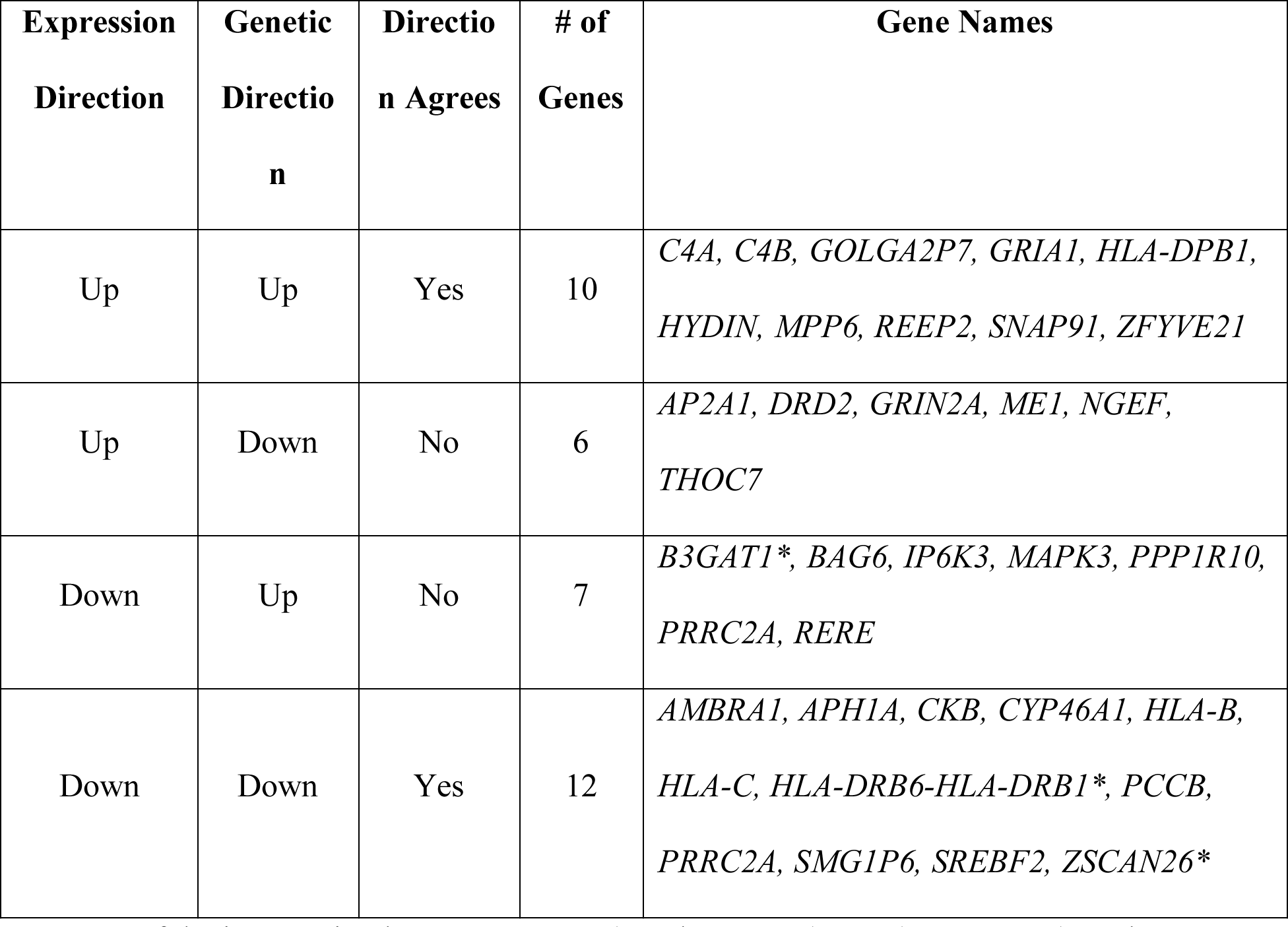
Summary of the intersection between GWAS (P-value < 5e-8), DE (FDR < 0.05), and eQTL (FDR < 0.05) for **exon-exon junctions**. * denotes unannotated junction within gene.

| **Expression Direction** | **Genetic Direction** | **Direction Agrees** | **# of Genes** | **Gene Names** |
| --- | --- | --- | --- | --- |
| Up | Up | Yes | 10 | *C4A, C4B, GOLGA2P7, GRIA1, HLA-DPB1, HYDIN, MPP6, REEP2, SNAP91, ZFYVE21* |
| Up | Down | No | 6 | *AP2A1, DRD2, GRIN2A, ME1, NGEF, THOC7* |
| Down | Up | No | 7 | *B3GAT1\*, BAG6, IP6K3, MAPK3, PPP1R10, PRRC2A, RERE* |
| Down | Down | Yes | 12 | *AMBRA1, APH1A, CKB, CYP46A1, HLA-B, HLA-C, HLA-DRB6-HLA-DRB1\*, PCCB, PRRC2A, SMG1P6, SREBF2, ZSCAN26\** |

**Table S11.**
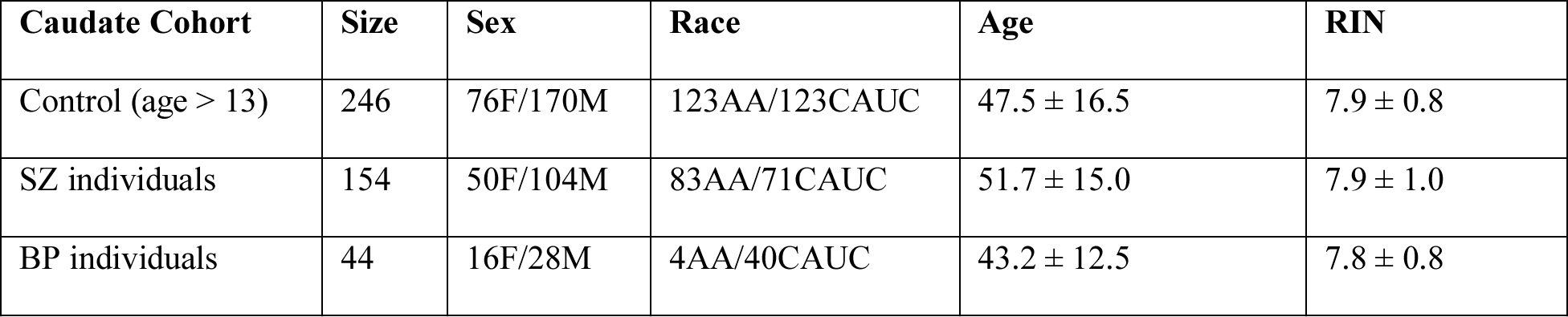
Demographic information for caudate nucleus samples. AA, African American; CAUC, Caucasian American; F, Female; M, Male; SZ, Schizophrenia; BP, Bipolar Disorder; RIN, RNA Integrity Number

| **Caudate Cohort** | **Size** | **Sex** | **Race** | **Age** | **RIN** |
| --- | --- | --- | --- | --- | --- |
| Control (age > 13) | 246 | 76F/170M | 123AA/123CAUC | 47.5 ± 16.5 | 7.9 ± 0.8 |
| SZ individuals | 154 | 50F/104M | 83AA/71CAUC | 51.7 ± 15.0 | 7.9 ± 1.0 |
| BP individuals | 44 | 16F/28M | 4AA/40CAUC | 43.2 ± 12.5 | 7.8 ± 0.8 |

