## Supplementary figures and images for "Genetic and environmental regulation of caudate nucleus transcriptome: insight into schizophrenia risk and the dopamine system"

### module0_go_enrichment.pdf

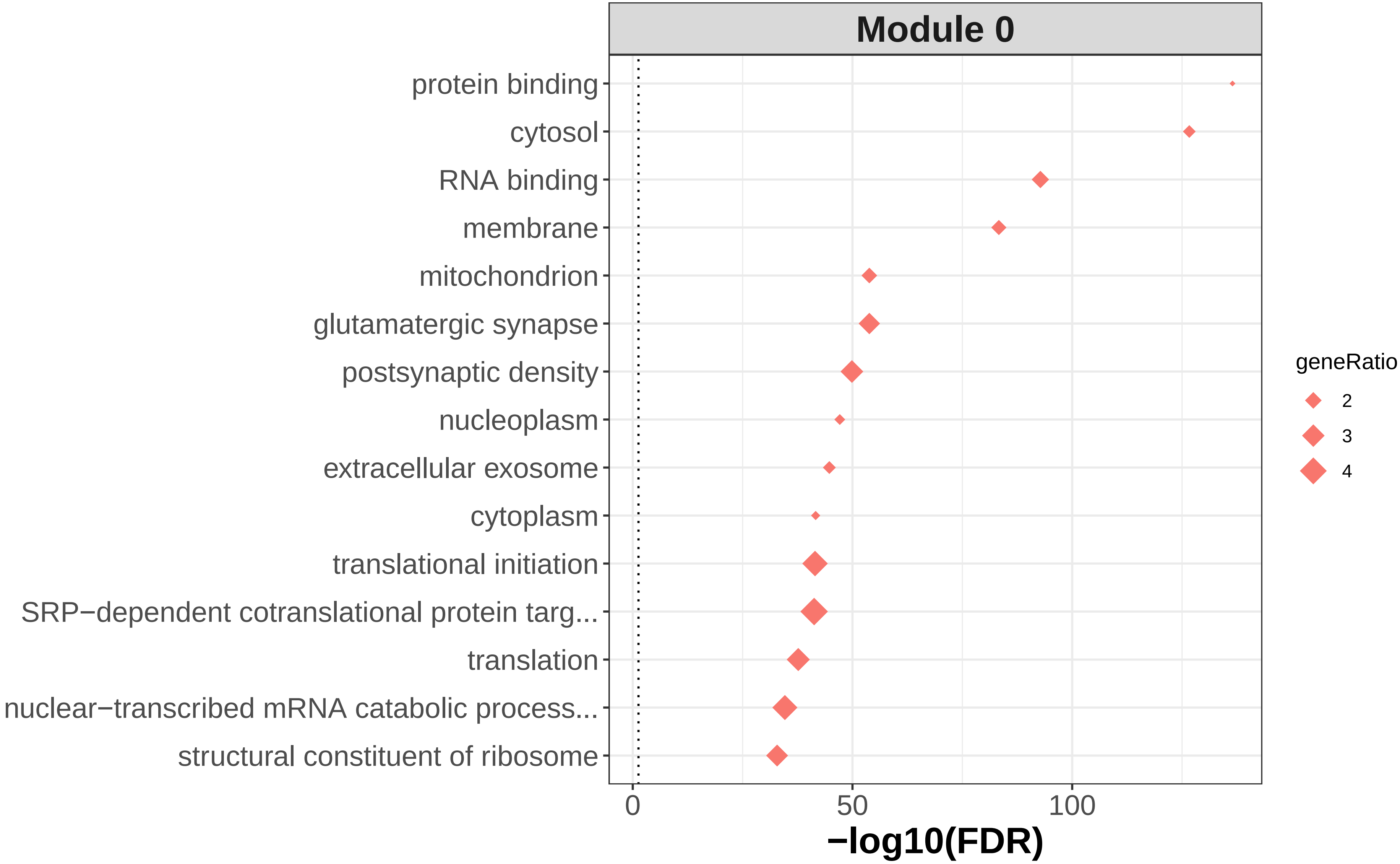

### module0_go_wordcloud.png

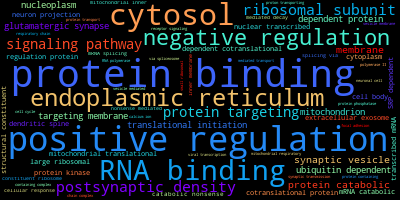

### module1_go_enrichment.pdf

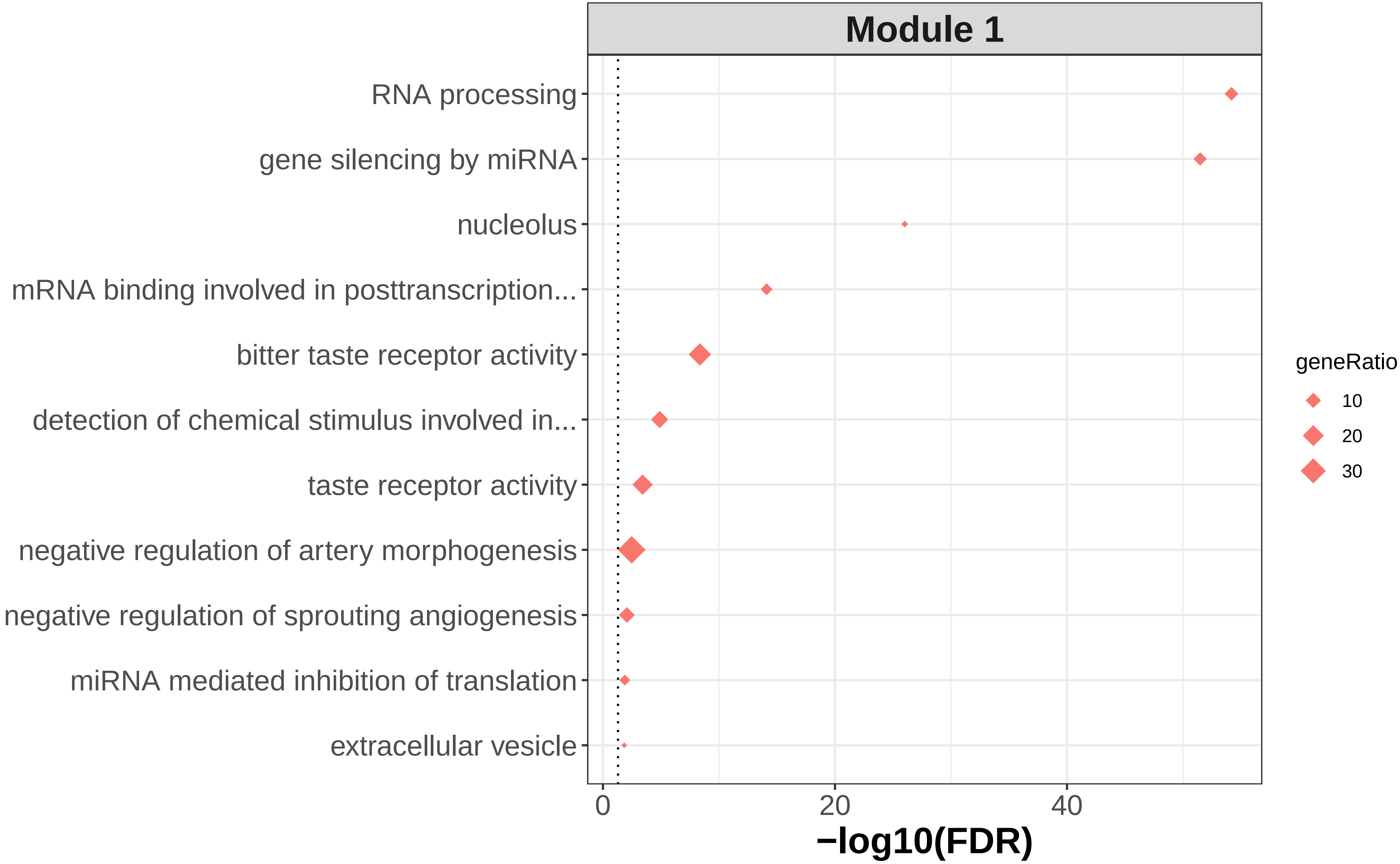

### module1_go_wordcloud.png

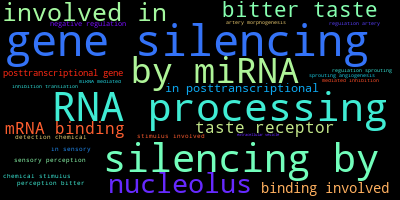

### module2_go_enrichment.pdf

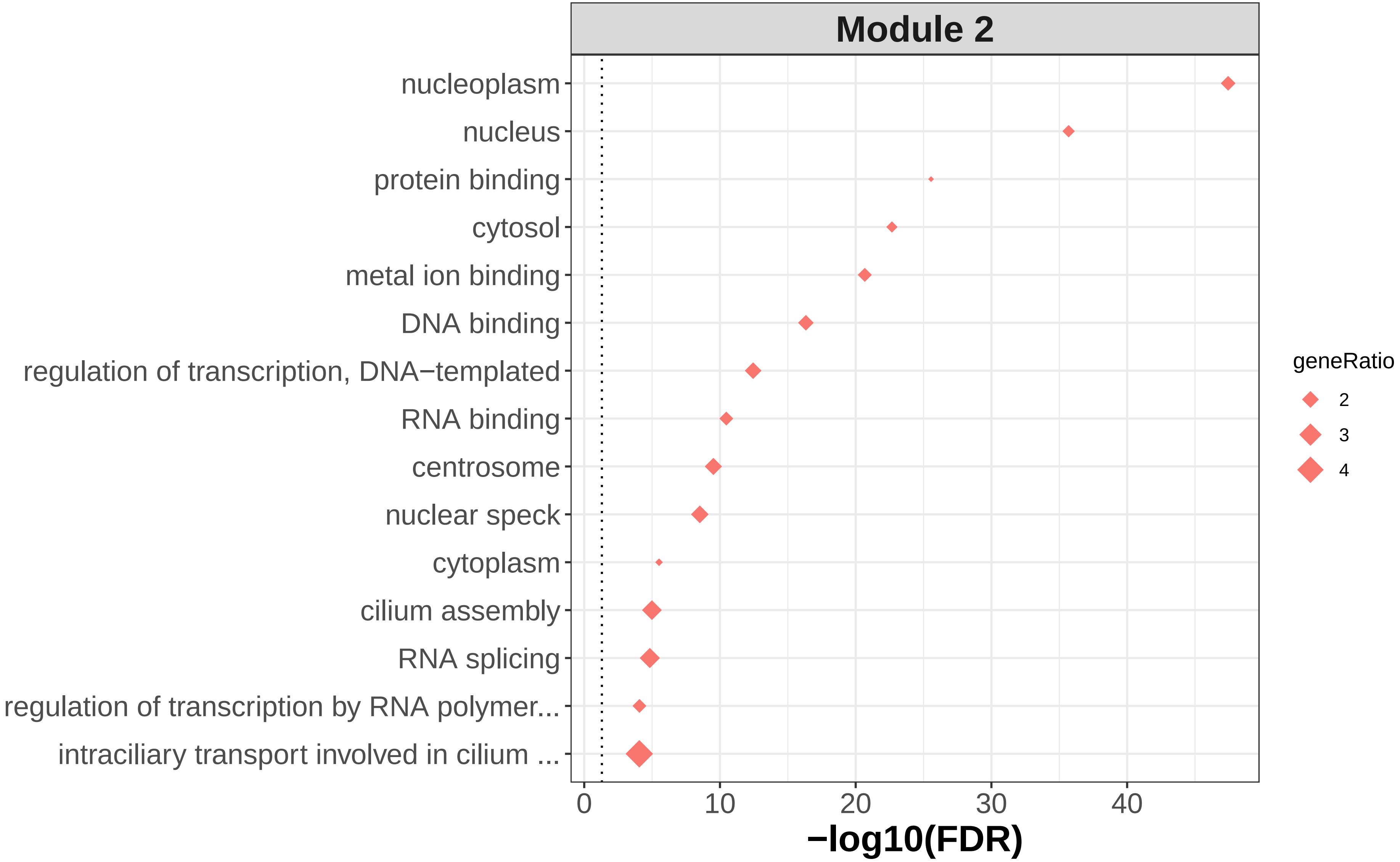

### module2_go_wordcloud.png

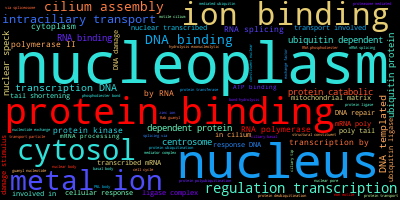

### module3_go_enrichment.pdf

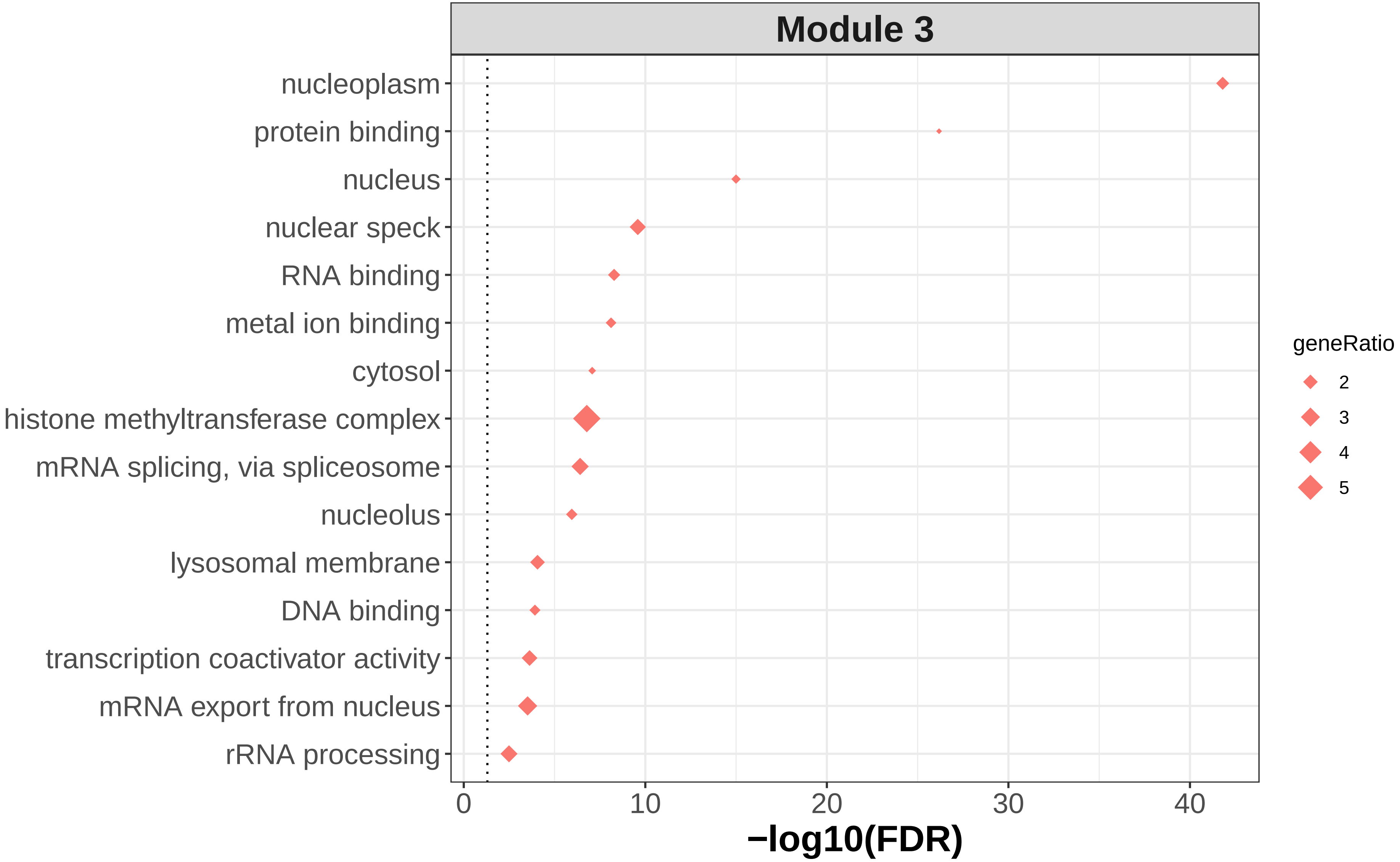

### module3_go_wordcloud.png

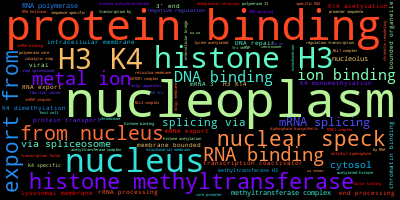

### module4_go_enrichment.pdf

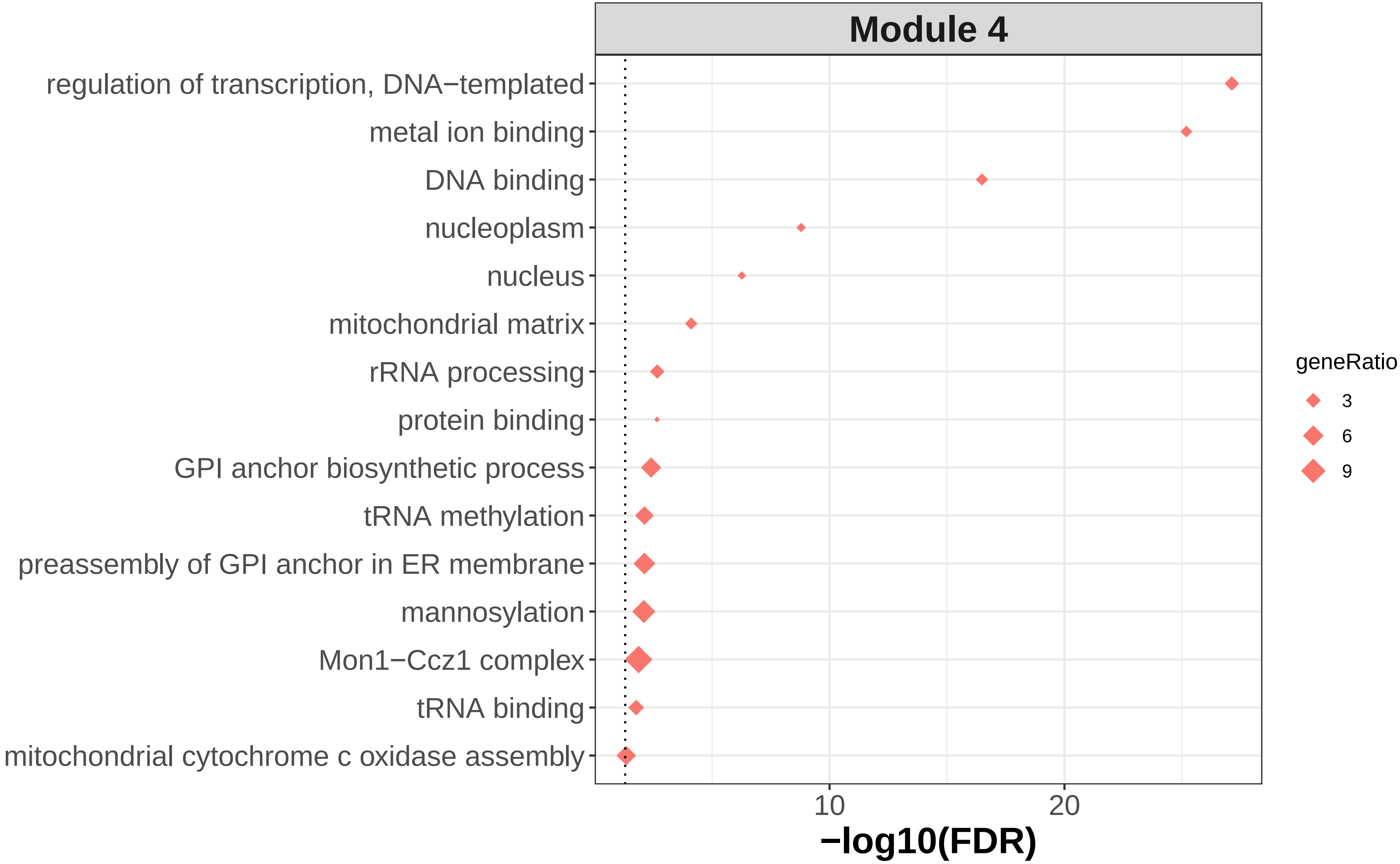

### module4_go_wordcloud.png

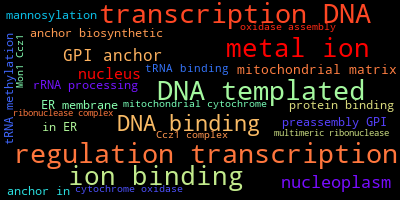

### module5_go_enrichment.pdf

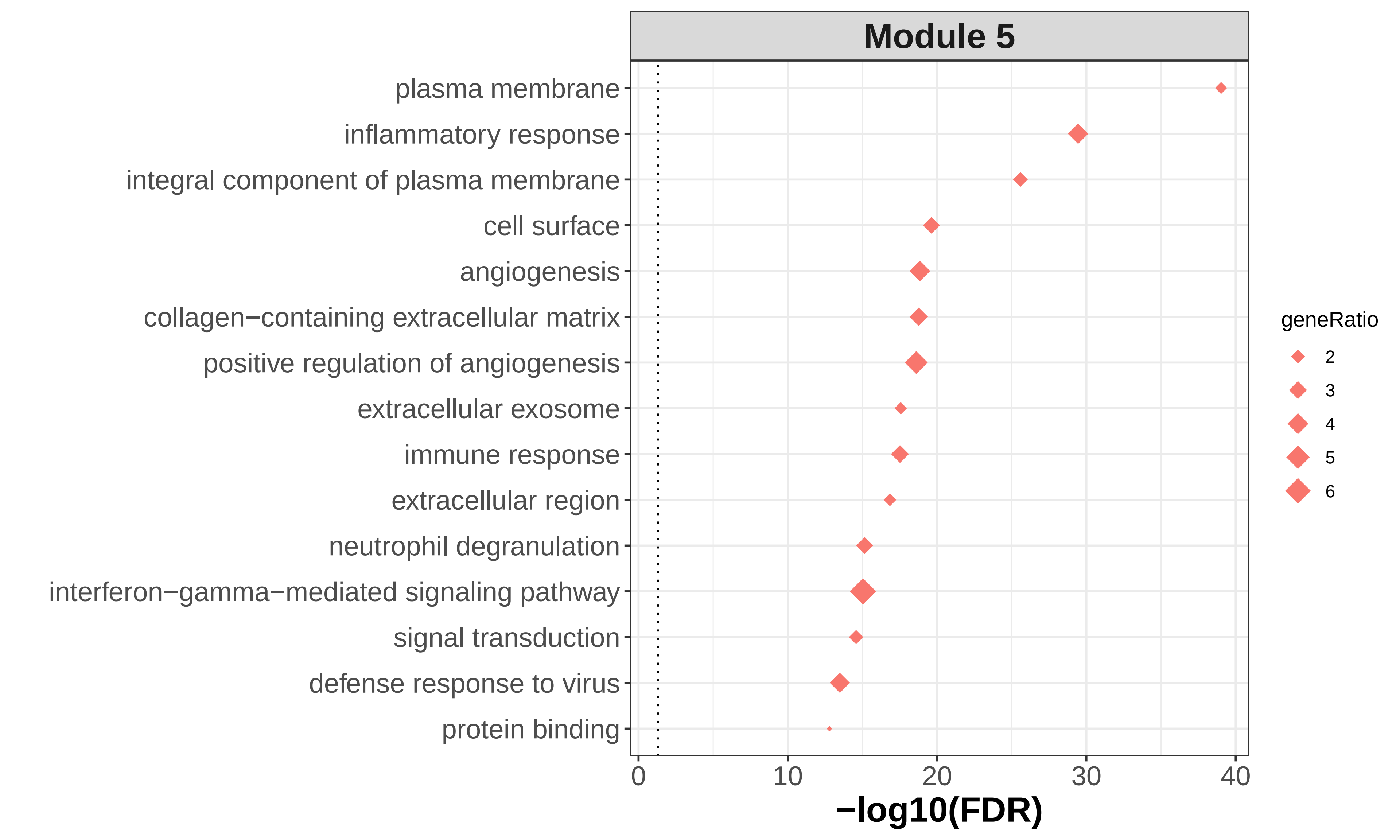

### module5_go_wordcloud.png

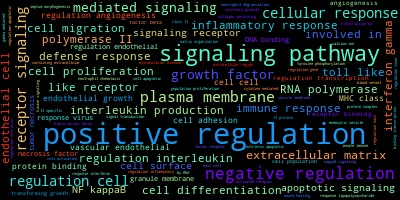

### module6_go_enrichment.pdf

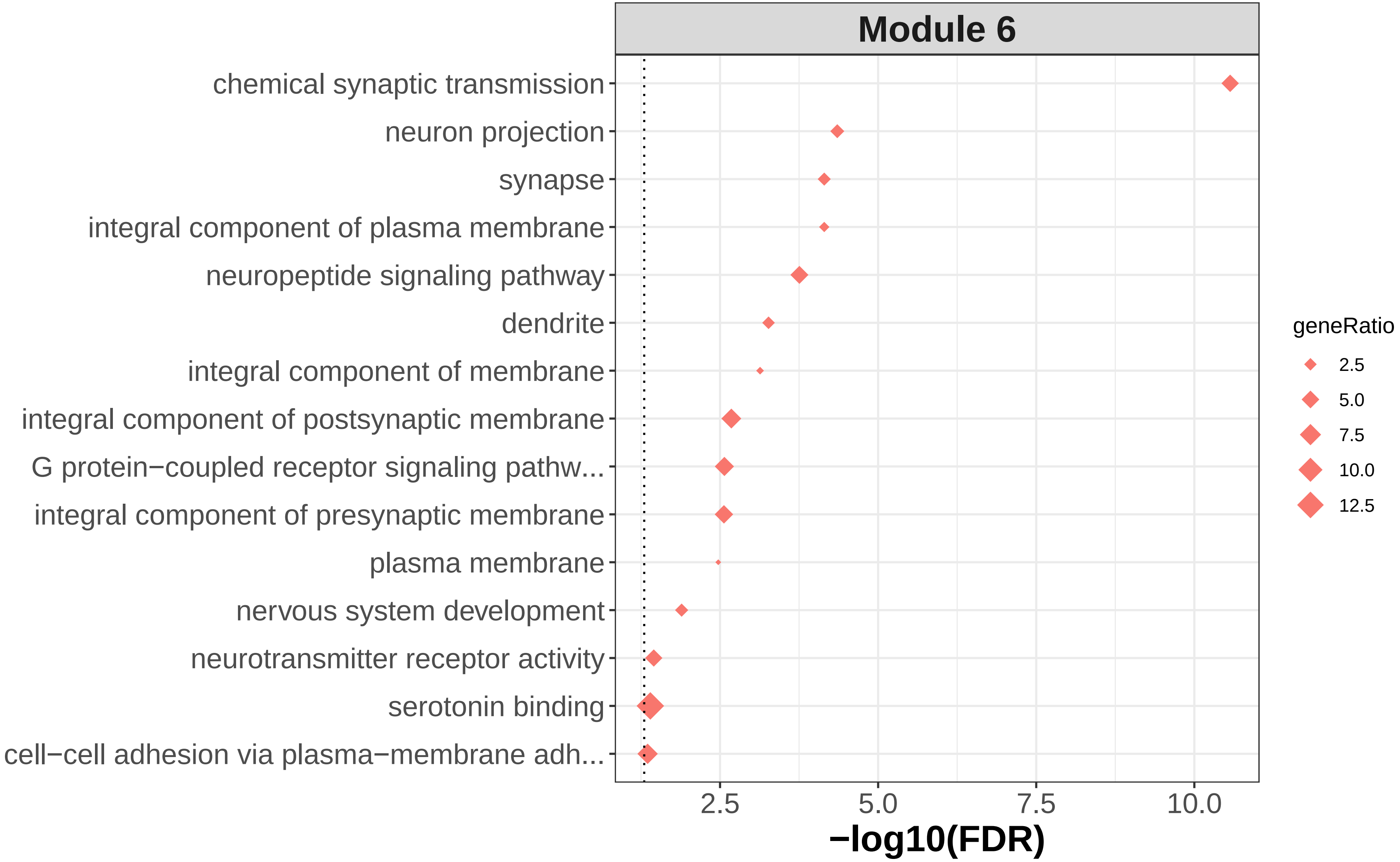

### module6_go_wordcloud.png

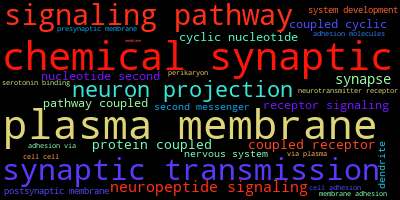

### module7_go_enrichment.pdf

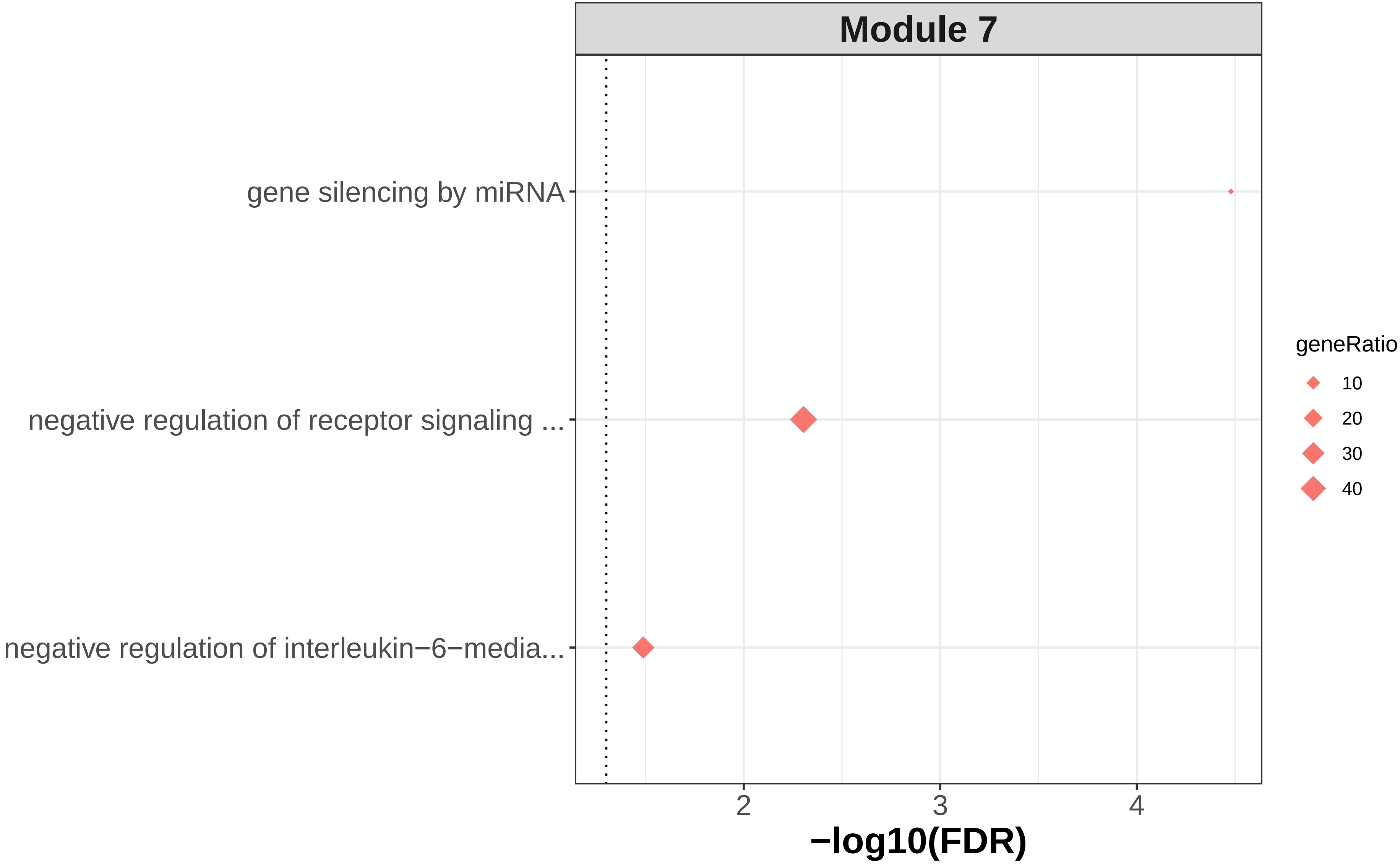

### module7_go_wordcloud.png

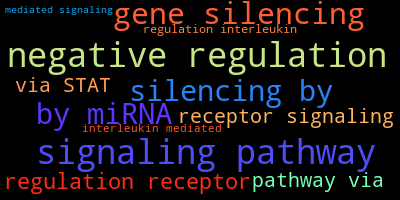

### module9_go_enrichment.pdf

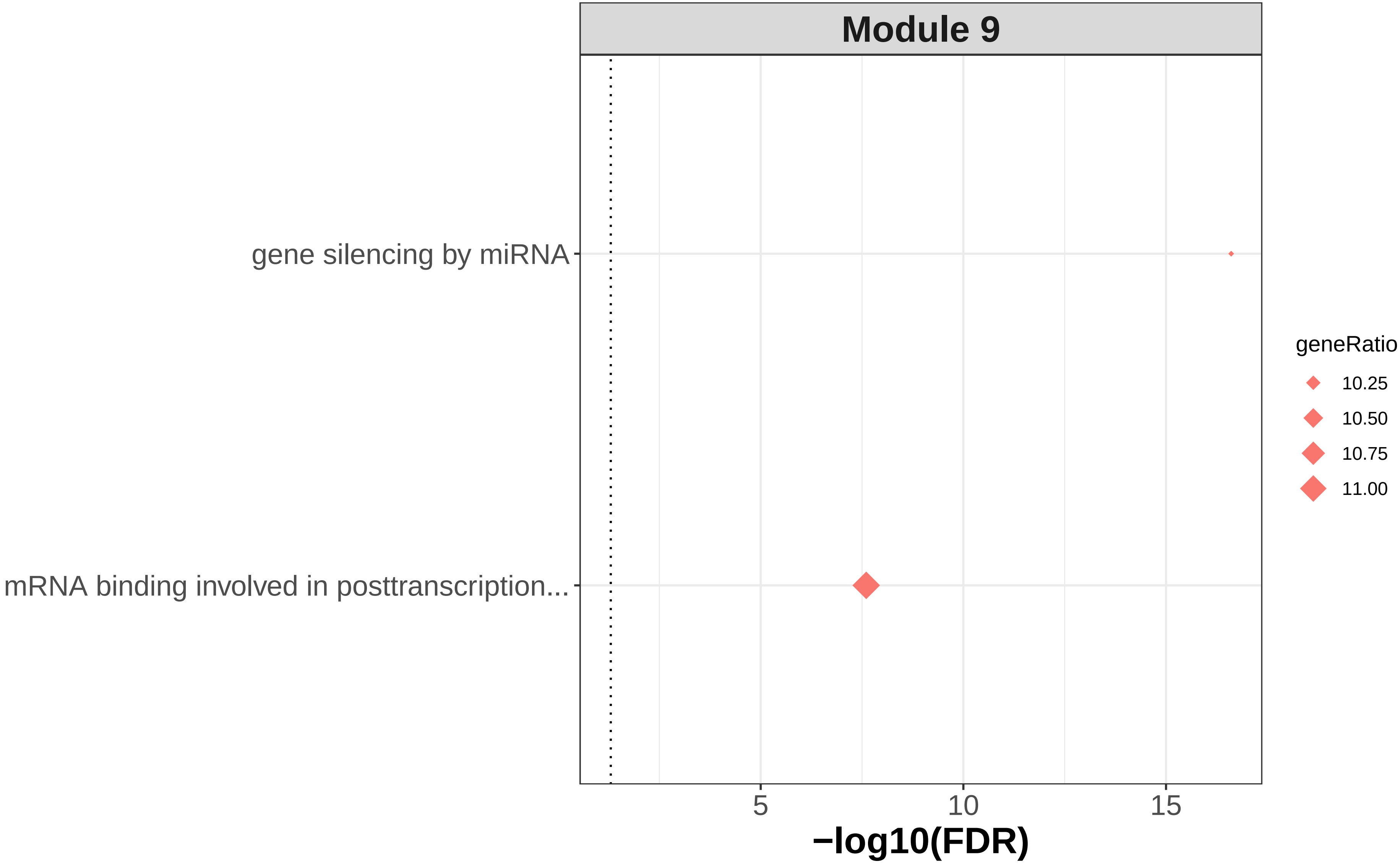

### module9_go_wordcloud.png

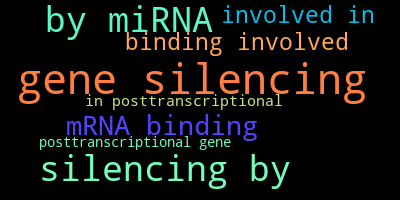

### module11_go_enrichment.pdf

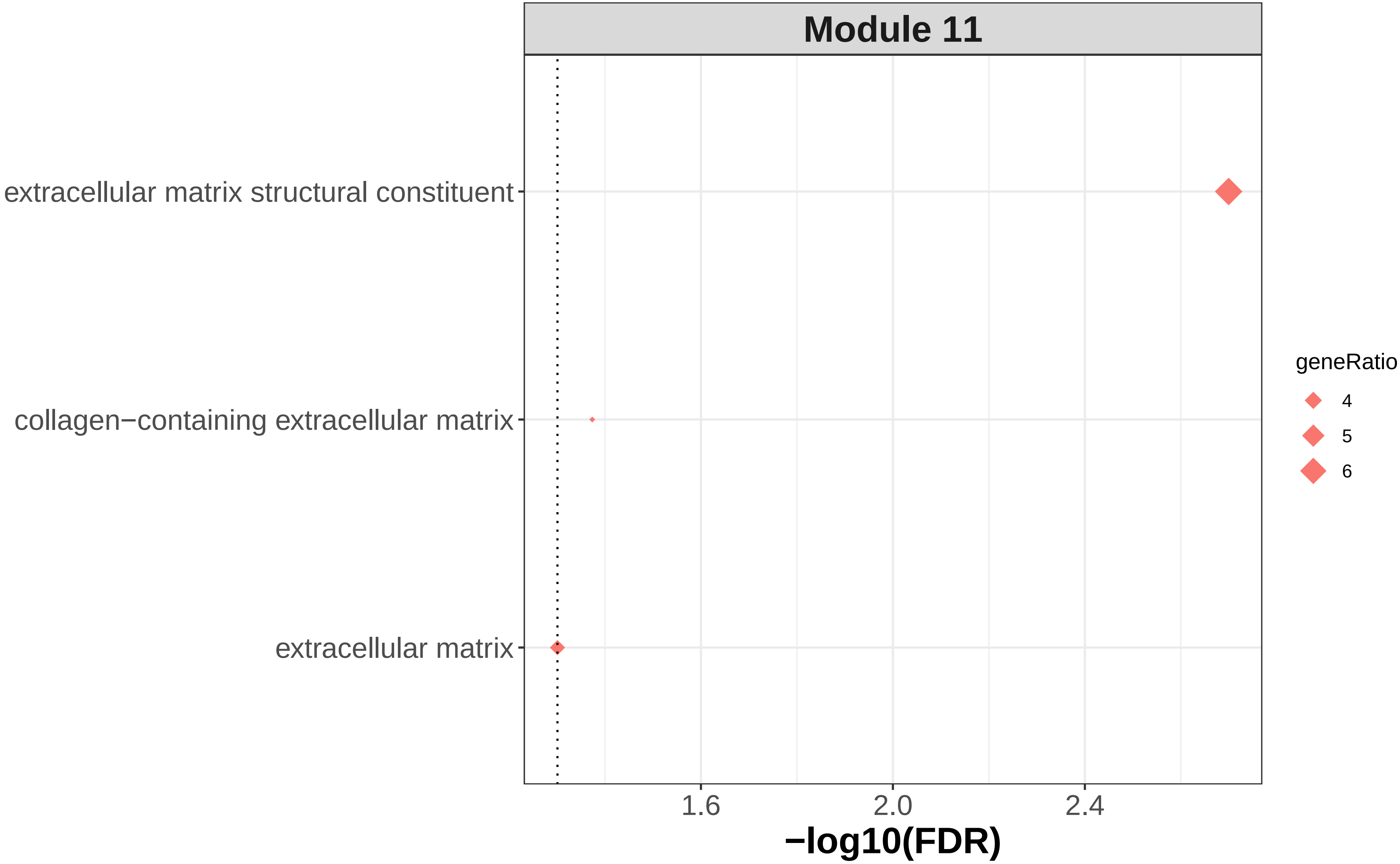

### module11_go_wordcloud.png

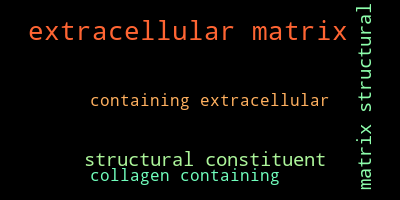

### module12_go_enrichment.pdf

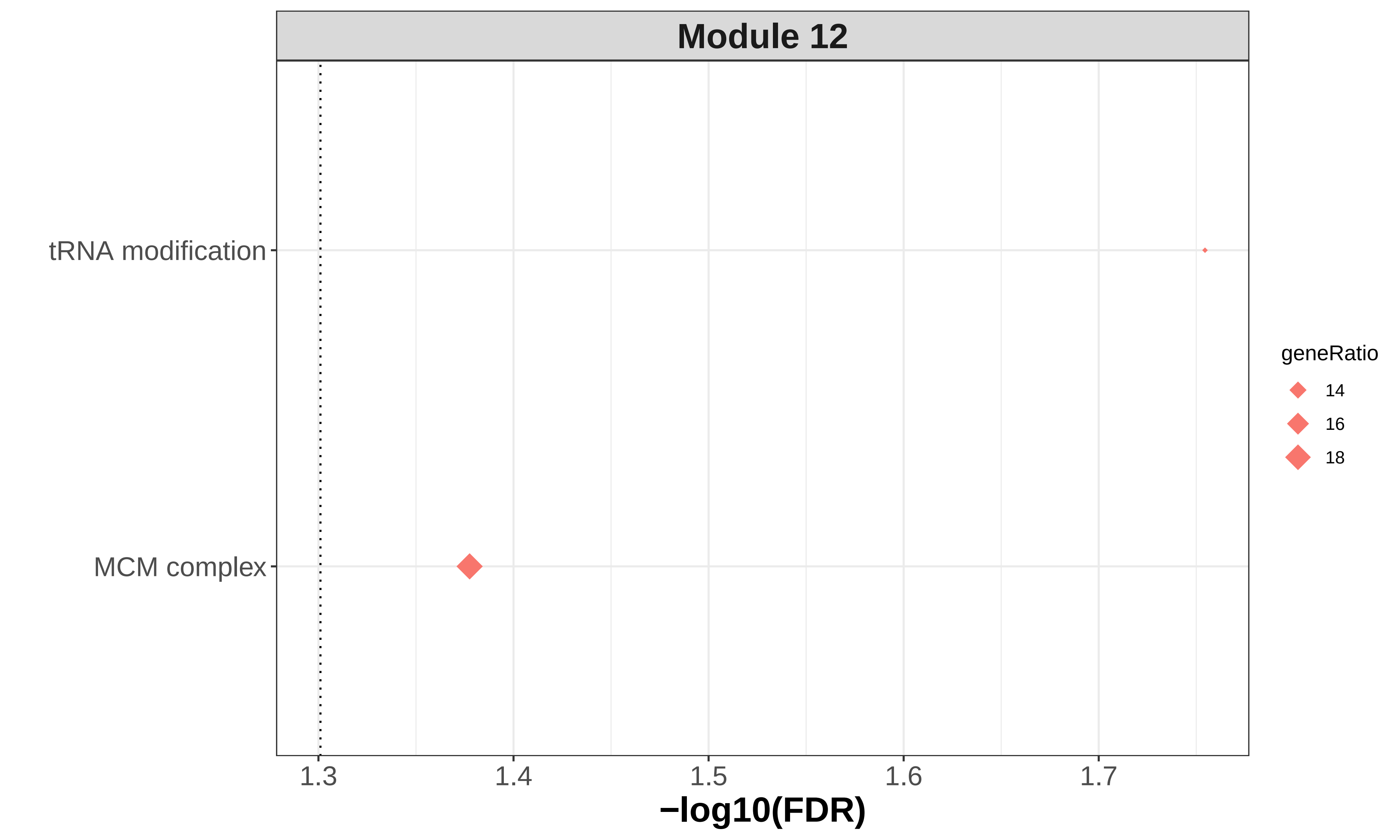

### module12_go_wordcloud.png

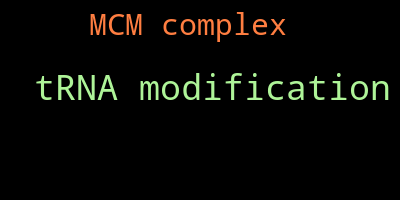

### module13_go_enrichment.pdf

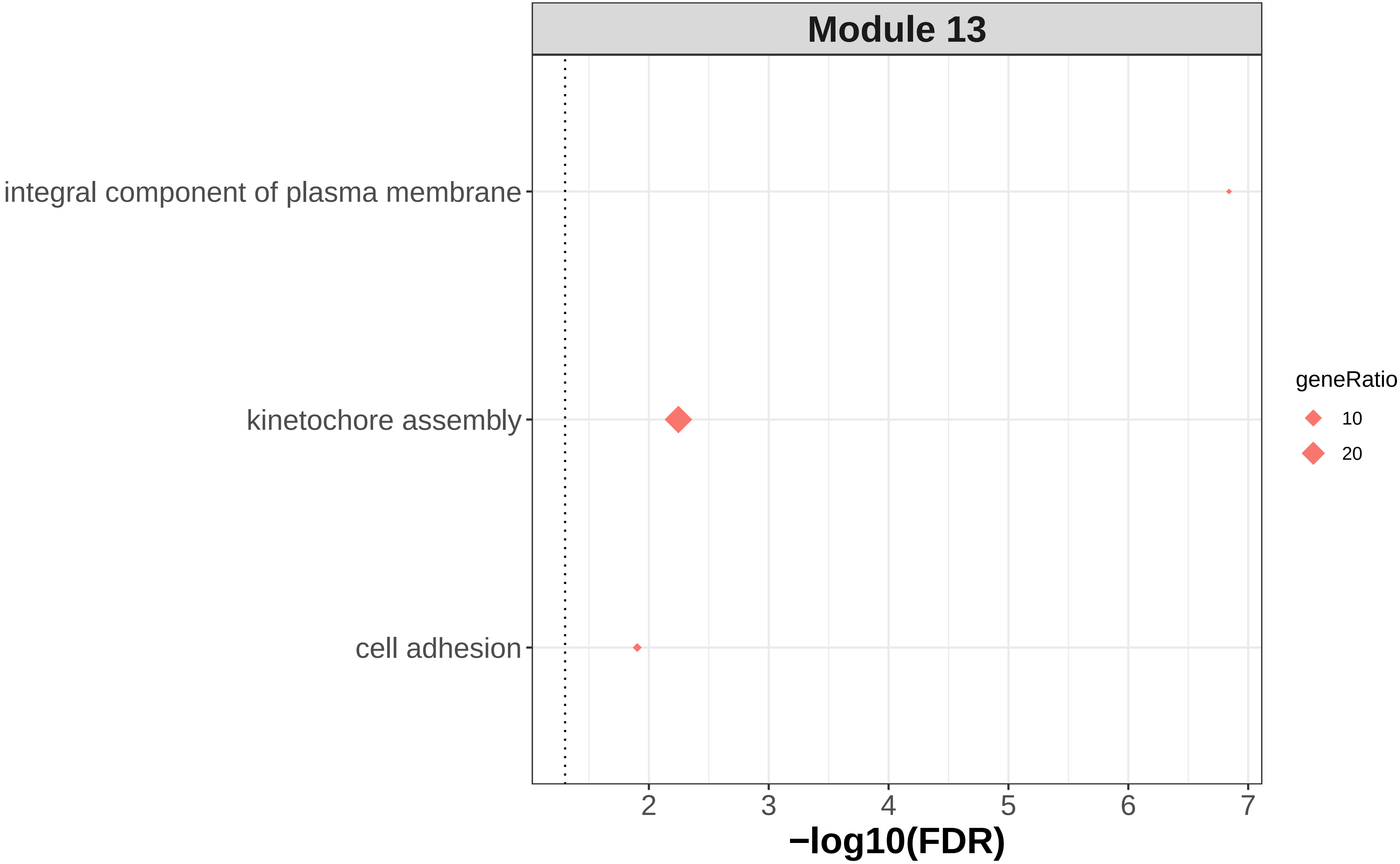

### module13_go_wordcloud.png

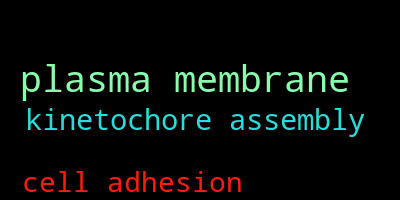

### module14_go_enrichment.pdf

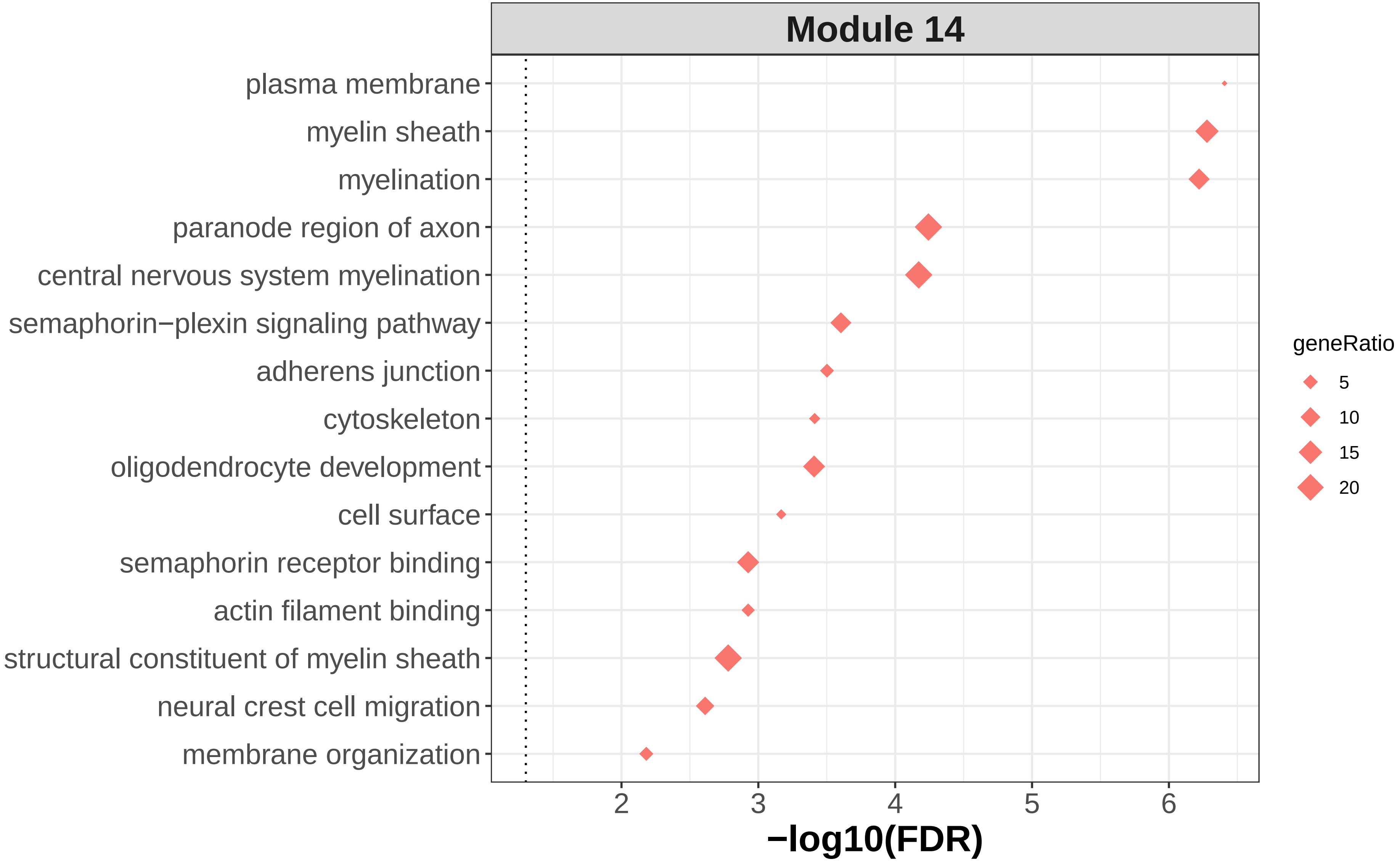

### module14_go_wordcloud.png

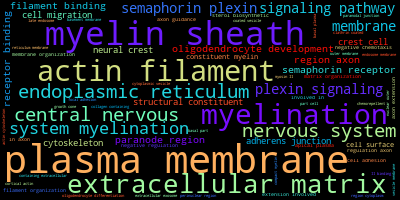

### module16_go_enrichment.pdf

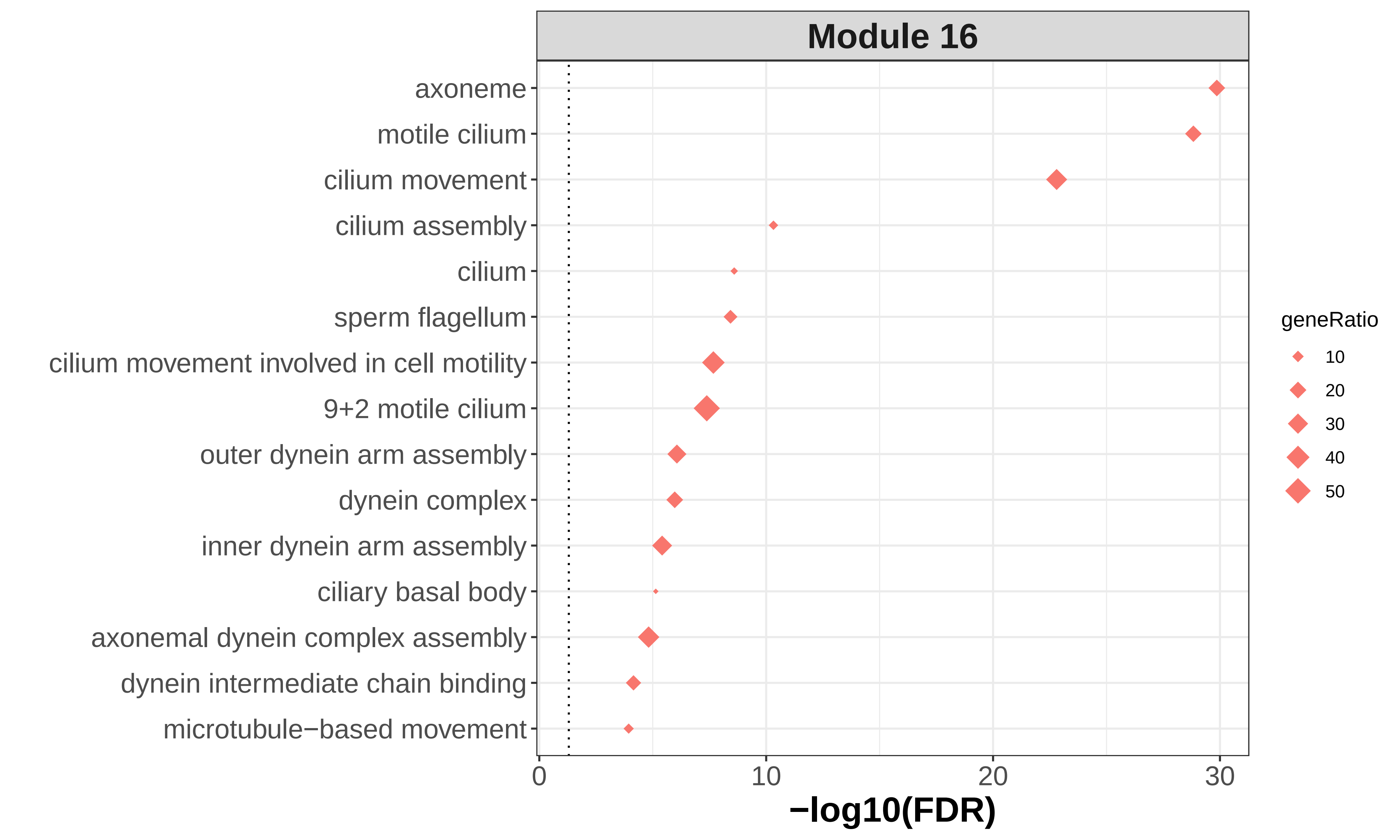

### module16_go_wordcloud.png

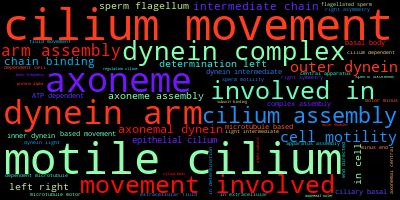

### module19_go_enrichment.pdf

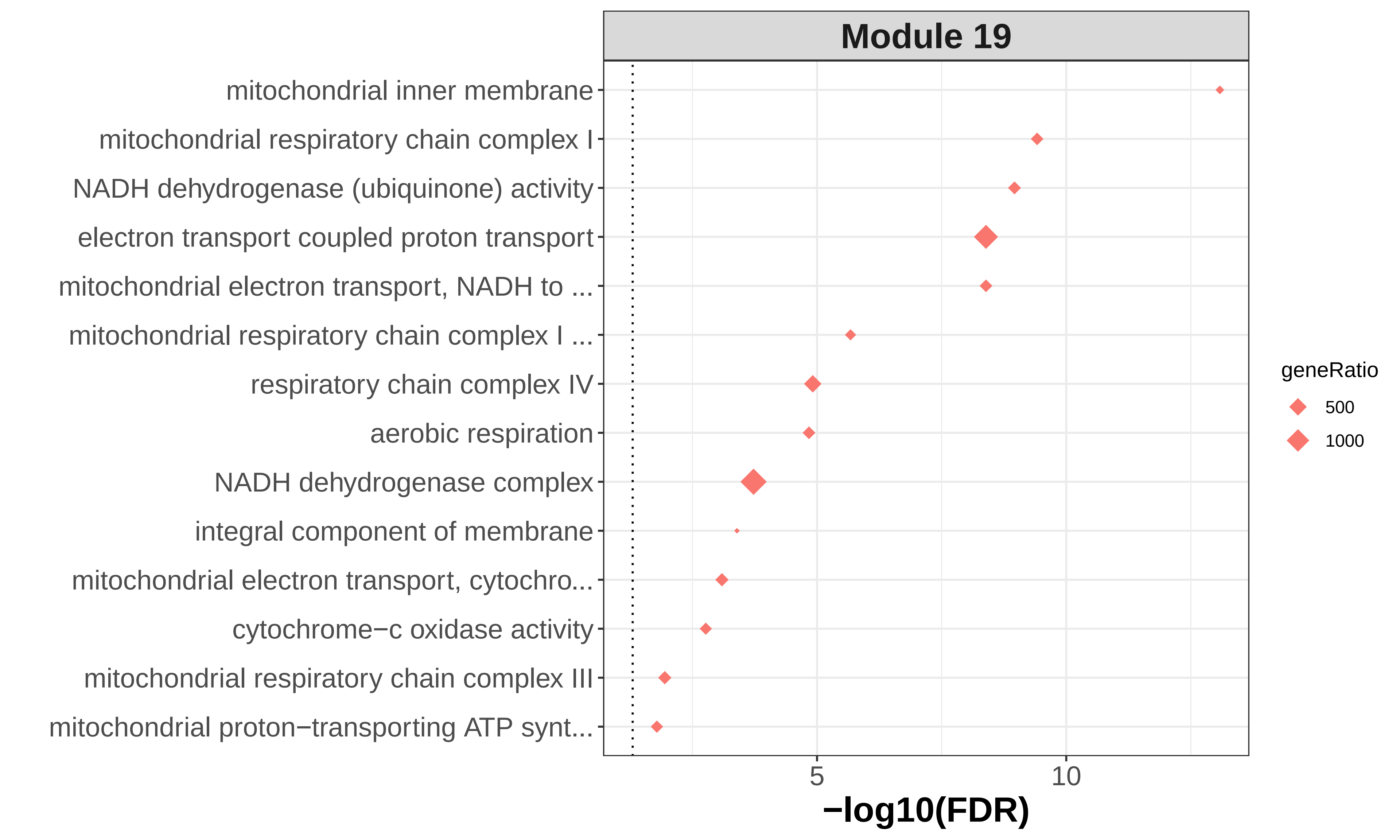

### module19_go_wordcloud.png

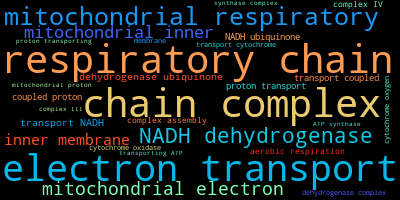
